# Genomic Landscape of Thrombosis Recurrence Risk Across Venous Thromboembolism Subtypes

**DOI:** 10.1101/2024.12.02.24317788

**Authors:** Gaëlle Munsch, Florian Thibord, Ohanna C Bezerra, Jennifer A. Brody, Astrid van Hylckama Vlieg, Lénaïck Gourhant, Ming-Huei Chen, Marine Germain, Ilana Caro, Pierre Suchon, Robert Olaso, Kerri L. Wiggins, Noémie Saut, Céline Besse, Louisa Goumidi, Delphine Bacq, Laura B Harrington, Anne Boland, CHARGE Hemostasis working group, INVENT consortium, Catherine A Lemarié, Sven Danckwardt, Stéphanie Debette, Jean-François Deleuze, Hélène Jacqmin-Gadda, Marc A Rodger, France Gagnon, Frits R Rosendaal, Andrew D Johnson, Nicholas L Smith, Francis Couturaud, Pierre-Emmanuel Morange, David-Alexandre Trégouët

## Abstract

Venous thromboembolism (VT) is a frequent (annual incidence of 1 to 2 per 1,000) and potentially life-threatening (case-fatality rate up to 10%) disease. VT is associated with serious short-term and long-term complications including a recurrence rate of approximately 20% within five years. Anticoagulant therapy, the mainstay of VT treatment, drastically reduces the risk of early VT recurrence, but it exposes patients to a substantial risk of bleeding. We analysed the genomic architecture of VT recurrence using data from 6,571 patients across eight cohorts, 1,816 of whom experienced recurrence, with a particular focus on the clinical manifestation of the type of first VT event. Through genome-wide association studies (GWAS), we identified three loci significantly associated (P<5×10^-8^) with VT recurrence in the general VT population: *GPR149/MME*, *L3MBTL4*, and *THSD7B*. Protein Quantitative Trait Locus and Mendelian Randomization analyses further identified elevated plasma levels of coagulation factor XI and GOLM2 as risk factors for recurrence, while decreased levels of PCSK9 and pro-IL16 were linked to reduced VT recurrence risk.

Subgroup analyses revealed 18 loci associated with VT recurrence, with notable differences between pulmonary embolism (PE) and deep vein thrombosis (DVT). For example, the exonic variant *SLC4A1* p.Glu40Lys was significantly associated with recurrence in PE patients (Hazard Ratio (HR)=3.23, P=9.7×10^-12^) but showed no effect in DVT (HR=1.00, P=0.98).

These findings emphasize the role of specific genetic loci and protein pathways in influencing VT recurrence and provide valuable insights into potential therapeutic targets. Further research is needed to clarify the biological mechanisms driving these associations.

**Key points:**

- 29 loci/proteins associated with VT recurrence risk.
- The genomic architecture of VT recurrence risk varies based on the initial clinical presentation.

## Introduction

Venous thromboembolism (VT) encompasses deep vein thrombosis (DVT) and pulmonary embolism (PE). With an incidence of 1 to 2 per 1,000 adults annually and a mortality of about 10%, VT is the third most common cause of cardiovascular death worldwide^1,2^. The mainstay of treatment is anticoagulant therapy, which should be administered for a minimum of three months to prevent early recurrence and death^3^. After discontinuation, the risk of recurrent VT is variable, depending on the presence of major factors at the time of VT. When VT is provoked by a major transient risk factor (*e.g*., surgery, immobilisation), the risk of recurrent VT is low (<2% at one year) and anticoagulation should not be continued^3^. For VT diagnosed in the context of cancer, therapeutic anticoagulation is needed for as long as the cancer is considered active^3^. However, in more than 50% of cases, VT occurs in absence of any major risk factors (termed unprovoked VT)^4^. In these cases, more than 35% will develop recurrent VT if anticoagulation is stopped, with a case-fatality rate of about 10% ^3,5–7^. Consequently, international guidelines recommend to treat patients with a first episode of unprovoked VT “indefinitely” ^3^. However, such practice exposes patients to a substantial linear increase in the risk of bleeding^5,7^ and proposing indefinite anticoagulation in all patients with unprovoked VT exposes 65% of patients, who would never have recurrence after anticoagulation discontinuation, to an unjustified risk of bleeding. Hence, determining the optimal duration of anticoagulant treatment for VT is a major public health issue^8^. For this purpose, a better understanding of the mechanisms involved in VT recurrence is crucial for improving prevention and identifying patients who may not require extended anticoagulation.

Established risk factors for VT recurrence currently established are: unprovoked first VT, elevated D-dimer levels and male sex^9^. Several biological clinical scores have been developed in order to predict the risk of VT recurrence and to identify who should stop or continue anticoagulants (*i.e.*, HERDOO2, DASH, VIENNA, L-TRRiP, and VTE-PREDICT scorers)^10–14^. However, the discriminant power of these scores is moderate and their use in current practice is limited. While several scores consider D-dimers levels as a predictor of VT recurrence, other biological candidate risk factors such as coagulation factor XI (FXI), coagulation factor VIII (FVIII), von Willebrand factor (vWF) and tissue factor pathway Inhibitor (TFPI) have been identified but reproducibility issues limit their use and they may not be independent^15–19^.

The observations that, in patients with unprovoked VT, the risk of recurrent VT is high and the risk of VT in their first degree relatives is high whether an inherited thrombophilia is detected, suggest that genetic factors may be underpinning VT recurrence in these patients^20–24^. However, among the aforementioned prediction scores, only L-TRRiP includes genetic predictors, the FV Leiden and non-O blood groups, both well known to associate with first VT^12^. Unfortunately, knowledge of the genetic factors for VT recurrence is currently scarce. Indeed, while over 100 genetic loci associated for first VT have been identified^25,26^, only few studies have thoroughly investigated the genetic factors of VT recurrence. To date, only one genome wide association study (GWAS) has been performed on VT recurrence (totalling 1,279 patients including 447 VT recurrences)^27^. This GWAS identified the *F5* locus as the only genome-wide significant (P<5×10^-8^) locus. Genotype analysis in replication cohorts suggested the existence of another susceptibility locus in the 18q22 region. However, in the combined meta-analysis, this association (P=5.8×10^-6^) did not reach genome-wide significance. Finally, it is worth noting that the *ABO* locus, and more precisely A1 and A2 haplotypes, has recently been observed to be associated with VT recurrence (P=4.2×10^-3^)^28^.

We sought to identify new susceptibility loci for VT recurrence in the largest GWAS meta-analysis encompassing 6,571 patients, including 1,816 VT recurrences. We conducted downstream analyses to investigate the identified loci, followed by the same strategy stratified by sex, provoked/unprovoked status of the initial VT and the type of first VT (*i.e.*, PE/DVT).

## Methods

### Studies

The recurrent VT GWAS meta-analysis included eight studies with confirmed unrelated VT cases of European ancestry free of cancer history: EDITH, FHS, HVH (HVH1 and HVH23), MARTHA, MEGA, PADIS-PE and REVERSE-I (**Supplementary Material**).

### GWAS analysis and meta-analysis

In each study, all variants with a minor allele frequency (MAF) higher than 0.01 and an imputation quality score (INFO) higher than 0.3 were considered. Each study implemented a Cox model to analyse VT recurrence considering the delay since the first VT as time scale and adjusting for age at the first VT, sex, the presence of provoking factors and location of the first VT (if relevant) and the first principal components of the population stratification. Detailed information on genotyping arrays is provided in **Supplementary Table S1**.

In the primary analysis, only SNPs present in at least seven out of the eight participating cohorts were included in the meta-analysis conducted with the GWAMA software, using a fixed-effects inverse-variance weighted (IVW) model^29^.

### Testing of genetic variants associated with first VT

In a secondary analysis, we assessed whether the 100 genetic loci associated at P<5×10^-8^ with first VT in the European ancestry meta-analysis were also associated with recurrent VT in the current GWAS meta-analysis^26^.

### Transcriptome-wide association studies

From GWAS meta-analysis of VT recurrence, transcriptome-wide association study (TWAS) complemented by conditional and colocalization analyses, were performed with FUSION pipeline^30,31^. Tissues considered are in **Supplementary Table S2**. Associations were considered significant if they reached the corrected threshold corresponding to the average number of genes tested in each tissue (N=6,528, P<7.7×10^-6^). Results with a posterior probability of sharing a single causal variant (PP4) from the colocalization analysis higher than 0.75 were considered^31^.

### Mendelian Randomization with haemostatic phenotypes

From GWAS summary statistics of VT recurrence, we performed Mendelian randomization (MR) analyses with 29 haemostatic traits (**Supplementary Table S3**) using publicly available summary statistics from GWAS catalog (https://www.ebi.ac.uk/gwas/downloads/summary-statistics) and resources from the CHARGE hemostasis working group.

For each trait, we identified *cis* and *trans* independent genetic instruments (P<5×10^-8^) after clumping for linkage disequilibrium (LD) at r^2^<0.01 for a distance of 10Mb (based on European 1000 Genomes phase 3 reference panel). For phenotypes with less than two instruments, the selection threshold was lowered to P<1×10^-6^. We applied the recommended IVW MR method and to assess robustness of the findings, we further applied alternative MR methods that are more robust for the presence of pleiotropic or outlier instruments (Weighted Median method, MR-Egger)^32^. MR analyses were performed in R v.4.1.0 using the TwoSampleMR R package^33^.

### Mendelian Randomization with proteins

We performed a proteome MR analysis using publicly available GWAS results on 4,907 and 4,979 blood proteins measured with the Somalogic platform from the deCODE project (N=35,559) and the Fenland study (N=10,708)^34,35^; and 2,940 proteins measured with the antibody-based Olink Explore 3072 PEA in UK Biobank study (N=34,557)^36^. In each proteogenomics resource, we selected proteins influenced by more than two independent *cis* or *trans* genetic instruments (P<5×10^-8^). This led to the selection of 4,677 unique proteins present in deCODE (N=3,469), Fenland (N=2,706) or UK Biobank (N=2,217). We applied IVW method on each protein as well as the different aforementioned MR methodologies for sensitivity analyses. To account for multiple testing, we used a proteome-wide significance level of P<1.07×10^-5^ (∼0.05/4677) with IVW method.

### Mendelian Randomization with metabolites

We used GWAS summary statistics from 1,091 blood metabolites and 309 metabolite ratios to perform MR on VT recurrence with a similar approach as described above^37^. We identified 459 metabolites with more than two genetic instruments (P<5×10^-8^) which led to a statistically significant threshold of P<1.09×10^-4^ (∼0.05/459).

### Subgroup analyses

To better characterize heterogeneity of VT recurrence, a similar framework was applied to six subgroups: male/female sex; first VT provoked/first VT unprovoked; DVT only as first VT/PE±DVT as first VT. To account for multiple testing, additional correction for six phenotypes was applied for GWAS meta-analyses and downstream analyses. Because of lower sample sizes, we retained only variants with low heterogeneity across studies (I2<50%) and present in at least N-1 studies, with N the number of studies contributing to the dedicated subgroup analysis.

## Results

### Population characteristics

A brief description of the eight participating cohorts totalling 6,571 patients among which 1,816 experienced VT recurrences is presented in **Table 1**. Some differences were observed: *e.g.*, DVT represented approximately 80% of first VT in MARTHA whereas PADIS-PE was fully composed of PE patients; first VT was unprovoked in 32% of MEGA patients and was an inclusion criterion in PADIS-PE and REVERSE-I. A detailed description according to the incidence of VT recurrence is presented in **Supplementary Table S4**.

**Table 1:**
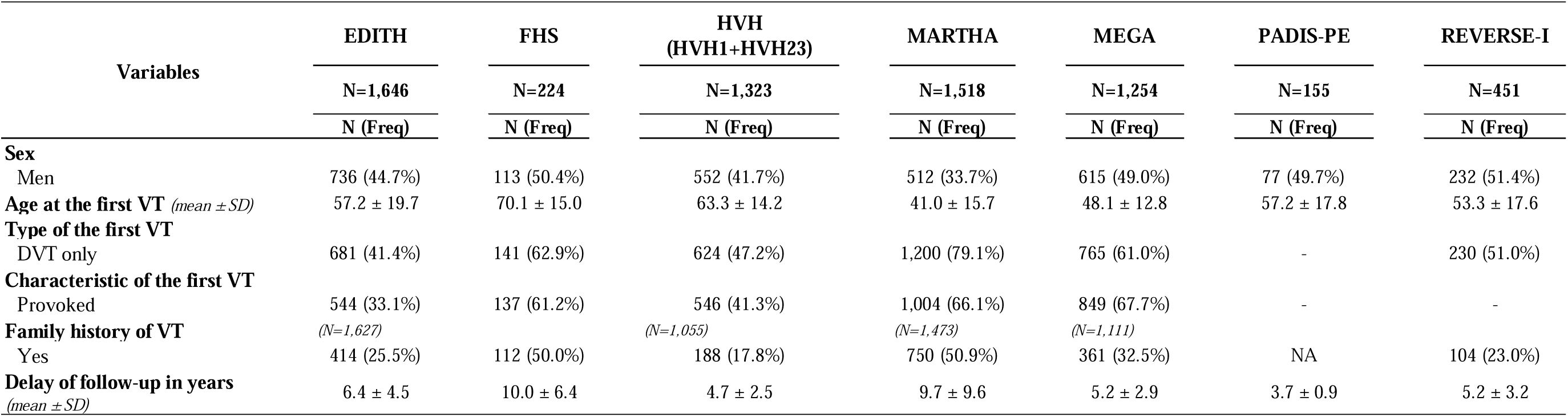
Description of the main population characteristics of the studies part of the meta-analysis.

### GWAS meta-analysis of VT recurrence

In the GWAS meta-analysis, 8,194,239 autosomal SNPs were tested with VT recurrence. The genomic inflation factor (lambda) was 1.04 and the associated Manhattan plot is presented in **Figure 1**. Three independent genetic loci were significantly (P<5×10^-8^) associated with VT recurrence (**Table 2, Supplementary Table S5**): -rs34097149 variant mapping to the *GPR149/MME* locus on 3q25.2 (**Figure 2A**) where the C allele (2.4%) was associated with an increasing risk of VT recurrence (hazard ratio (HR)=1.84 [1.49-2.29], P=2.65×10^-8^). Of note, *GPR149* was previously associated with coronary artery disease and with haemostatic traits (FXI, FVII, Fibrinogen, vWF)^38,39^.

**Figure 1:**
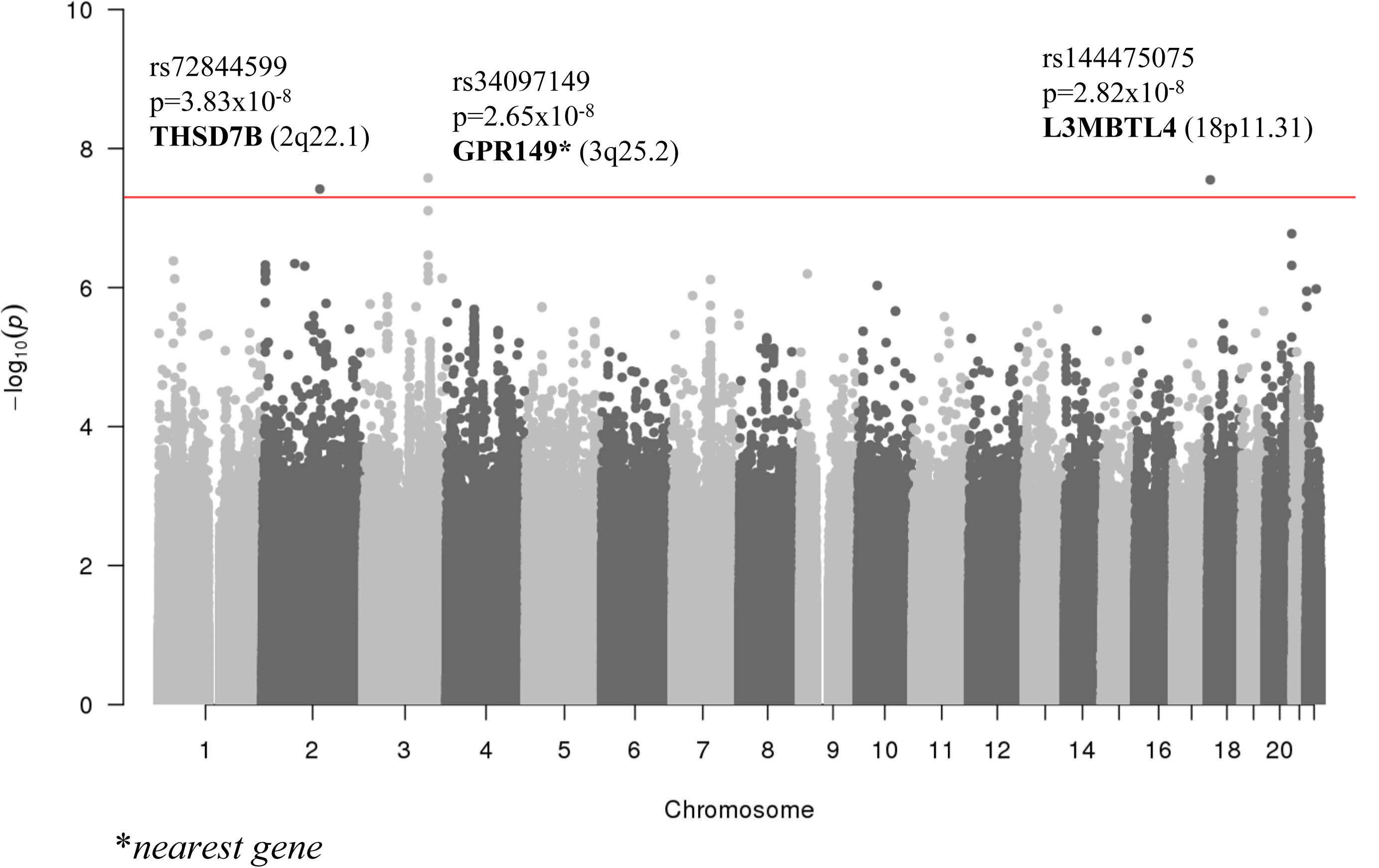
Manhattan plot representing GWAS results from the meta-analysis on VT recurrence. Horizontal red line represents the genome wide threshold (P<5×10^-8^). The three significant loci are annotated on this plot with their nearest gene.

**Figure 2:**
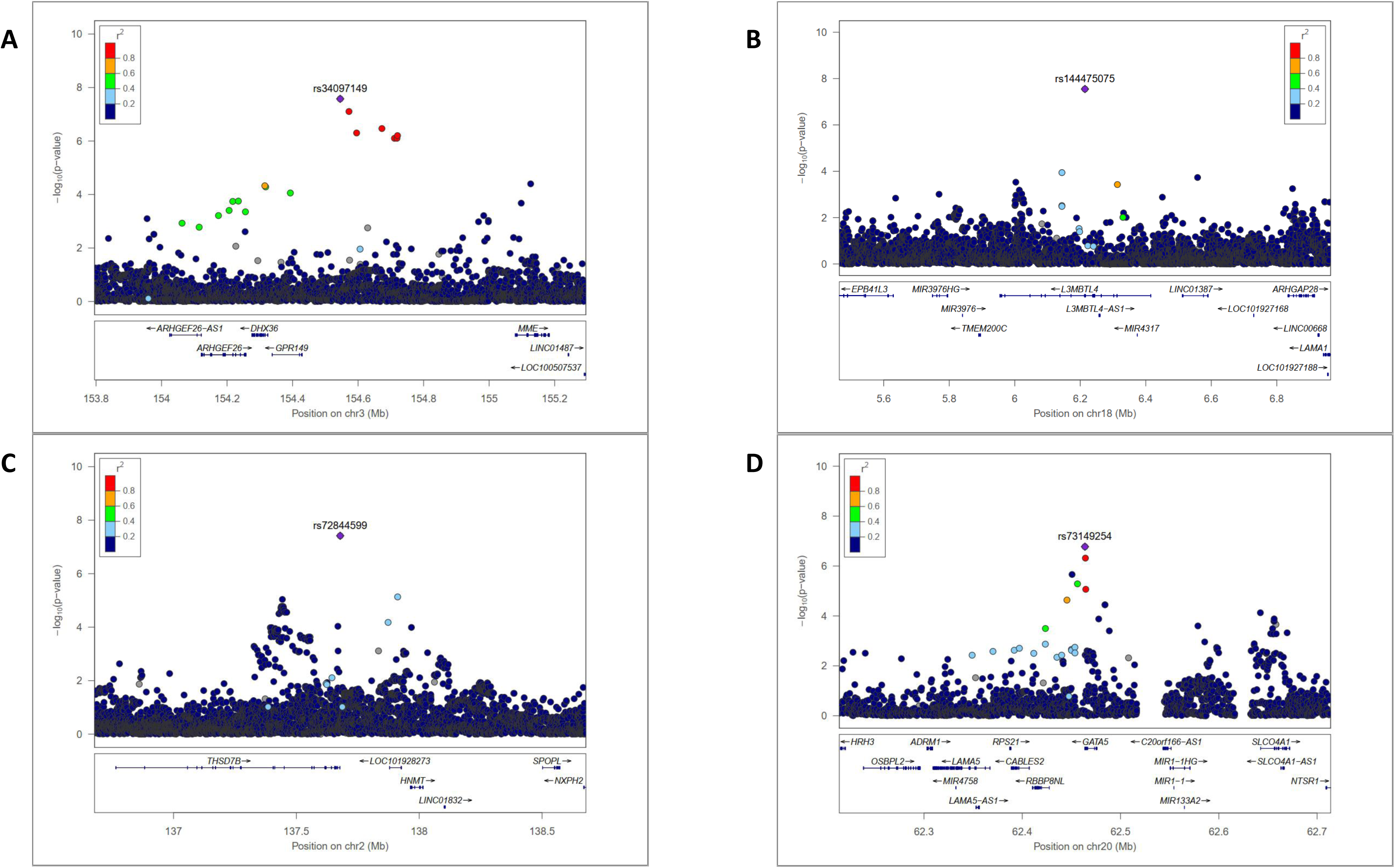
Regional association plot of the three significant loci and the suggestive locus identified in the main GWAS meta-analysis on VT recurrence. A) rs34097149 (3q25.2 *GPR149;MME*). B) rs144475075 (18p11.31 *L3MBTL4*). C) rs72844599 (2q22.1 *THSD7B*). D) rs73149254 (20q13.33 *GATA5*). This plot was generated with *locuszoom* software.

**Table 2:**
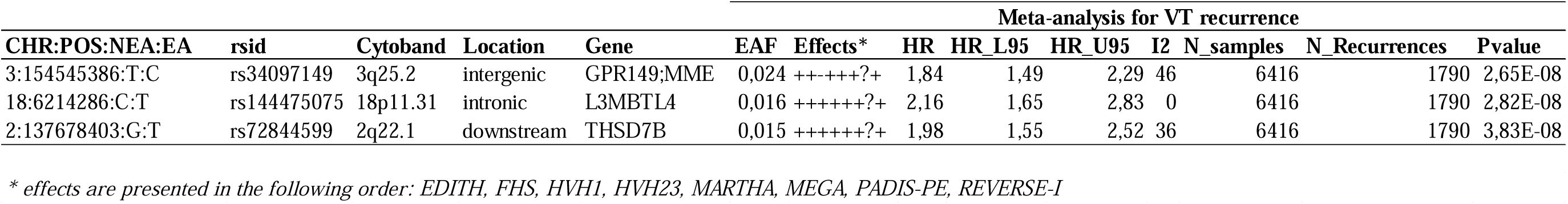
Genome wide significant loci associated with VT recurrence in the GWAS meta-analysis.

-rs144475075, intronic to *L3MBTL4* at the 18p11.31 locus where the T allele (1.6%) was associated with HR=2.16 [1.65-2.83] (P=2.82×10^-8^) (**Figure 2B**). *L3MBTL4* is a coding gene mainly expressed in vascular smooth muscle cells that may trigger vascular remodeling^40,41^. Little is known about its association with thrombotic disorders, except a potential link with hypertension^41^ and FVII activity^42^.

-rs72844599, downstream to *THSD7B* on chromosome 2q22.1 with T allele (1.4%) associated with HR=1.98 [1.55-2.52] (P=3.83×10^-8^) (**Figure 2C**). *THSD7B* belongs to the thrombospondin family, an inhibitor of angiogenesis that has been identified to associate with paediatric VT^43,44^. Of note, there was a trend for a stronger effect of this variant in younger patients (**Supplementary Table S6**). Thrombospondin is also known to be involved in platelet aggregation and has been proposed to interact with fibrinogen on the surface of activated platelets^45^.

For these three loci, the genetic effects were homogeneous across studies (**Supplementary** Figure 1A-C) as well as between subgroups (**Supplementary** Figure 2A-C). None of these lead SNPs were reported in GTEx portal as influencing gene expressions. They were however reported in the JASPAR database^46^ to be located in binding domains of a few transcription factors: *CDX1*, *CDX2*, *CDX4*, *HOXA10*, *HOXB13*, and *HOXD9* for rs34097149, *STAT4* for rs144475075 and *BCL6* for rs72844599.

Noteworthy, only the rs34097149 was reported to possibly act as a plasma protein quantitative trait locus (pQTL) (for *PDCD1LG2* and *SORBS3* at P<10^-4^) in Fenland (**Supplementary Table S7**).

In addition to these three significant loci, there was a variant that nearly reached (P<5×10^-7^) genome wide significance: rs73149254 in the 3’UTR of *GATA5* (HR=1.75 [1.42-2.16], P=1.68×10^-7^) (**Supplementary Table S8**, **Figure 2D, Supplementary Figure 1D-2D**). *GATA5* is a transcription factor that influences angiogenesis, endothelial cells function, platelets production and megakaryocyte development^47,48^. Rs73149254 influenced plasma levels of *ROBO2* in UK Biobank (**Supplementary Table S7**) and it was predicted to map to a binding site for hsa-miR-4737 ^49^ as well as for transcription factors *ZNF257* and *ZKSCAN5* ^46^.

### Effects of the variants associated with first VT on VT recurrence

Due to low frequency or imputation quality, only 88 first VT-associated SNPs^26^, could be tested for association with VT recurrence (**Supplementary Table S9**). Two associations passed the corrected threshold (P<5.7×10^-4^), *i.e.,* rs2066864-A in the 3’UTR of *FGG* (HR=1.14, P=2.3×10^-4^) and the *KNG1* exonic rs710446-C (HR=1.13, P=5.0×10^-4^). For these two variants, the observed genetic effects were in the same direction as that observed for first VT (**Table 3**).

**Table 3:** Nominal effects on VT recurrence of SNPs previously identified to associate with first VT.

|  |  |  |  |  | European meta-analysis for VT<br>(Thibord et al. 2022) |  |  |  |  | Meta-analysis for VT recurrence |  |  |  |  |  |  |  |
| --- | --- | --- | --- | --- | --- | --- | --- | --- | --- | --- | --- | --- | --- | --- | --- | --- | --- |
| CHR:POS:NEA:EA | rsid | Cytoband | Location | Gene | EAF | OR | OR_L95 | OR_U95 | I2 | Pvalue | EAF | HR | HR_L95 | HR_U95 | I2 | N_samples | Pvalue |
| 4:154604543:G:A | rs2066864 | 4q32.1 | UTR3 | FGG | 0,25 | 1,23 | 1,22 | 1,25 | 53 | 8,63E-218 | 0,29 | 1,14 | 1,07 | 1,23 | 57 | 6571 | 2,32E-04 |
| 3:186742138:T:C | rs710446 | 3q27.3 | exonic | KNG1 | 0,41 | 1,05 | 1,03 | 1,06 | 11 | 2,12E-14 | 0,43 | 1,13 | 1,05 | 1,21 | 38 | 6571 | 4,96E-04 |
| 10:94263320:T:C | rs57866767 | 10q23.33 | intronic | PLCE1 | 0,43 | 1,04 | 1,03 | 1,05 | 0 | 4,64E-11 | 0,46 | 1,12 | 1,05 | 1,19 | 10 | 6571 | 9,36E-04 |
| 3:90388838:A:G | rs9866664 | 3p11.1 | intergenic | EPHA3;NONE | 0,49 | 0,95 | 0,94 | 0,97 | 0 | 1,38E-13 | 0,45 | 0,88 | 0,81 | 0,95 | 50 | 6571 | 1,68E-03 |
| 9:133261703:G:A | rs687289 | 9q34.2 | intronic | ABO | 0,36 | 1,34 | 1,32 | 1,35 | 86 | 7.25e-507 | 0,44 | 1,11 | 1,04 | 1,19 | 0 | 6571 | 2,19E-03 |
| 1:109272258:C:T | rs4970834 | 1p13.3 | intronic | CELSR2 | 0,19 | 1,05 | 1,03 | 1,06 | 4 | 6,92E-10 | 0,19 | 0,88 | 0,80 | 0,95 | 20 | 6571 | 2,51E-03 |
| 11:46739505:G:A | rs1799963 | 11p11.2 | UTR3 | F2 | 0,01 | 1,99 | 1,90 | 2,09 | 74 | 8,44E-170 | 0,03 | 1,43 | 1,12 | 1,81 | 19 | 5317 | 3,40E-03 |
| 17:2075894:T:C | rs4790311 | 17p13.3 | intronic | SMG6 | 0,60 | 1,05 | 1,04 | 1,06 | 0 | 3,19E-15 | 0,62 | 0,91 | 0,85 | 0,98 | 0 | 6571 | 8,69E-03 |
| 4:186285095:C:A | rs3756011 | 4q35.2 | intronic | F11 | 0,40 | 1,22 | 1,21 | 1,23 | 66 | 1,15E-238 | 0,47 | 1,09 | 1,02 | 1,17 | 5 | 6571 | 1,31E-02 |
| 12:54336088:T:C | rs4759076 | 12q13.13 | intronic | COPZ1 | 0,47 | 0,95 | 0,94 | 0,96 | 0 | 6,35E-17 | 0,42 | 0,92 | 0,85 | 0,98 | 0 | 6571 | 1,64E-02 |
| 8:105569300:A:T | rs6993770 | 8q23.1 | intronic | ZFPM2 | 0,28 | 0,93 | 0,92 | 0,94 | 0 | 4,61E-29 | 0,27 | 0,91 | 0,84 | 0,99 | 0 | 6571 | 2,11E-02 |
| 12:8901065:T:C | rs7311483 | 12p13.31 | intergenic | A2ML1;PHC1 | 0,68 | 1,04 | 1,02 | 1,05 | 0 | 3,63E-08 | 0,65 | 1,08 | 1,01 | 1,16 | 31 | 6571 | 2,61E-02 |
| 1:207108804:C:A | rs2842700 | 1q32.2 | intronic | C4BPA | 0,06 | 1,12 | 1,10 | 1,15 | 26 | 2,50E-19 | 0,07 | 1,14 | 1,01 | 1,29 | 0 | 6571 | 3,49E-02 |
| 10:119250744:A:G | rs10886430 | 10q26.11 | intronic | GRK5 | 0,12 | 1,12 | 1,10 | 1,14 | 0 | 3,59E-34 | 0,14 | 1,10 | 1,01 | 1,21 | 0 | 6571 | 3,64E-02 |
| 1:169549811:T:C | rs6025 | 1q24.2 | exonic | F5 | 0,97 | 0,34 | 0,33 | 0,35 | 84 | 1.57e-1028 | 0,92 | 0,87 | 0,77 | 0,99 | 74 | 6571 | 3,83E-02 |
| 19:10628160:A:G | rs8109681 | 19p13.2 | intronic | SLC44A2 | 0,79 | 1,12 | 1,10 | 1,14 | 14 | 9,19E-55 | 0,78 | 1,10 | 1,00 | 1,20 | 10 | 6571 | 4,47E-02 |
| 22:42719770:G:C | rs9611844 | 22q13.2 | intronic | A4GALT | 0,12 | 1,08 | 1,06 | 1,10 | 48 | 7,94E-17 | 0,13 | 1,10 | 1,00 | 1,21 | 0 | 6571 | 4,54E-02 |
| 12:6051448:C:T | rs7135039 | 12p13.31 | intronic | VWF | 0,36 | 1,07 | 1,06 | 1,09 | 11 | 1,20E-31 | 0,39 | 1,07 | 1,00 | 1,15 | 45 | 6571 | 4,76E-02 |

### TWAS on VT recurrence

The main results (P<1×10^-5^) of the TWAS on VT recurrence are provided in **Supplementary Table S11**. No new loci were identified using the pre-specified statistical threshold of P<7.7×10^-6^.

### Mendelian randomization with haemostatic phenotypes

Results of MR on 29 haemostatic phenotypes are presented in **Table 4**. At the Bonferroni corrected threshold for multiple testing (P<1.7×10^-3^), only higher levels of FXI were significantly associated with a greater risk of VT recurrence (HR=1.21 [1.09-1.35], P=4.76×10^-4^).

**Table 4:**
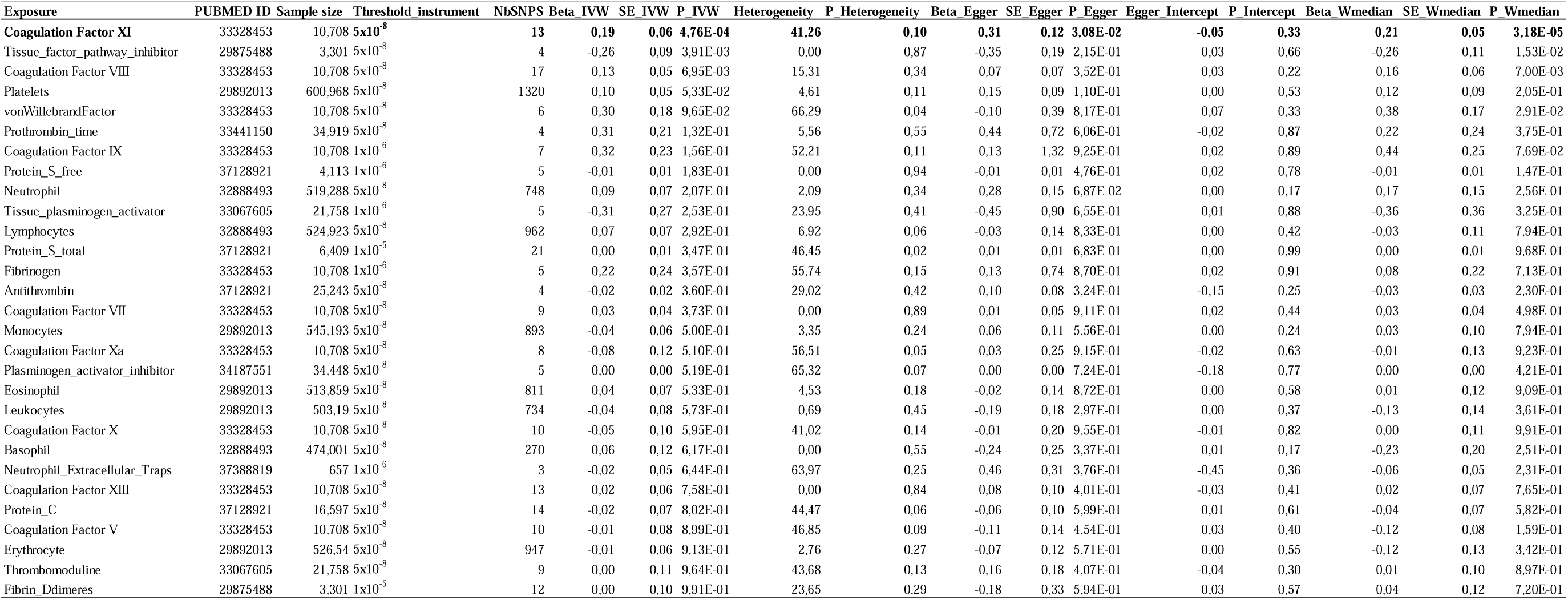
Results of MR analyses with 29 haemostatic phenotypes on the risk of VT recurrence.

### Protein QTL Mendelian Randomization on VT recurrence

MR analysis on 4,677 human plasma circulating proteins (all results in **Supplementary Table S11**) identified three significant (P<1.07×10^-5^) associations (**Table 5**) which were consistent across the three used MR methods (**Supplementary Figure S3**). Effects of the genetic instruments for the three proteins identified are presented in **Supplementary Table S12**.

**Table 5:**
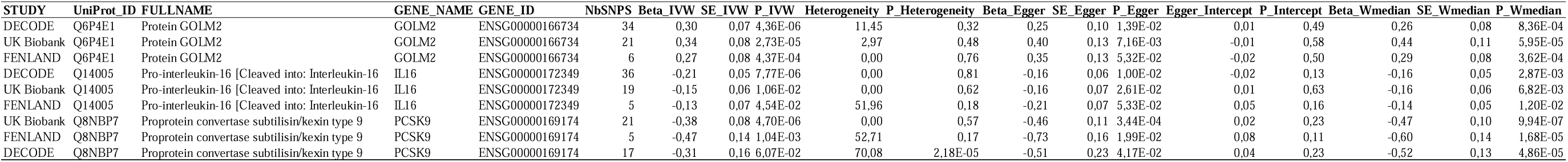
Significant results of pQTL MR on VT recurrence.

From deCODE, MR results showed that one unit increase of genetically-determined levels of protein GOLM2 was significantly associated with an increasing risk of VT recurrence (HR=1.35 [1.18-1.55], P=4.36×10^-6^). In both Fenland and UK Biobank, the *ABO* rs550057-C was the strongest genetic instrument for GOLM2 while it was rs687289-A in deCODE. Interestingly, rs550057-C tagged *ABO* A1 and A2 haplotypes while rs687289-A tagged non-O *ABO* haplotypes (**Supplementary Figure S4**). Using raw proteogenomic data from the 3C-study (**Supplementary Materials**)^50^ we confirmed that GOLM2 plasma levels were increased in A1 carriers (**Supplementary Table S13**). Associations of ABO blood group tagging SNPs with VT recurrence are shown in **Supplementary Table S14**.

Secondly, we observed that one unit increase of genetically-determined plasma levels of pro-Interleukin-16 (*IL16*) was significantly associated with a decreased risk of VT recurrence (HR=0.81 [0.78-0.95], P=7.77×10^-6^) in deCODE. Similar trends were observed in UKBiobank (HR=0.86, P=0.011) and Fenland studies (HR=0.88, P=0.045). The strongest genetic instrument for pro-IL16 in deCODE (**Supplementary Table S12**) was the intronic *IL16* rs17875523 variant, which was in complete LD (r^2^=1 according to HaploReg v4.2)^51^ with the missense rs11556218 (p.Asn1147Lys). The rs11556218-G allele was associated with decreased pro-IL16 levels and slightly with increased risk of VT recurrence (HR=1.18, P=4.5×10^-3^). Of note, MR association observed in deCODE remained unchanged after removing rs11556218 (or any SNP in strong LD) from the genetic instruments (HR=0.70, P=8.9×10^-5^).

Finally, we observed that increase of genetically-determined plasma levels of proprotein convertase subtilisin/kexin type 9 (*PCSK9*) was significantly associated with a lower risk of VT recurrence (HR=0.68 [0.58-0.80], P=4.70×10^-6^) from UK Biobank. Similar associations were observed in deCODE (HR=0.73, P=0.06) and Fenland (HR=0.63, P=1.04×10^-3^). The strongest genetic instrument of PCSK9 in UK Biobank was the missense *PCSK9* rs11591147-T (p.Arg46Leu) variant associated with decreased PCSK9 levels (β=-1.06, P<1×10^-200^) and increasing risk of VT recurrence (HR=1.76, P=4.28×10^-6^). After removing this variant from the MR analysis, no association remained (UK Biobank: HR=0.81, P=0.11; Fenland: HR=0.89, P=0.55; deCODE: HR=0.96, P=0.86).

### Mendelian randomization with metabolites on VT recurrence

Results of MR with 459 metabolites on the risk of VT recurrence are presented in **Supplementary Table S15**. No significant association was identified at the predefined threshold of P<1.09×10^-4^.

### Subgroup analysis

The same GWAS workflow, together with downstream analyses, were deployed in specific subgroups of VT patients. All results are provided in **Supplementary Table S16-20 and Supplementary Figures S5-22**.

By conducting subgroup specific GWAS meta-analyses, we identified 25 SNPs, mapping to 18 independent loci, which reached the predefined statistical threshold of P<8.3×10^-9^ (**Supplementary Table S16**). All the identified SNPs were low-frequency variants (about 2%) and showed concordant directions of effect across the contributing studies. Among these results, the most significant finding was the *SLC4A1* rs45562031-T missense variant (p.Glu40Lys) identified in the PE subgroup (HR=3.23 [2.30-4.52], P=9.7×10^-12^). Interestingly, this variant had no effect (HR=1.00, P=0.98) in the DVT subgroup (**Supplementary Figure S4B**).

TWAS results that passed the corrected threshold used for the main analysis (P<7.7×10^-6^) are presented in **Supplementary Table S17**. After correction, only two associations remained significant (P<1.3×10^-6^) but there was no evidence of colocalization.

For pQTL-MR, MR analyses with metabolites and haemostatics phenotypes, no significant association remained after correction for multiple phenotypes (P<1.78×10^-6^, P<1.82×10^-5^ and P<2.87×10^-4^, respectively). Results are presented in **Supplementary Table S18** for MR with haemostatics phenotypes and nominal associations in **Supplementary Table S19-20** for pQTL-MR and metabolites MR, respectively.

## Discussion

The current study, representing the largest effort to identify molecular risk factors for VT recurrence, identified 29 significant molecular markers (summarized in **Figure 3**). Consistent with the hypothesis that VT recurrence is a complex trait, we observed that 9 of these markers (*FGG*, *KNG1*, *GPR149*, *L3MBTL4* and *THSD7B* loci; GOLM2, FXI, IL16 and PCSK9 plasma levels) pertained to VT recurrence in the general populations of VT patients while the remaining 20 were more specific to subgroups of VT patients based on sex, type of first event (PE/DVT), or provoked/unprovoked status of the first VT. Furthermore, we demonstrated that the genetics of VT recurrence differed from that of the first VT as only three markers (*FGG*, *KNG1* loci and genetically-determined FXI plasma levels) were shared. Sensitivity analyses addressing the possible impact of the collider bias phenomenon due to case-only design did not modify these findings (**Supplementary Note**).

**Figure 3:**
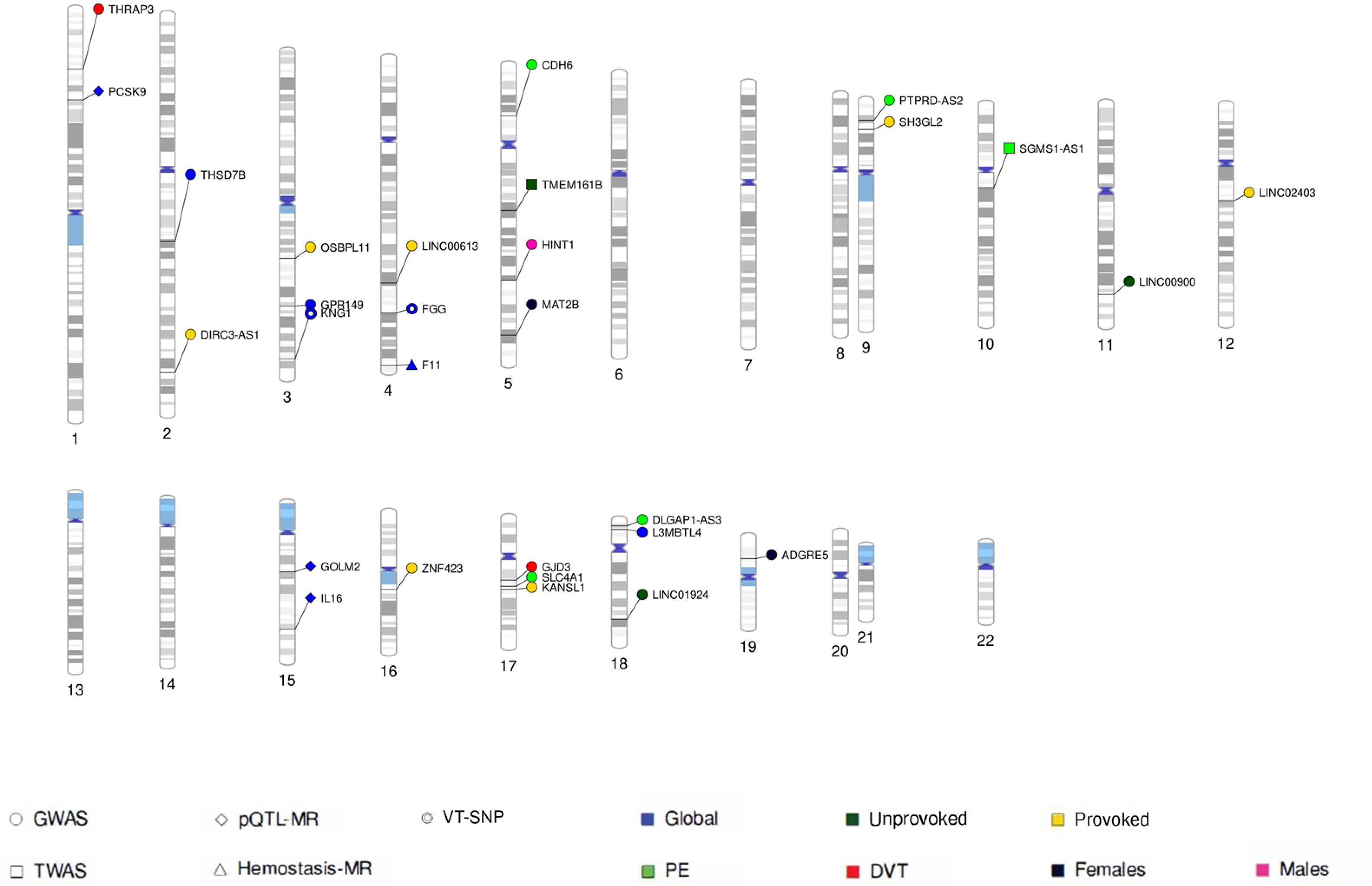
Representation of all loci identified to significantly associate with VT recurrence. Shapes are specific to the analysis: round for GWAS, diamond for pQTL-MR, circle for look-up of first VT SNPs, square for TWAS and triangle for MR on haemostasis phenotypes. Different colours are used to differentiate the groups: blue for the global analysis, dark green for unprovoked, yellow for provoked, light green for PE, red for DVT, dark for females and pink for males. This plot was generated with *PhenoGram* web tool (http://visualization.ritchielab.org/).

MR analyses supported the causal association between the plasma levels of 4 proteins (GOLM2, FXI, IL16 and PCSK9) and the risk of recurrence. Increasing genetically-determined FXI levels associated with increased risk of recurrence, which was in line with the use of FXI inhibitors to prevent VT recurrence^52^ and consistent with previously reported results^15^. In the same study, lower plasma levels of free TFPI^16^ and high plasma levels of FVIII^17^ were associated with a higher risk of VT recurrence. Results of our MR analyses showed consistent suggestive evidence for both haemostatic traits on the risk of VT recurrence, HR=0.77 (P=3.91×10^-3^) and HR=1.14 (P=6.95×10^-3^) for TFPI and FVIII plasma levels, respectively (**Table 4**). Of note, the association between FXI and recurrent VT remained unchanged when excluding from the MR analysis the *KNG1* rs710446 variant (HR=1.18, P=0.03), which is known to influence plasma FXI levels^53^.

By contrast, MR results observed for GOLM2, IL16 and PCSK9 were novel.

The GOLM2 protein is involved in cellular processes related to cancer development and progression. While its precise functions are not fully understood, GOLM2 is believed to be involved in cell proliferation, apoptosis, contributing to tumorigenesis when dysregulated^54^. The association between GOLM2 and recurrent VT was consistent across subgroups, including provoked and unprovoked VT patients (**Supplementary Table S19**), suggesting that the biological impact of GOLM2 extends beyond cancer related mechanisms. The most consistent pQTLs for GOLM2 were located in the *ABO* (9q34.2), *SIK3* (11q23.3) and *POC1B* (12q21.33) loci, among which *ABO* was the most strongly associated with VT recurrence (**Supplementary Table S21**). However, ABO blood groups only contributed to 2.5% of GOLM2 plasma levels variability in the 3C study.

We also identified that higher levels of pro-IL16 were protective against VT recurrence. Interestingly, the missense *IL16* rs11556218-G variant has recently been reported to associate with VT in leukemia patients^55^. However, no trend for association was observed in the GWAS on first VT (OR=1.01, P=0.56)^26^. Pro-IL16 has been described to act as a transcriptional repressor in lymphocytes cells^56^ which regulates cell proliferation^57^. While high levels of the mature form of pro-IL16 are associated with a poorer cancer prognosis^56^, there is growing evidence supporting a protective role of pro-IL16 in cardiovascular diseases. Indeed, several studies demonstrated a protective role of IL16 in atherosclerotic plaque stabilization and a reduced incidence of cardiovascular events^58–60^. By facilitating plaque stabilization, pro-IL16 may attenuate tissue factor activation, fibrin formation and platelet recruitment^61^ thereby contributing to protection against VT recurrence.

The third protein identified was PCSK9, a protein well known for its role in lipid metabolism by its ability to enhance the degradation of low-density lipoprotein (LDL) receptors^62^. However, our results identifying a lower VT recurrence risk with increased PCSK9 levels were counterintuitive and seemed to contradict previous studies showing that PCSK9 inhibitors, prescribed to lower LDL cholesterol, were associated with a reduced risk of cardiovascular diseases, including the risk of VT^63–65^. Given that the association with PCSK9 levels disappeared after removing the effect of the missense variant *PCSK9* p.Arg46Leu, it is possible that this variant qualitatively impacts the ability of the proteomic technique used to measure PCSK9 plasma levels. However, this could not explain why we observed a deleterious effect of the Leu46 variant on the risk of VT recurrence, while it has a protective effect on coronary artery disease^66,67^. Conversely, this variant did not show any association with VT incidence^26^ nor with stroke^68^. MR analysis with LDL cholesterol levels^69^ on VT recurrence did not support a causal association (HR=0.97, P=0.59). This suggests that the PCSK9 association identified may reflect mechanisms outside the LDL-pathway. In line with this hypothesis is the observation that statin therapy, which has been proposed to prevent VT recurrence^70–72^, can also increase PCSK9 levels^65^. Of note, in the EDITH study, the *PCSK9* Leu46 variant tended to associate with VT recurrence more strongly in patients under statins therapy (HR=14.40, P=2.0×10^-3^) than in the group of non-statins users (HR=1.45, P=0.13) (**Supplementary Table S22**). Altogether, these findings suggest either the presence of an unmeasured confounder and/or more complex interactions between the underlying mechanisms. Further works are needed to better clarify the association of PCSK9 and the risk of VT recurrence.

Beyond these MR findings, we identified 3 novel loci (*GPR149*, *L3MBTL4* and *THSD7B*) harbouring SNPs associated with VT recurrence in the general population of VT patients. None of the lead SNPs demonstrated strong statistical association with gene expression or plasma protein levels, highlighting the need for further characterization of their functional impact. In our meta-GWAS analysis, we did not replicate the previously reported association of the rs9946608-C (HR=1.13, P=0.28, I^2^=21%, **Supplementary Figure S24**) and observed a strong heterogeneity in the association of FV Leiden with VT recurrence (HR=1.14, P=0.04, I^2^=74%, **Supplementary Figure S23**).

In subgroups analyses, attention was drawn to the missense *SLC4A1* p.Glu40Lys (rs45562031) variant associated with a three-fold increased risk of recurrence in PE patients (**Supplementary Figure S10**). Interestingly, a rare missense variant in exon 6 of *SLC4A1* (p.Gly130Arg, rs121912749) was identified in an Asian pedigree with unexplained venous and arterial thrombi^73^. The band 3 protein encoded by *SLC4A1* is a chloride/bicarbonate exchanger lying in the red blood cell (RBC) membrane and involved in carbon dioxide transport from tissues to lungs. It is also found in the kidney, where it is involved in acid secretion. Rare coding mutations in *SLC4A1* are responsible for new blood group antigens belonging to the Diego blood group system. These mutations can also alter the RBC membrane and kidney functions. Whether the *SLC4A1* p.Glu40Lys variant has similar functional impacts on RBC or on kidney remains to be elucidated. The broad biological implications of the band 3 protein in functions of the respiratory system^74^ may explain why this gene was identified only in the PE subgroup. Of note, no criteria on the localisation was applied to qualify a recurrent VT, as in approximately 80% of cases, the clinical manifestation of a VT recurrence is the same as the initial VT presentation^75^.

Despite being the largest GWAS on VT recurrence, the moderate size of our study may have hampered our power to detect additional significant findings. Some borderline findings would require further investigations such as the association of the *GATA5* 3’UTR rs73149254 or the association of the *PLXNA4* locus with VT recurrence in PE patients (**Supplementary Table S16**), a locus identified with the risk of PE in a plasma proteomic study^76^.

By bringing together almost all known existing cohorts with data on recurrent VT, we were unable to identify a substantial sample to replicate the associations identified. To support our findings and go further to decipher the mechanisms involved, we have extensively leveraged publicly available genomic data.

Another limitation of this work is its focus exclusively on populations of European ancestry. It is now widely acknowledged that incorporating cross-ancestry populations can improve the power of GWAS to identify genetic loci. However, it is important to note that the MAF of the identified variants is often lower in non-European ancestry populations (**Supplementary Table S23**), which will substantially reduce the power to replicate these findings in other ancestral groups.

In conclusion, we described 25 loci and 4 proteins significantly associated with VT recurrence. Some of these associations pertained to the risk of recurrence in the general population of VT patients of European ancestry and some were restricted to specific subgroups. Our findings provided novel insights into the genomic architecture of VT recurrence, and highlighted potential targets for developing or repositioning drugs. The newly identified markers now need to be integrated into prediction tools to evaluate their clinical relevance for more personalized anticoagulation therapy.

## Statements and Declarations

## Data Availability

Summary statistics from the main analysis will be uploaded on GWAS catalog.

## Acknowledgments

Statistical analyses benefited from the technical support of the CBiB computing centre of the University of Bordeaux.

The views expressed in this article are those of the authors and do not necessarily represent the views of the National Institutes of Health or the US Department of Health and Human Services.

The authors thank Lee Vernich for her diligent support coordinating the array analyses of the REVERSE-I study samples, Elham Sabri for sharing her deep knowledge of the REVERSE-I data, and Liz Li of The Centre for Applied Genomics, The Hospital for Sick Children, Toronto, Canada, for assistance in the quality control of the genotyping data from REVERSE-I.

## Fundings

GM and D-AT are supported by the EPIDEMIOM-VT Senior Chair from the University of Bordeaux initiative of excellence IdEX. IC is supported by the EUR DPH, a PhD program supported within the framework of the PIA3 (Investment for the future), project reference 17-EURE-0019.

EDITH, MARTHA, MEGA and PADIS-PE genomics research programs were supported by the GENMED Laboratory of Excellence on Medical Genomics [ANR-10-LABX-0013], a research program managed by the National Research Agency (ANR) as part of the French Investment for the Future.

The MARTHA project was supported by a grant from the Program Hospitalier de la Recherche Clinique The 3C proteomics project was supported by a grant overseen by the French National Research Agency (ANR) as part of the “Investment for the Future Programme” ANR-18-RHUS-0002 and by the Precision and Global Vascular Brain Health Institute (VBHI) funded by the France 2030 IHU3 initiative.

This project was conducted in the framework of the MORPHEUS project funded by the HORIZON-HLTH-2022-TOOL-11-01 program.

The REVERSE I study genetic analysis was supported by a grant of the Canadian Institutes of Health Research (grant PJT-162233) and a fellowship from the CANSSI Ontario STAGE (Genetic Epidemiology and Statistical Genetics Research training program). The computational analyses were performed at the SciNET HPC Consortium of the University of Toronto.

In France, the EDITH, MARTHA and PADIS-PE studies are part of the French Clinical Research Infrastructure Network on Venous Thrombo-Embolism (F-CRIN INNOVTE). In Canada participating centers are members of the CanVECTOR Network. These national networks are members of INVENT-VTE, the International Network of Venous Thromboembolism Clinical Research Networks (www.invent-VTE.com).

The MEGA (Multiple Environmental and Genetic Assessment of risk factors for venous thrombosis) study was supported by the Netherlands Heart Foundation (NHS98.113 and NHS208B086), the Dutch Cancer Foundation (RUL 99/1992), and the Netherlands Organization for Scientific Research (912– 03-033|2003).

The FHS (Framingham Heart Study) is a joint project of the NIH and Boston University School of Medicine and was supported by contract N01-HC-25195. This research was supported by NHLBI Intramural Research Funding. The computational work contributed to this paper for the FHS was performed on the Shared Computing Cluster which is administered by Boston University’s Research Computing Services. URL: www.bu.edu/tech/support/research/.

The HVH study was supported by National Heart, Lung, and Blood Institute (NHLBI) grants R01HL60739, R01HL73410, R01HL95080, and R01HL134894. L.B. Harrington’s time on this project was supported by NHLBI grants K01HL139997 and R01HL166292.

Work in the Danckwardt lab is supported by the German Research Foundation (SPP 1935, DA 1189/2-1, DA 1189/5-1, DA 1189/7-1, INST 371/33-1, GRK 2859), the Federal Ministry of Education and Research (BMBF01EO1003), the German Society of Clinical and Laboratory Medicine, the Hella Bühler Award for Cancer Research and the EU Horizon 2020 ITN ‘Thromboinflammation in Cardiovascular disorders’ (TICARDIO, Marie Skłodowska-Curie grant agreement No 813409).

## Authorship Contributions

GM, NLS, FC, PEM and DAT participated in study concept and design. GM, JAB, AvHV, PS, KLW, L.Goumidi, LBH, MAR, FG, FRR, ADJ, NLS, FC, PEM and DAT participated in phenotype data acquisition or control quality. MHC, AvHV, L.Gourhant, MG, IC, RO, NS, CB, DB, AB, S.Debette, JFD, FG, FRR, ADJ, NLS, FC, PEM and DAT participated in genotype or biological data acquisition or control quality. GM, MHC, OCB, JAB, FT, LBH, CAL, S.Danckwardt, HJG, ADJ, NLS, FC, PEM and DAT participated in data analysis and interpretation. GM and DAT wrote the initial draft of the manuscript which was reviewed and approved by all co-authors.

## Ethics approval

Research have been performed in accordance with the Declaration of Helsinki.

All experimental protocols to study the genetics of VT recurrence were approved by the local ethic committee “Mediterranean I Committee for the Protection of Individuals” (reference: 12 61) for MARTHA study.

The MEGA study was approved by the local ethic committee “Medical Ethics Committee of the Leiden University Medical Center”.

The HVH study was approved by the institutional review board of the Kaiser Permanente Washington Health Research Institute.

The EDITH study was approved by Brest University Hospital scientific and ethics board, in accordance with the Declaration of Helsinki.

The Institutional Review Board of Boston University Medical Center approved the study protocol for FHS study.

PADIS-PE study was conducted in accordance with the ethical principles stated in the Declaration of Helsinki, Good Clinical Practice, and relevant French regulations regarding ethics and data protection. The protocol and amendments were approved by a central independent ethics committee and written informed consent was obtained from all participants.

Protocols to study genetics in REVERSE I was approved by the University of Toronto Research Ethics Board. Institutional research ethics board approvals were obtained by all participating centers (Ottawa Hospital Research Ethics Board).

## Consent to participate

Written informed consent to participate was obtained from all participants.

## Data sharing statement

Summary statistics from the main analysis will be uploaded on GWAS catalog.

## SUPPLEMENTARY TABLE TITLES

**Supplementary Table S1:** Design and genetic/phenotypic details of studies that contributed to the meta-analysis on VT recurrence.

**Supplementary Table S2:** List of the tissues from GTEx v8 analyzed in TWAS.

**Supplementary Table S3:** Description of the 29 haemostatic phenotypes from GWAS catalog used to perform targeted MR with VT recurrence.

**Supplementary Table S4:** Description of the clinical characteristics of the patients from the studies part of the meta-analysis according to the incidence of VT recurrence.

**Supplementary Table S5:** Study specific effects of the three genome-wide significant loci associated with VT recurrence in the GWAS meta-analysis.

**Supplementary Table S6:** Effect of rs72844599 (*THSD7B*) on VT recurrence in younger patients in EDITH, MARTHA and MEGA.

**Supplementary Table S7:** Lead variants identified in GWAS for VT recurrence (in global and in subgroups) and their impact in proteogenomic resources.

**Supplementary Table S8:** Additional *GATA5* locus strongly associated with VT recurrence in the GWAS meta-analysis.

**Supplementary Table S9:** Association with VT recurrence of 88 SNPs previously identified to associate with first VT.

**Supplementary Table S10:** Main results of TWAS performed on the summary statistics of the GWAS meta-analysis on VT recurrence.

**Supplementary Table S11:** All results of pQTL MR with IVW method on VT recurrence

**Supplementary Table S12:** Instruments used for MR analyses on GOLM2, IL16 (from deCODE) and PCSK9 (from UK Biobank) and their association with first VT and VT recurrence.

**Supplementary Table S13:** Association of ABO haplotypes with GOLM2 plasma levels in the 3C-study (N=1,087).

**Supplementary Table S14:** Effect of ABO variants on VT recurrence in the GWAS meta-analysis (N=6,571).

**Supplementary Table S15:** Results of MR analyses with 459 metabolites from *Chen et al. (2022)* on the risk of VT recurrence.

**Supplementary Table S16:** Genome wide significant variant (P<5×10^-8^) associated with VT recurrence in the GWAS meta-analysis for the 6 subgroups.

**Supplementary Table S17:** Results of TWAS on VT recurrence in the 6 subgroups that passed the statistical threshold used for the main analysis (P<7.7×10^-6^).

**Supplementary Table S18:** Results of MR analyses with 29 haemostatic phenotypes on the risk of VT recurrence in 6 subgroups.

**Supplementary Table S19:** Nominal results of pQTL MR analyses with IVW method on the risk of VT recurrence in 6 subgroups.

**Supplementary Table S20:** Nominal associations of MR analyses with 459 metabolites on the risk of VT recurrence in the 6 subgroups.

**Supplementary Table S21:** Effect of GOLM2 pQTLs identified in the three proteogenomic resources on the risk of VT recurrence in the GWAS meta-analysis.

**Supplementary Table S22:** Effects of the missense variant PCSK9 p.Arg46Leu (rs11591147-T) on the risk of VT recurrence in EDITH patients with available information on treatment.

**Supplementary Table S23:** Effects of the variants identified with VT recurrence on the risk of first VT in *Thibord et al. (2022)* and their frequencies in ethnic groups.

## SUPPLEMENTARY FIGURE LEGENDS

**Supplementary Figure S1:** Study-specific effect of the 4 loci identified in the GWAS meta-analysis for VT recurrence. Figure A corresponds to rs34097149-C (*GPR149;MME*), B to rs144475075-T (*L3MBTL4*), C to rs72844599-T (*THSD7B*) and D to rs73149254-A (*GATA5*). N: Total sample size / NREC: Number of VT recurrences / EAF: Effect Allele Frequency / HR: Hazard Ratio for VT recurrence.

**Supplementary Figure S2:** Subgroup-specific effect of the 4 loci identified in the GWAS meta-analysis for VT recurrence. Figure A corresponds to rs34097149-C (*GPR149;MME*), B to rs144475075-T (*L3MBTL4*), C to rs72844599-T (*THSD7B*) and D to rs73149254-A (*GATA5*). N: Total sample size / NREC: Number of VT recurrences / EAF: Effect Allele Frequency / HR: Hazard Ratio for VT recurrence.

**Supplementary Figure S3:** Representation of the associations with VT recurrence in the three proteogenomics resources and according to the MR sensitivity methods, for the three proteins identified in the pQTL-MR analysis.

**Supplementary Figure S4:** Construction of haplotypes with LD Link tool^77^ (https://ldlink.nih.gov/) using the 5 variants identified by *Goumidi et al. (2021)*^78^ to tag ABO haplotypes and 2 pQTLs of GOLM2 located in the *ABO* locus (rs550057 by Fenland and UK Biobank ; rs687289 by deCODE).

**Supplementary Figure S5:** Associations of rs145765113-T identified in the female subgroup. A) Forest plot representing the hazard ratio for VT recurrence of the SNP-effects across the subgroups and in the main analysis. B) Forest plot representing the hazard ratio for VT recurrence of the SNP in the female subgroup for all studies that contributed to this subgroup analysis. C) Regional association plot around the SNP in the female subgroup analysis. N: sample size / NREC: number of VT recurrences / EAF: effect allele frequency / INFO: imputation quality score / HR: Hazard ratio.

**Supplementary Figure S6:** Associations of rs185486549-A identified in the female subgroup. A) Forest plot representing the hazard ratio for VT recurrence of the SNP-effects across the subgroups and in the main analysis. B) Forest plot representing the hazard ratio for VT recurrence of the SNP in the female subgroup for all studies that contributed to this subgroup analysis. C) Regional association plot around the SNP in the female subgroup analysis. N: sample size / NREC: number of VT recurrences / EAF: effect allele frequency / INFO: imputation quality score / HR: Hazard ratio.

**Supplementary Figure S7:** Associations of rs115157965-T identified in the male subgroup. A) Forest plot representing the hazard ratio for VT recurrence of the SNP-effects across the subgroups and in the main analysis. B) Forest plot representing the hazard ratio for VT recurrence of the SNP in the male subgroup for all studies that contributed to this subgroup analysis. C) Regional association plot around the SNP in the male subgroup analysis. N: sample size / NREC: number of VT recurrences / EAF: effect allele frequency / INFO: imputation quality score / HR: Hazard ratio.

**Supplementary Figure S8:** Associations of rs116666788-A identified in the DVT only as first VT subgroup. A) Forest plot representing the hazard ratio for VT recurrence of the SNP-effects across the subgroups and in the main analysis. B) Forest plot representing the hazard ratio for VT recurrence of the SNP in the DVT subgroup for all studies that contributed to this subgroup analysis. C) Regional association plot around the SNP in the DVT subgroup analysis. N: sample size / NREC: number of VT recurrences / EAF: effect allele frequency / INFO: imputation quality score / HR: Hazard ratio.

**Supplementary Figure S9:** Associations of rs117366080-A identified in the DVT only as first VT subgroup. A) Forest plot representing the hazard ratio for VT recurrence of the SNP-effects across the subgroups and in the main analysis. B) Forest plot representing the hazard ratio for VT recurrence of the SNP in the DVT subgroup for all studies that contributed to this subgroup analysis. C) Regional association plot around the SNP in the DVT subgroup analysis. N: sample size / NREC: number of VT recurrences / EAF: effect allele frequency / INFO: imputation quality score / HR: Hazard ratio.

**Supplementary Figure S10:** Associations of rs45562031-T identified in the PE as first VT subgroup. A) Forest plot representing the hazard ratio for VT recurrence of the SNP-effects across the subgroups and in the main analysis. B) Forest plot representing the hazard ratio for VT recurrence of the SNP in the PE subgroup for all studies that contributed to this subgroup analysis. C) Regional association plot around the SNP in the PE subgroup analysis. N: sample size / NREC: number of VT recurrences / EAF: effect allele frequency / INFO: imputation quality score / HR: Hazard ratio.

**Supplementary Figure S11:** Associations of rs140802879-G identified in the PE as first VT subgroup. A) Forest plot representing the hazard ratio for VT recurrence of the SNP-effects across the subgroups and in the main analysis. B) Forest plot representing the hazard ratio for VT recurrence of the SNP in the PE subgroup for all studies that contributed to this subgroup analysis. C) Regional association plot around the SNP in the PE subgroup analysis. N: sample size / NREC: number of VT recurrences / EAF: effect allele frequency / INFO: imputation quality score / HR: Hazard ratio.

**Supplementary Figure S12:** Associations of rs34475559-A identified in the PE as first VT subgroup. A) Forest plot representing the hazard ratio for VT recurrence of the SNP-effects across the subgroups and in the main analysis. B) Forest plot representing the hazard ratio for VT recurrence of the SNP in the PE subgroup for all studies that contributed to this subgroup analysis. C) Regional association plot around the SNP in the PE subgroup analysis. N: sample size / NREC: number of VT recurrences / EAF: effect allele frequency / INFO: imputation quality score / HR: Hazard ratio.

**Supplementary Figure S13:** Associations of rs6474692-A identified in the PE as first VT subgroup. A) Forest plot representing the hazard ratio for VT recurrence of the SNP-effects across the subgroups and in the main analysis. B) Forest plot representing the hazard ratio for VT recurrence of the SNP in the PE subgroup for all studies that contributed to this subgroup analysis. C) Regional association plot around the SNP in the PE subgroup analysis. N: sample size / NREC: number of VT recurrences / EAF: effect allele frequency / INFO: imputation quality score / HR: Hazard ratio.

**Supplementary Figure S14:** Associations of rs148611543-A identified in the unprovoked first VT subgroup. A) Forest plot representing the hazard ratio for VT recurrence of the SNP-effects across the subgroups and in the main analysis. B) Forest plot representing the hazard ratio for VT recurrence of the SNP in the unprovoked subgroup for all studies that contributed to this subgroup analysis. C) Regional association plot around the SNP in the unprovoked subgroup analysis. N: sample size / NREC: number of VT recurrences / EAF: effect allele frequency / INFO: imputation quality score / HR: Hazard ratio.

**Supplementary Figure S15:** Associations of rs117509298-C identified in the unprovoked first VT subgroup. A) Forest plot representing the hazard ratio for VT recurrence of the SNP-effects across the subgroups and in the main analysis. B) Forest plot representing the hazard ratio for VT recurrence of the SNP in the unprovoked subgroup for all studies that contributed to this subgroup analysis. C) Regional association plot around the SNP in the unprovoked subgroup analysis. N: sample size / NREC: number of VT recurrences / EAF: effect allele frequency / INFO: imputation quality score / HR: Hazard ratio.

**Supplementary Figure S16:** Associations of rs148811092-A identified in the provoked first VT subgroup. A) Forest plot representing the hazard ratio for VT recurrence of the SNP-effects across the subgroups and in the main analysis. B) Forest plot representing the hazard ratio for VT recurrence of the SNP in the provoked subgroup for all studies that contributed to this subgroup analysis. C) Regional association plot around the SNP in the provoked subgroup analysis. N: sample size / NREC: number of VT recurrences / EAF: effect allele frequency / INFO: imputation quality score / HR: Hazard ratio.

**Supplementary Figure S17:** Associations of rs3744462-G identified in the provoked first VT subgroup. A) Forest plot representing the hazard ratio for VT recurrence of the SNP-effects across the subgroups and in the main analysis. B) Forest plot representing the hazard ratio for VT recurrence of the SNP in the provoked subgroup for all studies that contributed to this subgroup analysis. C) Regional association plot around the SNP in the provoked subgroup analysis. N: sample size / NREC: number of VT recurrences / EAF: effect allele frequency / INFO: imputation quality score / HR: Hazard ratio.

**Supplementary Figure S18:** Associations of rs112440311-T identified in the provoked first VT subgroup. A) Forest plot representing the hazard ratio for VT recurrence of the SNP-effects across the subgroups and in the main analysis. B) Forest plot representing the hazard ratio for VT recurrence of the SNP in the provoked subgroup for all studies that contributed to this subgroup analysis. C) Regional association plot around the SNP in the provoked subgroup analysis. N: sample size / NREC: number of VT recurrences / EAF: effect allele frequency / INFO: imputation quality score / HR: Hazard ratio.

**Supplementary Figure S19:** Associations of rs111665167-A identified in the provoked first VT subgroup. A) Forest plot representing the hazard ratio for VT recurrence of the SNP-effects across the subgroups and in the main analysis. B) Forest plot representing the hazard ratio for VT recurrence of the SNP in the provoked subgroup for all studies that contributed to this subgroup analysis. C) Regional association plot around the SNP in the provoked subgroup analysis. N: sample size / NREC: number of VT recurrences / EAF: effect allele frequency / INFO: imputation quality score / HR: Hazard ratio.

**Supplementary Figure S20:** Associations of rs6475173-A identified in the provoked first VT subgroup. A) Forest plot representing the hazard ratio for VT recurrence of the SNP-effects across the subgroups and in the main analysis. B) Forest plot representing the hazard ratio for VT recurrence of the SNP in the provoked subgroup for all studies that contributed to this subgroup analysis. C) Regional association plot around the SNP in the provoked subgroup analysis. N: sample size / NREC: number of VT recurrences / EAF: effect allele frequency / INFO: imputation quality score / HR: Hazard ratio.

**Supplementary Figure S21:** Associations of rs62174841-T identified in the provoked first VT subgroup. A) Forest plot representing the hazard ratio for VT recurrence of the SNP-effects across the subgroups and in the main analysis. B) Forest plot representing the hazard ratio for VT recurrence of the SNP in the provoked subgroup for all studies that contributed to this subgroup analysis. C) Regional association plot around the SNP in the provoked subgroup analysis. N: sample size / NREC: number of VT recurrences / EAF: effect allele frequency / INFO: imputation quality score / HR: Hazard ratio.

**Supplementary Figure S22:** Associations of rs180737225-G identified in the provoked first VT subgroup. A) Forest plot representing the hazard ratio for VT recurrence of the SNP-effects across the subgroups and in the main analysis. B) Forest plot representing the hazard ratio for VT recurrence of the SNP in the provoked subgroup for all studies that contributed to this subgroup analysis. C) Regional association plot around the SNP in the provoked subgroup analysis. N: sample size / NREC: number of VT recurrences / EAF: effect allele frequency / INFO: imputation quality score / HR: Hazard ratio.

**Supplementary Figure S23:** Forest plot representing the associations of rs6035-T in the studies and the whole meta-analysis on VT recurrence. N: sample size / NREC: number of VT recurrences / EAF: effect allele frequency / INFO: imputation quality score / HR: Hazard ratio.

**Supplementary Figure S24:** Forest plot representing the associations of rs9946608-C in the studies and the whole meta-analysis on VT recurrence. N: sample size / NREC: number of VT recurrences / EAF: effect allele frequency / INFO: imputation quality score / HR: Hazard ratio.

## References

1. Raskob GE, Angchaisuksiri P, Blanco AN, et al. Thrombosis: A Major Contributor to Global Disease Burden. ATVB. 2014;34(11):2363–2371.

2. Cohen AT, Agnelli G, Anderson FA, et al. Venous thromboembolism (VTE) in Europe. The number of VTE events and associated morbidity and mortality. Thromb Haemost. 2007;98(4):756–764.

3. Stevens SM, Woller SC, Kreuziger LB, et al. Antithrombotic Therapy for VTE Disease: Second Update of the CHEST Guideline and Expert Panel Report. Chest. 2021;160(6):e545–e608.

4. Delluc A, Tromeur C, Le Ven F, et al. Current incidence of venous thromboembolism and comparison with 1998: a community-based study in Western France. Thromb Haemost. 2016;116(11):967–974.

5. Couturaud F, Sanchez O, Pernod G, et al. Six Months vs Extended Oral Anticoagulation After a First Episode of Pulmonary Embolism: The PADIS-PE Randomized Clinical Trial. JAMA. 2015;314(1):31.

6. Khan F, Rahman A, Carrier M, et al. Long term risk of symptomatic recurrent venous thromboembolism after discontinuation of anticoagulant treatment for first unprovoked venous thromboembolism event: systematic review and meta-analysis. BMJ. 2019;l4363.

7. Lecumberri R, Alfonso A, Jiménez D, et al. Dynamics of case-fatalilty rates of recurrent thromboembolism and major bleeding in patients treated for venous thromboembolism. Thromb Haemost. 2013;110(4):834–843.

8. Tagalakis V, Patenaude V, Kahn SR, Suissa S. Treatment patterns of venous thromboembolism in a real-world population: the Q-VTE study cohort. Thromb Res. 2014;134(4):795–802.

9. 2014 ESC Guidelines on the diagnosis and management of acute pulmonary embolism | European Heart Journal | Oxford Academic.

10. Tosetto A, Iorio A, Marcucci M, et al. Predicting disease recurrence in patients with previous unprovoked venous thromboembolism: a proposed prediction score (DASH). J Thromb Haemost. 2012;10(6):1019–1025.

11. Eichinger S, Heinze G, Jandeck LM, Kyrle PA. Risk assessment of recurrence in patients with unprovoked deep vein thrombosis or pulmonary embolism: the Vienna prediction model. Circulation. 2010;121(14):1630–1636.

12. Timp JF, Braekkan SK, Lijfering WM, et al. Prediction of recurrent venous thrombosis in all patients with a first venous thrombotic event: The Leiden Thrombosis Recurrence Risk Prediction model (L-TRRiP). PLoS Med. 2019;16(10):e1002883.

13. De Winter MA, Büller HR, Carrier M, et al. Recurrent venous thromboembolism and bleeding with extended anticoagulation: the VTE-PREDICT risk score. European Heart Journal. 2023;44(14):1231–1244.

14. Rodger MA, Le Gal G, Anderson DR, et al. Validating the HERDOO2 rule to guide treatment duration for women with unprovoked venous thrombosis: multinational prospective cohort management study. BMJ. 2017;356:j1065.

15. Kyrle PauA, Eischer L, Šinkovec H, Eichinger S. Factor XI and recurrent venous thrombosis: an observational cohort study. Journal of Thrombosis and Haemostasis. 2019;17(5):782–786.

16. Hoke M, Kyrle PA, Minar E, et al. Tissue factor pathway inhibitor and the risk of recurrent venous thromboembolism. Thromb Haemost. 2005;94(4):787–790.

17. Kyrle PA, Minar E, Hirschl M, et al. High Plasma Levels of Factor VIII and the Risk of Recurrent Venous Thromboembolism. New England Journal of Medicine. 2000;343(7):457–462.

18. Edvardsen MS, Hindberg K, Hansen E-S, et al. Plasma levels of von Willebrand factor and future risk of incident venous thromboembolism. Blood Adv. 2021;5(1):224–232.

19. Rodger MA, Kahn SR, Wells PS, et al. Identifying unprovoked thromboembolism patients at low risk for recurrence who can discontinue anticoagulant therapy. Canadian Medical Association Journal. 2008;179(5):417–426.

20. Christiansen SC, Cannegieter SC, Koster T, Vandenbroucke JP, Rosendaal FR. Thrombophilia, clinical factors, and recurrent venous thrombotic events. JAMA. 2005;293(19):2352–2361.

21. Baglin T, Luddington R, Brown K, Baglin C. Incidence of recurrent venous thromboembolism in relation to clinical and thrombophilic risk factors: prospective cohort study. Lancet. 2003;362(9383):523–526.

22. Prandoni P, Noventa F, Ghirarduzzi A, et al. The risk of recurrent venous thromboembolism after discontinuing anticoagulation in patients with acute proximal deep vein thrombosis or pulmonary embolism. A prospective cohort study in 1,626 patients. Haematologica. 2007;92(2):199–205.

23. Lutsey PL, Zakai NA. Epidemiology and prevention of venous thromboembolism. Nat Rev Cardiol. 2023;20(4):248–262.

24. Couturaud F, Leroyer C, Tromeur C, et al. Factors that predict thrombosis in relatives of patients with venous thromboembolism. Blood. 2014;124(13):2124–2130.

25. Klarin D, Busenkell E, Judy R, et al. Genome-wide association analysis of venous thromboembolism identifies new risk loci and genetic overlap with arterial vascular disease. Nat Genet. 2019;51(11):1574–1579.

26. Thibord F, Klarin D, Brody JA, et al. Cross-Ancestry Investigation of Venous Thromboembolism Genomic Predictors. Circulation. 2022;146(16):1225–1242.

27. de Haan HG, van Hylckama Vlieg A, Germain M, et al. Genome-Wide Association Study Identifies a Novel Genetic Risk Factor for Recurrent Venous Thrombosis. Circulation: Genomic and Precision Medicine. 2018;11(2):e001827.

28. Munsch G, Goumidi L, van Hylckama Vlieg A, et al. Association of ABO blood groups with venous thrombosis recurrence in middle-aged patients: insights from a weighted Cox analysis dedicated to ambispective design. BMC Med Res Methodol. 2023;23(1):99.

29. Mägi R, Morris AP. GWAMA: software for genome-wide association meta-analysis. BMC Bioinformatics. 2010;11(1):288.

30. Gusev A, Ko A, Shi H, et al. Integrative approaches for large-scale transcriptome-wide association studies. Nat Genet. 2016;48(3):245–252.

31. Giambartolomei C, Vukcevic D, Schadt EE, et al. Bayesian Test for Colocalisation between Pairs of Genetic Association Studies Using Summary Statistics. PLoS Genet. 2014;10(5):e1004383.

32. Burgess S, Davey Smith G, Davies NM, et al. Guidelines for performing Mendelian randomization investigations: update for summer 2023. Wellcome Open Res. 2019;4:186.

33. Hemani G, Haycock P, Zheng J, et al. TwoSampleMR R package. 2023;

34. Ferkingstad E, Sulem P, Atlason BA, et al. Large-scale integration of the plasma proteome with genetics and disease. Nat Genet. 2021;53(12):1712–1721.

35. Pietzner M, Wheeler E, Carrasco-Zanini J, et al. Mapping the proteo-genomic convergence of human diseases. Science. 2021;374(6569):eabj1541.

36. Sun BB, Chiou J, Traylor M, et al. Plasma proteomic associations with genetics and health in the UK Biobank. Nature. 2023;622(7982):329–338.

37. Chen Y, Lu T, Pettersson-Kymmer U, et al. Genomic atlas of the plasma metabolome prioritizes metabolites implicated in human diseases. Nat Genet. 2023;55(1):44–53.

38. van der Harst P, Verweij N. Identification of 64 Novel Genetic Loci Provides an Expanded View on the Genetic Architecture of Coronary Artery Disease. Circ Res. 2018;122(3):433–443.

39. Temprano-Sagrera G, Sitlani CM, Bone WP, et al. Multi-phenotype analyses of hemostatic traits with cardiovascular events reveal novel genetic associations. Journal of Thrombosis and Haemostasis. 2022;20(6):1331–1349.

40. Hu C, Zuo K, Li K, et al. p38/JNK Is Required for the Proliferation and Phenotype Changes of Vascular Smooth Muscle Cells Induced by L3MBTL4 in Essential Hypertension. Int J Hypertens. 2020;2020:3123968.

41. Liu X, Hu C, Bao M, et al. Genome Wide Association Study Identifies L3MBTL4 as a Novel Susceptibility Gene for Hypertension. Sci Rep. 2016;6(1):30811.

42. de Vries PS, Sabater-Lleal M, Huffman JE, et al. A genome-wide association study identifies new loci for factor VII and implicates factor VII in ischemic stroke etiology. Blood. 2019;133(9):967– 977.

43. Iruela-Arispe ML, Luque A, Lee N. Thrombospondin modules and angiogenesis. Int J Biochem Cell Biol. 2004;36(6):1070–1078.

44. Rühle F, Witten A, Barysenka A, et al. Rare genetic variants in SMAP1, B3GAT2, and RIMS1 contribute to pediatric venous thromboembolism. Blood. 2017;129(6):783–790.

45. Leung LL. Role of thrombospondin in platelet aggregation. J. Clin. Invest. 1984;74(5):1764– 1772.

46. Rauluseviciute I, Riudavets-Puig R, Blanc-Mathieu R, et al. JASPAR 2024: 20th anniversary of the open-access database of transcription factor binding profiles. Nucleic Acids Research. 2024;52(D1):D174–D182.

47. Ren A, Gan Q, Han W, et al. Endothelial GATA5 positively regulates angiogenesis via cathepsin S-mediated Angpt2/Flk1 and MMP2/9 signaling pathways. Biochemical and Biophysical Research Communications. 2022;609:111–118.

48. Cantor AB. Chapter 28 - Thrombocytopoiesis. Hematology (Seventh Edition). 2018;334–349.

49. Barenboim M, Zoltick BJ, Guo Y, Weinberger DR. MicroSNiPer: a web tool for prediction of SNP effects on putative microRNA targets. Hum Mutat. 2010;31(11):1223–1232.

50. Godin O, Dufouil C, Maillard P, et al. White matter lesions as a predictor of depression in the elderly: the 3C-Dijon study. Biol Psychiatry. 2008;63(7):663–669.

51. Ward LD, Kellis M. HaploReg: a resource for exploring chromatin states, conservation, and regulatory motif alterations within sets of genetically linked variants. Nucleic Acids Res. 2012;40(Database issue):D930–934.

52. Gailani D, Gruber A. Targeting factor XI and factor XIa to prevent thrombosis. Blood. 2024;143(15):1465–1475.

53. Sabater-Lleal M, Martinez-Perez A, Buil A, et al. A genome-wide association study identifies KNG1 as a genetic determinant of plasma factor XI Level and activated partial thromboplastin time. Arterioscler Thromb Vasc Biol. 2012;32(8):2008–2016.

54. Bapat J, Yamamoto TM, Woodruff ER, et al. CASC4/GOLM2 drives high grade serous carcinoma anoikis resistance through the recycling of EGFR. Cancer Gene Ther. 2023;

55. Mootoosamy C, Kondyli M, Serfaty SA, et al. IL16 and factor V gene variations are associated with asparaginase-related thrombosis in childhood acute lymphoblastic leukemia patients. Pharmacogenomics. 2023;24(4):199–206.

56. de Souza VH, de Alencar JB, Tiyo BT, et al. Association of functional IL16 polymorphisms with cancer and cardiovascular disease: a meta-analysis. Oncotarget. 2020;11(36):3405–3417.

57. Richmond J, Tuzova M, Cruikshank W, Center D. Regulation of Cellular Processes by Interleukin_16 in Homeostasis and Cancer. Journal Cellular Physiology. 2014;229(2):139–147.

58. He H, Ding M, Zhu Y, et al. Interleukin-16/STAT6 recruits CBP/p300 to upregulate TIMP-3 and promote atherosclerotic plaque stability. 2023;

59. Grönberg C, Bengtsson E, Fredrikson GN, et al. Human Carotid Plaques With High Levels of Interleukin-16 Are Associated With Reduced Risk for Cardiovascular Events. Stroke. 2015;46(10):2748–2754.

60. Grönberg C, Asciutto G, Persson A, et al. Endarterectomy patients with elevated levels of circulating IL-16 have fewer cardiovascular events during follow-up. Cytokine. 2016;85:137–139.

61. Badimon L, Vilahur G. Thrombosis formation on atherosclerotic lesions and plaque rupture. J Intern Med. 2014;276(6):618–632.

62. Puteri MU, Azmi NU, Kato M, Saputri FC. PCSK9 Promotes Cardiovascular Diseases: Recent Evidence about Its Association with Platelet Activation-Induced Myocardial Infarction. Life (Basel). 2022;12(2):190.

63. Marston NA, Gurmu Y, Melloni GEM, et al. The Effect of PCSK9 (Proprotein Convertase Subtilisin/Kexin Type 9) Inhibition on the Risk of Venous Thromboembolism. Circulation. 2020;141(20):1600–1607.

64. Wang H, Wang Q, Wang J, et al. Proprotein convertase subtilisin/kexin type 9 (PCSK9) Deficiency is Protective Against Venous Thrombosis in Mice. Sci Rep. 2017;7(1):14360.

65. Kheirkhah A, Schachtl-Riess JF, Lamina C, et al. Meta-GWAS on PCSK9 concentrations reveals associations of novel loci outside the PCSK9 locus in White populations. Atherosclerosis. 2023;386:117384.

66. Aragam KG, Jiang T, Goel A, et al. Discovery and systematic characterization of risk variants and genes for coronary artery disease in over a million participants. Nat Genet. 2022;54(12):1803– 1815.

67. Hartiala JA, Han Y, Jia Q, et al. Genome-wide analysis identifies novel susceptibility loci for myocardial infarction. European Heart Journal. 2021;42(9):919–933.

68. Mishra A, Malik R, Hachiya T, et al. Stroke genetics informs drug discovery and risk prediction across ancestries. Nature. 2022;611(7934):115–123.

69. Graham SE, Clarke SL, Wu K-HH, et al. The power of genetic diversity in genome-wide association studies of lipids. Nature. 2021;600(7890):675–679.

70. Stewart LK, Sarmiento EJ, Kline JA. Statin Use is Associated with Reduced Risk of Recurrence in Patients with Venous Thromboembolism. Am J Med. 2020;133(8):930–935.e8.

71. Li R, Yuan M, Yu S, et al. Effect of statins on the risk of recurrent venous thromboembolism: A systematic review and meta-analysis. Pharmacol Res. 2021;165:105413.

72. Biedermann JS, Kruip MJHA, van der Meer FJ, et al. Rosuvastatin use improves measures of coagulation in patients with venous thrombosis. Eur Heart J. 2018;39(19):1740–1747.

73. Chang W, Sheu C, Liu K, et al. Identification of mutations in SLC4A1, GP1BA and HFE in a family with venous thrombosis of unknown cause by next_generation sequencing. Exp Ther Med. 2018;

74. Zhong J, Dong J, Ruan W, Duan X. Potential Theranostic Roles of SLC4 Molecules in Human Diseases. IJMS. 2023;24(20):15166.

75. Douketis JD. Risk of Fatal Pulmonary Embolism in Patients With Treated Venous Thromboembolism. JAMA. 1998;279(6):458.

76. Razzaq M, Iglesias MJ, Ibrahim-Kosta M, et al. An artificial neural network approach integrating plasma proteomics and genetic data identifies PLXNA4 as a new susceptibility locus for pulmonary embolism. Sci Rep. 2021;11(1):14015.

77. Machiela MJ, Chanock SJ. LDlink: a web-based application for exploring population-specific haplotype structure and linking correlated alleles of possible functional variants. Bioinformatics. 2015;31(21):3555–3557.

78. Goumidi L, Thibord F, Wiggins KL, et al. Association between ABO haplotypes and the risk of venous thrombosis: impact on disease risk estimation. Blood. 2021;137(17):2394–2402.

