## Supplementary Figures for "Genomic Landscape of Thrombosis Recurrence Risk Across Venous Thromboembolism Subtypes"

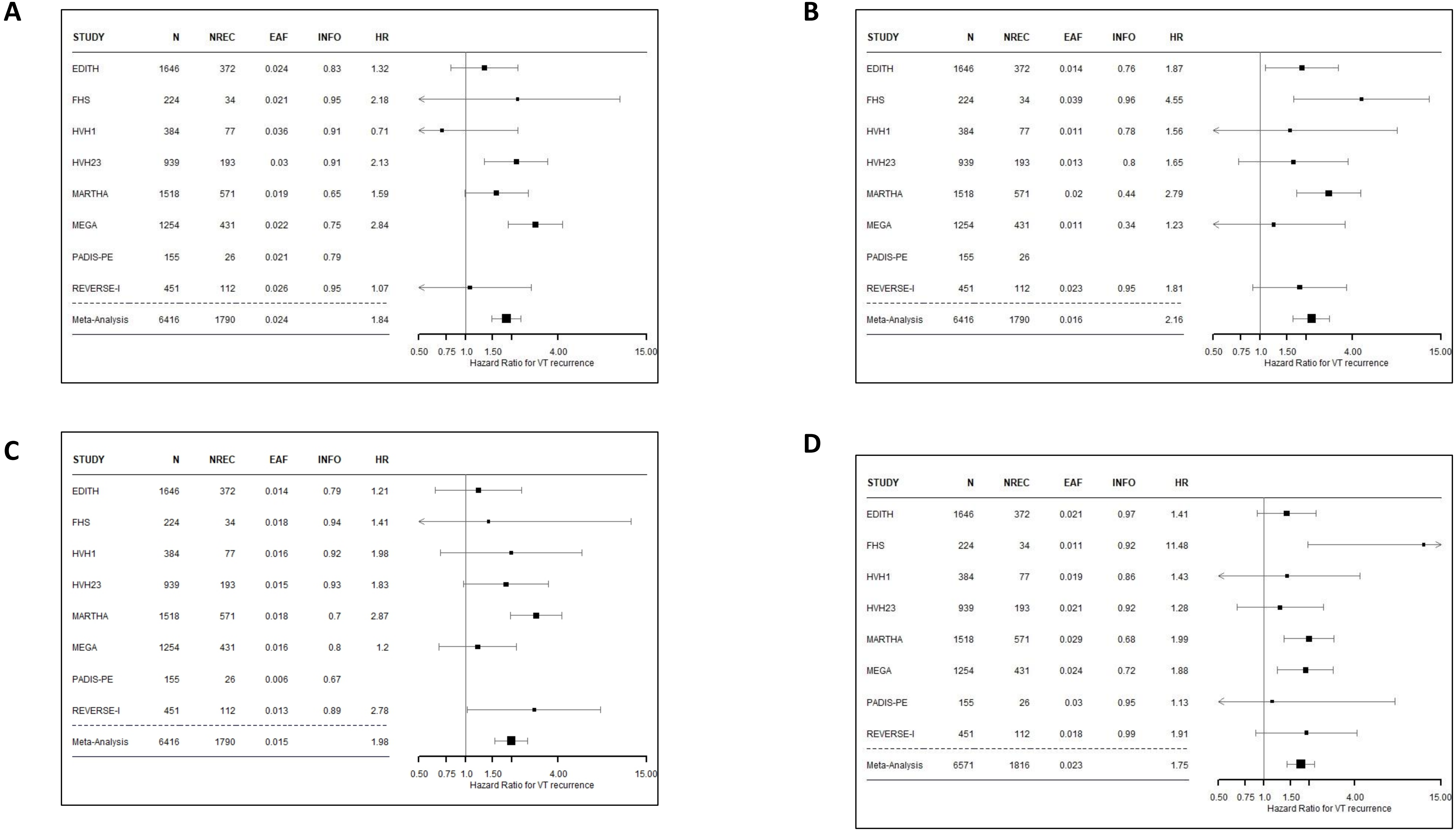
A
STUDY N NREC EAF INFO HR
EDITH 1646 372 0.024 0.83 1.32
FHS 224 34 0.021 0.95 2.18
HVH1 384 77 0.036 0.91 0.71
HVH23 939 193 0.03 0.91 2.13
MARTHA 1518 571 0.019 0.65 1.59
MEGA 1254 431 0.022 0.75 2.84
PADIS-PE 155 26 0.021 0.79
REVERSE-I 451 112 0.026 0.95 1.07
Meta-Analysis 6416 1790 0.024 1.84
0.50 0.75 1.0 1.50 4.00 15.00
Hazard Ratio for VT recurrence
B
STUDY N NREC EAF INFO HR
EDITH 1646 372 0.014 0.76 1.87
FHS 224 34 0.039 0.96 4.55
HVH1 384 77 0.011 0.78 1.56
HVH23 939 193 0.013 0.8 1.65
MARTHA 1518 571 0.02 0.44 2.79
MEGA 1254 431 0.011 0.34 1.23
PADIS-PE 155 26
REVERSE-I 451 112 0.023 0.95 1.81
Meta-Analysis 6416 1790 0.016 2.16
0.50 0.75 1.0 1.50 4.00 15.00
Hazard Ratio for VT recurrence
C
STUDY N NREC EAF INFO HR
EDITH 1646 372 0.014 0.79 1.21
FHS 224 34 0.018 0.94 1.41
HVH1 384 77 0.016 0.92 1.98
HVH23 939 193 0.015 0.93 1.83
MARTHA 1518 571 0.018 0.7 2.87
MEGA 1254 431 0.016 0.8 1.2
PADIS-PE 155 26 0.006 0.67
REVERSE-I 451 112 0.013 0.89 2.78
Meta-Analysis 6416 1790 0.015 1.98
0.50 0.75 1.0 1.50 4.00 15.00
Hazard Ratio for VT recurrence
D
STUDY N NREC EAF INFO HR
EDITH 1646 372 0.021 0.97 1.41
FHS 224 34 0.011 0.92 11.48
HVH1 384 77 0.019 0.86 1.43
HVH23 939 193 0.021 0.92 1.28
MARTHA 1518 571 0.029 0.68 1.99
MEGA 1254 431 0.024 0.72 1.88
PADIS-PE 155 26 0.03 0.95 1.13
REVERSE-I 451 112 0.018 0.99 1.91
Meta-Analysis 6571 1816 0.023 1.75
0.50 0.75 1.0 1.50 4.00 15.00
Hazard Ratio for VT recurrence

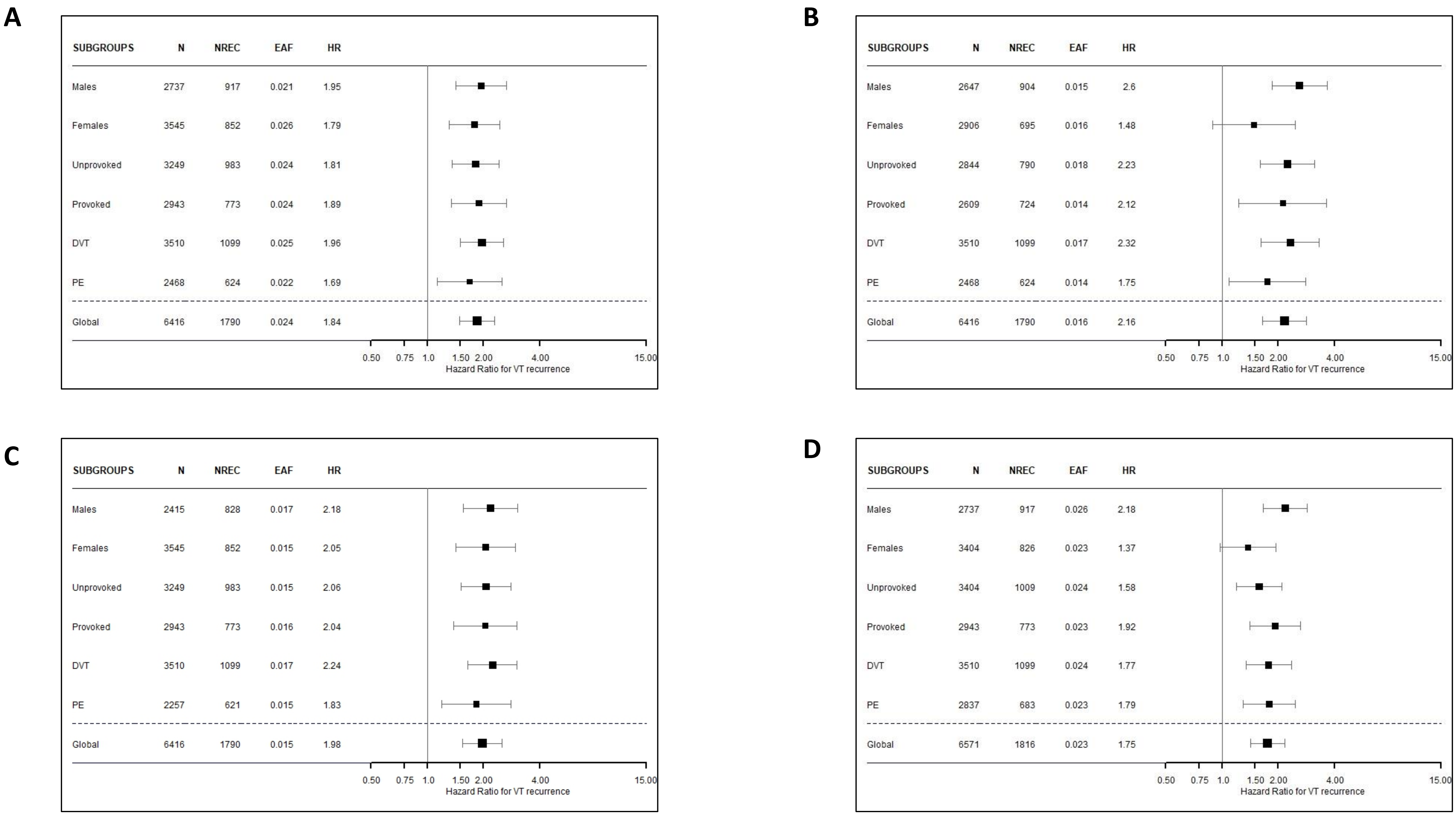
A
SUBGROUPS N NREC EAF HR
Males 2737 917 0.021 1.95
Females 3545 852 0.026 1.79
Unprovoked 3249 983 0.024 1.81
Provoked 2943 773 0.024 1.89
DVT 3510 1099 0.025 1.96
PE 2468 624 0.022 1.69
Global 6416 1790 0.024 1.84
0.50 0.75 1.0 1.50 2.00 4.00 15.00
Hazard Ratio for VT recurrence
B
SUBGROUPS N NREC EAF HR
Males 2647 904 0.015 2.6
Females 2906 695 0.016 1.48
Unprovoked 2844 790 0.018 2.23
Provoked 2609 724 0.014 2.12
DVT 3510 1099 0.017 2.32
PE 2468 624 0.014 1.75
Global 6416 1790 0.016 2.16
0.50 0.75 1.0 1.50 2.00 4.00 15.00
Hazard Ratio for VT recurrence
C
SUBGROUPS N NREC EAF HR
Males 2415 828 0.017 2.18
Females 3545 852 0.015 2.05
Unprovoked 3249 983 0.015 2.06
Provoked 2943 773 0.016 2.04
DVT 3510 1099 0.017 2.24
PE 2257 621 0.015 1.83
Global 6416 1790 0.015 1.98
0.50 0.75 1.0 1.50 2.00 4.00 15.00
Hazard Ratio for VT recurrence
D
SUBGROUPS N NREC EAF HR
Males 2737 917 0.026 2.18
Females 3404 826 0.023 1.37
Unprovoked 3404 1009 0.024 1.58
Provoked 2943 773 0.023 1.92
DVT 3510 1099 0.024 1.77
PE 2837 683 0.023 1.79
Global 6571 1816 0.023 1.75
0.50 0.75 1.0 1.50 2.00 4.00 15.00
Hazard Ratio for VT recurrence

| RSID | *ABO* Haplotypes | | | | |
|---|---|---|---|---|---|
| **ABO tag SNPs** | | | | | |
| rs8176719 | - | C | C | C | C |
| rs2519093 | C | T | C | C | C |
| rs1053878 | G | G | A | G | G |
| rs8176743 | C | C | C | T | C |
| rs41302905 | C | C | C | C | T |
| **GOLM2 pQTLs** | | | | | |
| rs550057 | C | T | T | C | C |
| rs687289 | G | A | A | A | G |
| **Frequency** | **60% - O1** | **18% - A1** | **9% - A2** | **9% - B** | **3% - O2** |

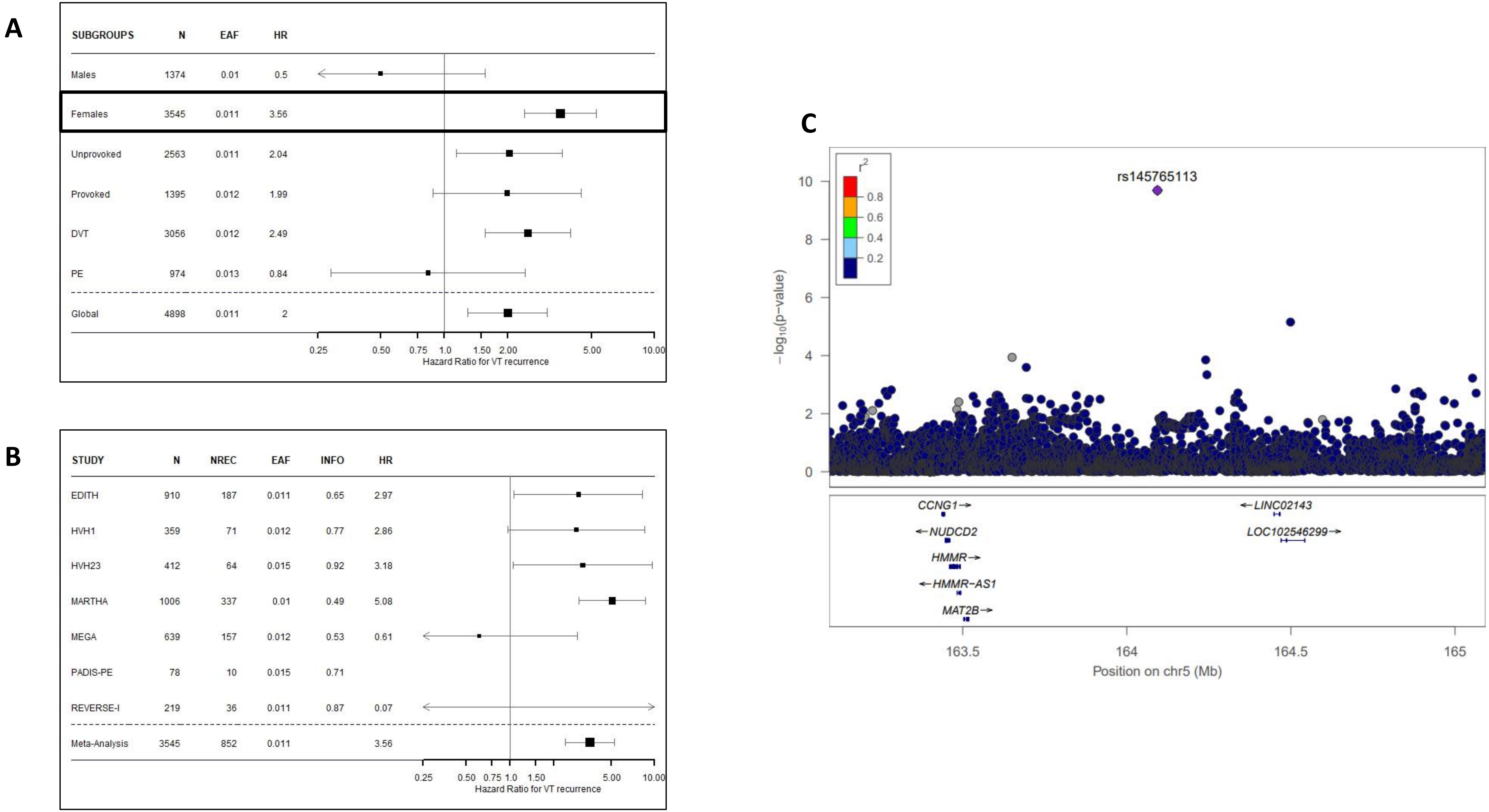
A
SUBGROUPS N EAF HR
Males 1374 0.01 0.5
Females 3545 0.011 3.56
Unprovoked 2563 0.011 2.04
Provoked 1395 0.012 1.99
DVT 3056 0.012 2.49
PE 974 0.013 0.84
Global 4898 0.011 2
0.25 0.50 0.75 1.0 1.50 2.00 5.00 10.00
Hazard Ratio for VT recurrence
B
STUDY N NREC EAF INFO HR
EDITH 910 187 0.011 0.65 2.97
HVH1 359 71 0.012 0.77 2.86
HVH23 412 64 0.015 0.92 3.18
MARTHA 1006 337 0.01 0.49 5.08
MEGA 639 157 0.012 0.53 0.61
PADIS-PE 78 10 0.015 0.71
REVERSE-I 219 36 0.011 0.87 0.07
Meta-Analysis 3545 852 0.011 3.56
0.25 0.50 0.75 1.0 1.50 5.00 10.00
Hazard Ratio for VT recurrence
C
rs145765113
r2
0.8
0.6
0.4
0.2
−log10(p-value)
CCNG1→
←NUDCD2
HMMR→
←HMMR-AS1
MAT2B→
←LINC02143
LOC102546299→
163.5 164 164.5 165
Position on chr5 (Mb)

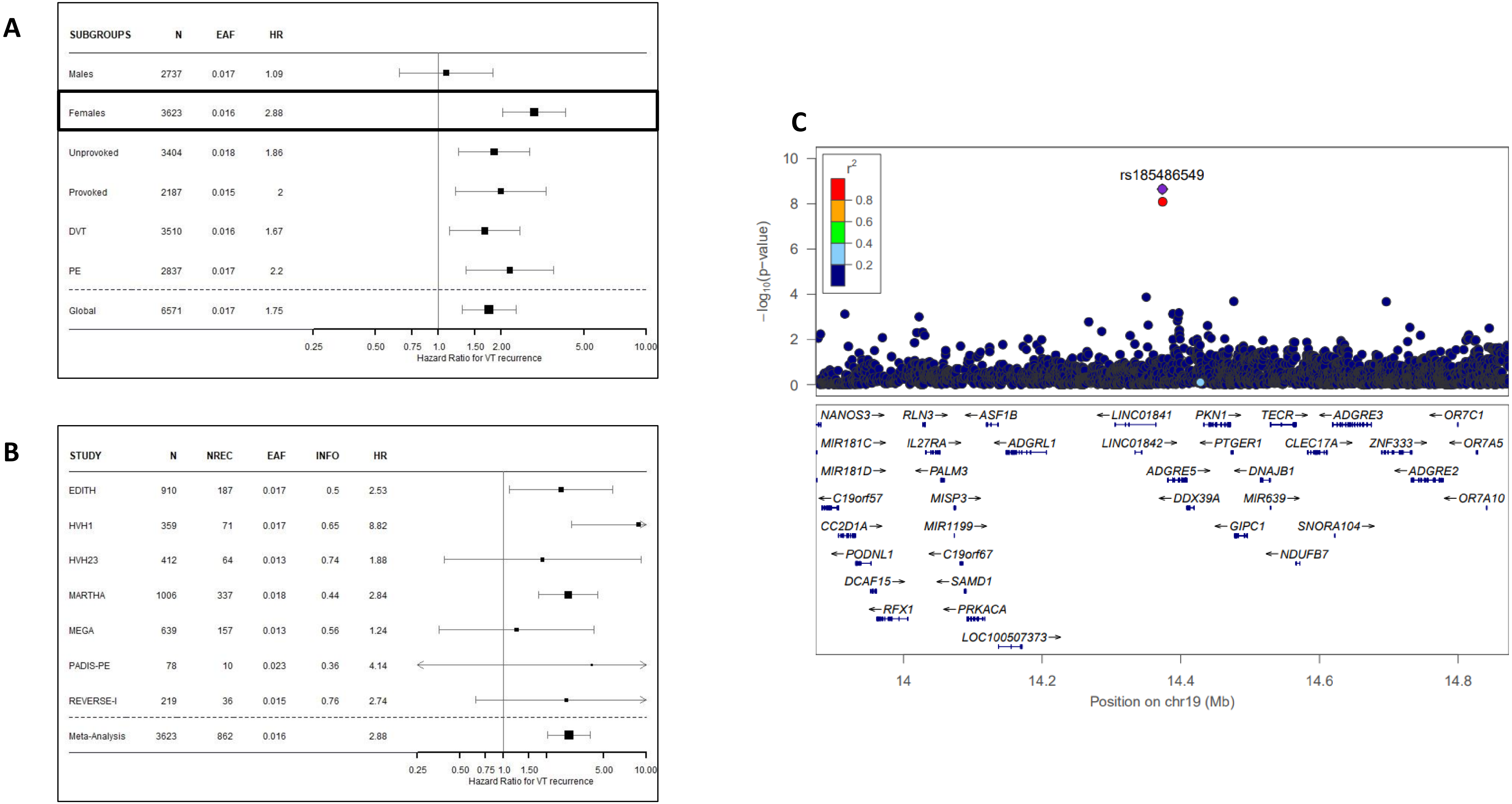
A
SUBGROUPS N EAF HR
Males 2737 0.017 1.09
Females 3623 0.016 2.88
Unprovoked 3404 0.018 1.86
Provoked 2187 0.015 2
DVT 3510 0.016 1.67
PE 2837 0.017 2.2
Global 6571 0.017 1.75
0.25 0.50 0.75 1.0 1.50 2.00 5.00 10.00
Hazard Ratio for VT recurrence
B
STUDY N NREC EAF INFO HR
EDITH 910 187 0.017 0.5 2.53
HVH1 359 71 0.017 0.65 8.82
HVH23 412 64 0.013 0.74 1.88
MARTHA 1006 337 0.018 0.44 2.84
MEGA 639 157 0.013 0.56 1.24
PADIS-PE 78 10 0.023 0.36 4.14
REVERSE-I 219 36 0.015 0.76 2.74
Meta-Analysis 3623 862 0.016 2.88
0.25 0.50 0.75 1.0 1.50 5.00 10.00
Hazard Ratio for VT recurrence
C
rs185486549
r2
0.8
0.6
0.4
0.2
−log10(p-value)
10
8
6
4
2
0
NANOS3 RLN3 ASF1B LINC01841 PKN1 TECR ADGRE3 OR7C1
MIR181C IL27RA ADGRL1 LINC01842 PTGER1 CLEC17A ZNF333 OR7A5
MIR181D PALM3 ADGRE5 DNAJB1 ADGRE2
C19orf57 MISP3 DDX39A MIR639 OR7A10
CC2D1A MIR1199 GIPC1 SNORA104
PODNL1 C19orf67 NDUFB7
DCAF15 SAMD1
RFX1 PRKACA
LOC100507373
14 14.2 14.4 14.6 14.8
Position on chr19 (Mb)

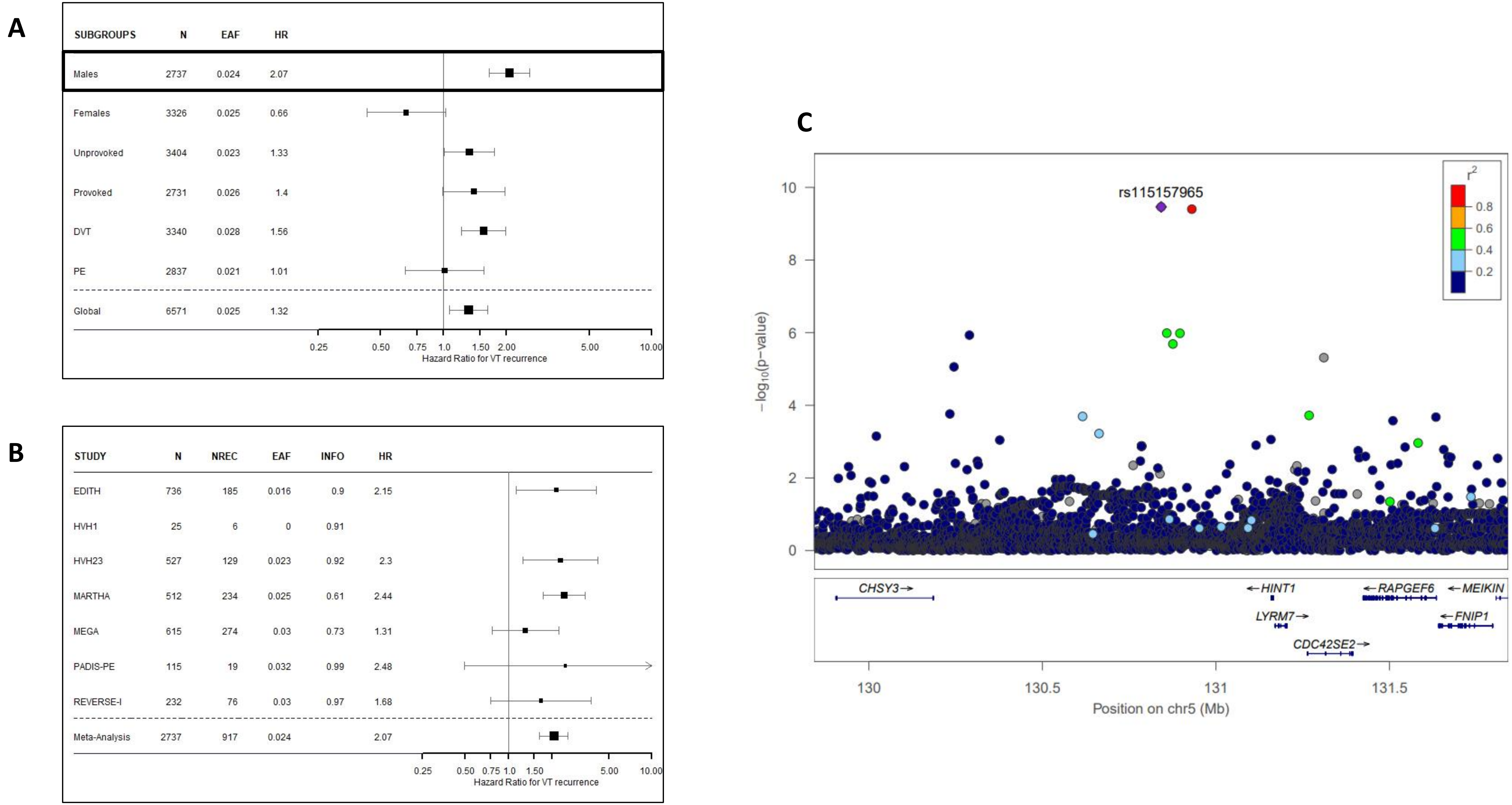
A
SUBGROUPS N EAF HR
Males 2737 0.024 2.07
Females 3326 0.025 0.66
Unprovoked 3404 0.023 1.33
Provoked 2731 0.026 1.4
DVT 3340 0.028 1.56
PE 2837 0.021 1.01
Global 6571 0.025 1.32
0.25 0.50 0.75 1.0 1.50 2.00 5.00 10.00
Hazard Ratio for VT recurrence
B
STUDY N NREC EAF INFO HR
EDITH 736 185 0.016 0.9 2.15
HVH1 25 6 0 0.91
HVH23 527 129 0.023 0.92 2.3
MARTHA 512 234 0.025 0.61 2.44
MEGA 615 274 0.03 0.73 1.31
PADIS-PE 115 19 0.032 0.99 2.48
REVERSE-I 232 76 0.03 0.97 1.68
Meta-Analysis 2737 917 0.024 2.07
0.25 0.50 0.75 1.0 1.50 5.00 10.00
Hazard Ratio for VT recurrence
C
rs115157965
r2
0.8
0.6
0.4
0.2
10
8
6
4
2
0
−log10(p−value)
CHSY3→
←HINT1
←RAPGEF6
←MEIKIN
LYRM7→
←FNIP1
CDC42SE2→
130
130.5
131
131.5
Position on chr5 (Mb)

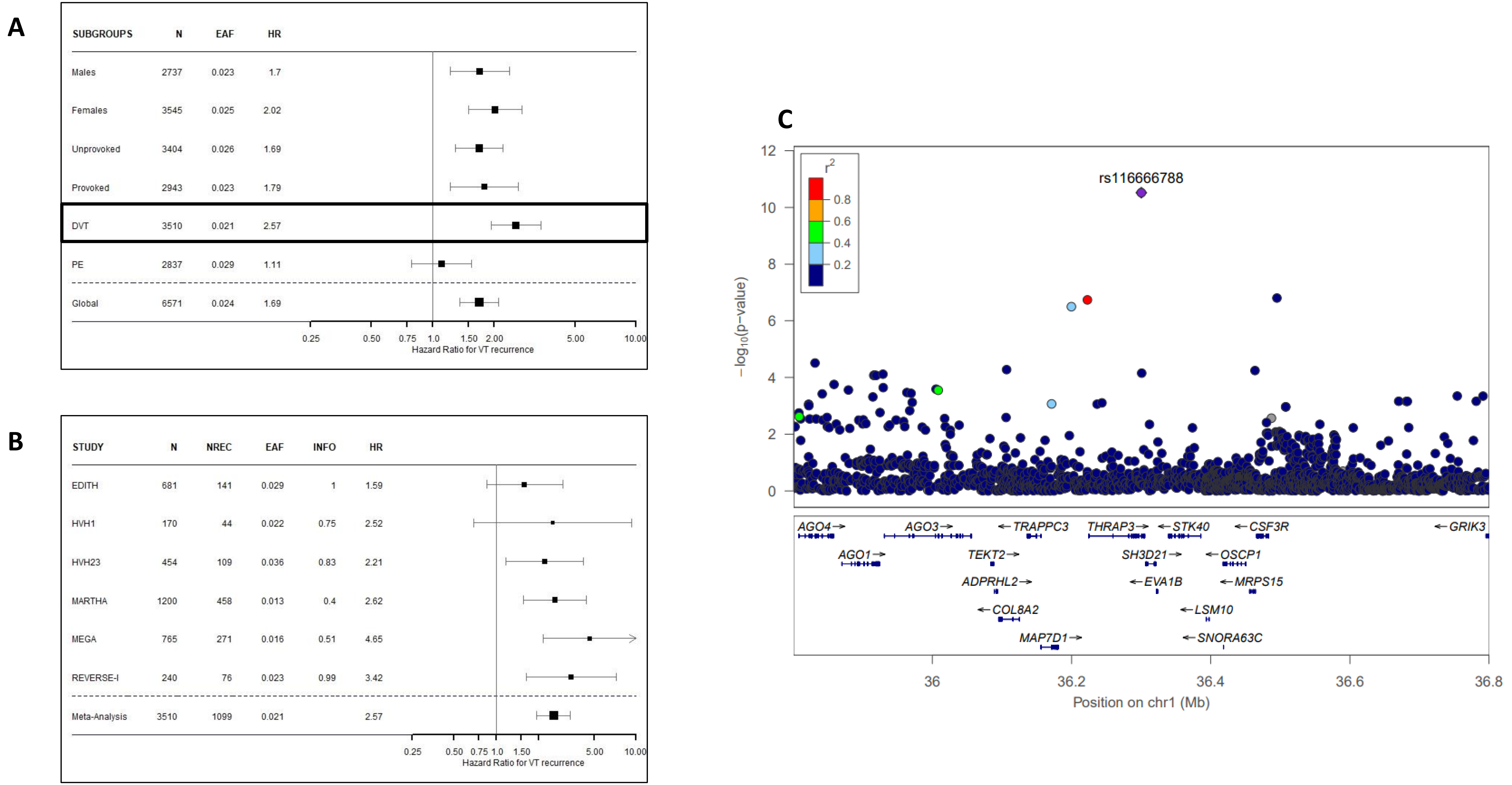
A
SUBGROUPS N EAF HR
Males 2737 0.023 1.7
Females 3545 0.025 2.02
Unprovoked 3404 0.026 1.69
Provoked 2943 0.023 1.79
DVT 3510 0.021 2.57
PE 2837 0.029 1.11
Global 6571 0.024 1.69
0.25 0.50 0.75 1.0 1.50 2.00 5.00 10.00
Hazard Ratio for VT recurrence
B
STUDY N NREC EAF INFO HR
EDITH 681 141 0.029 1 1.59
HVH1 170 44 0.022 0.75 2.52
HVH23 454 109 0.036 0.83 2.21
MARTHA 1200 458 0.013 0.4 2.62
MEGA 765 271 0.016 0.51 4.65
REVERSE-I 240 76 0.023 0.99 3.42
Meta-Analysis 3510 1099 0.021 2.57
0.25 0.50 0.75 1.0 1.50 5.00 10.00
Hazard Ratio for VT recurrence
C
rs116666788
r2
0.8
0.6
0.4
0.2
−log10(p−value)
0 2 4 6 8 10 12
AGO4→ AGO3→ ←TRAPPC3 THRAP3→ ←STK40 ←CSF3R ←GRIK3
AGO1→ TEKT2→ SH3D21→ ←OSCP1
ADPRHL2→ ←EVA1B ←MRPS15
←COL8A2 ←LSM10
MAP7D1→ ←SNORA63C
36 36.2 36.4 36.6 36.8
Position on chr1 (Mb)

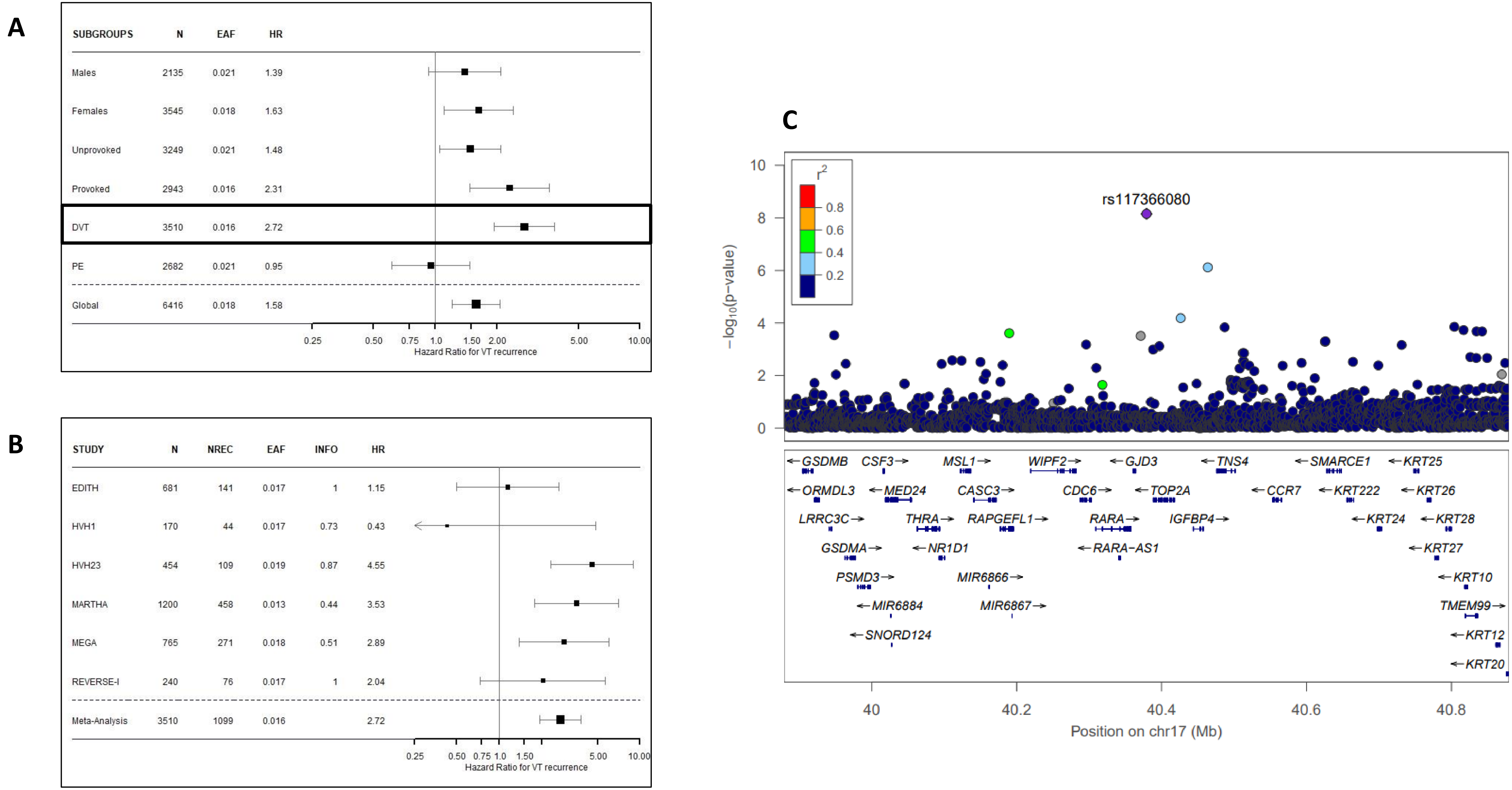
A
SUBGROUPS N EAF HR
Males 2135 0.021 1.39
Females 3545 0.018 1.63
Unprovoked 3249 0.021 1.48
Provoked 2943 0.016 2.31
DVT 3510 0.016 2.72
PE 2682 0.021 0.95
Global 6416 0.018 1.58
0.25 0.50 0.75 1.0 1.50 2.00 5.00 10.00
Hazard Ratio for VT recurrence
B
STUDY N NREC EAF INFO HR
EDITH 681 141 0.017 1 1.15
HVH1 170 44 0.017 0.73 0.43
HVH23 454 109 0.019 0.87 4.55
MARTHA 1200 458 0.013 0.44 3.53
MEGA 765 271 0.018 0.51 2.89
REVERSE-I 240 76 0.017 1 2.04
Meta-Analysis 3510 1099 0.016 2.72
0.25 0.50 0.75 1.0 1.50 5.00 10.00
Hazard Ratio for VT recurrence
C
rs117366080
r2
0.8
0.6
0.4
0.2
-log10(p-value)
GSDMB CSF3 MSL1 WIPF2 GJD3 TNS4 SMARCE1 KRT25
ORMDL3 MED24 CASC3 CDC6 TOP2A CCR7 KRT222 KRT26
LRRC3C THRA RAPGEFL1 RARA IGFBP4 KRT24 KRT28
GSDMA NR1D1 RARA-AS1 KRT27
PSMD3 MIR6866 KRT10
MIR6884 MIR6867 TMEM99
SNORD124 KRT12
KRT20
40 40.2 40.4 40.6 40.8
Position on chr17 (Mb)

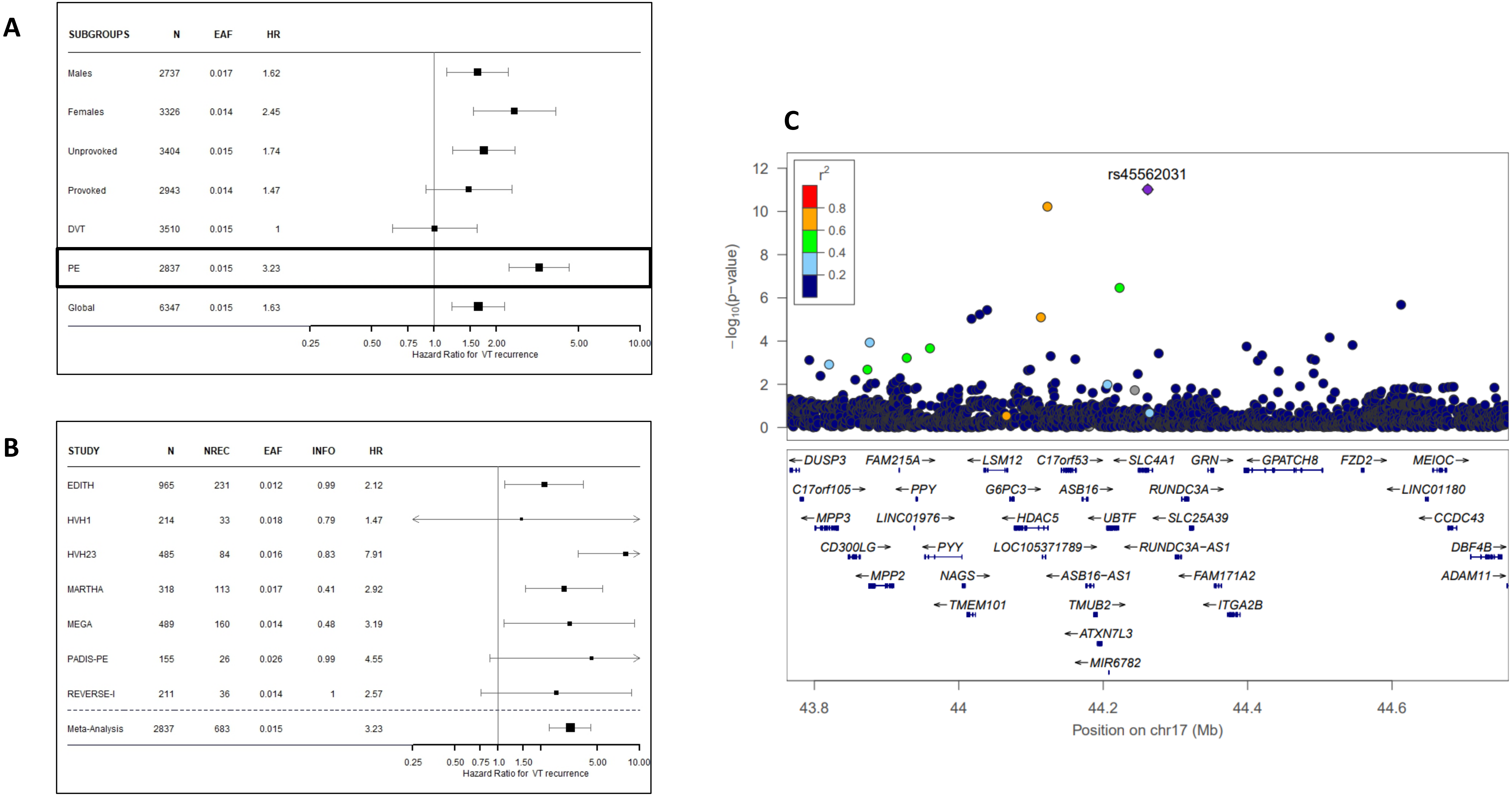
A
SUBGROUPS N EAF HR
Males 2737 0.017 1.62
Females 3326 0.014 2.45
Unprovoked 3404 0.015 1.74
Provoked 2943 0.014 1.47
DVT 3510 0.015 1
PE 2837 0.015 3.23
Global 6347 0.015 1.63
0.25 0.50 0.75 1.0 1.50 2.00 5.00 10.00
Hazard Ratio for VT recurrence
B
STUDY N NREC EAF INFO HR
EDITH 965 231 0.012 0.99 2.12
HVH1 214 33 0.018 0.79 1.47
HVH23 485 84 0.016 0.83 7.91
MARTHA 318 113 0.017 0.41 2.92
MEGA 489 160 0.014 0.48 3.19
PADIS-PE 155 26 0.026 0.99 4.55
REVERSE-I 211 36 0.014 1 2.57
Meta-Analysis 2837 683 0.015 3.23
0.25 0.50 0.75 1.0 1.50 5.00 10.00
Hazard Ratio for VT recurrence
C
rs45562031
$r^2$
0.8 0.6 0.4 0.2
$-\log_{10}$(p-value)
0 2 4 6 8 10 12
DUSP3 FAM215A LSM12 C17orf53 SLC4A1 GRN GPATCH8 FZD2 MEIOC
C17orf105 PPY G6PC3 ASB16 RUNDC3A LINC01180
MPP3 LINC01976 HDAC5 UBTF SLC25A39 CCDC43
CD300LG PYY LOC105371789 RUNDC3A-AS1 DBF4B
MPP2 NAGS ASB16-AS1 FAM171A2 ADAM11
TMEM101 TMUB2 ITGA2B
ATXN7L3
MIR6782
43.8 44 44.2 44.4 44.6
Position on chr17 (Mb)

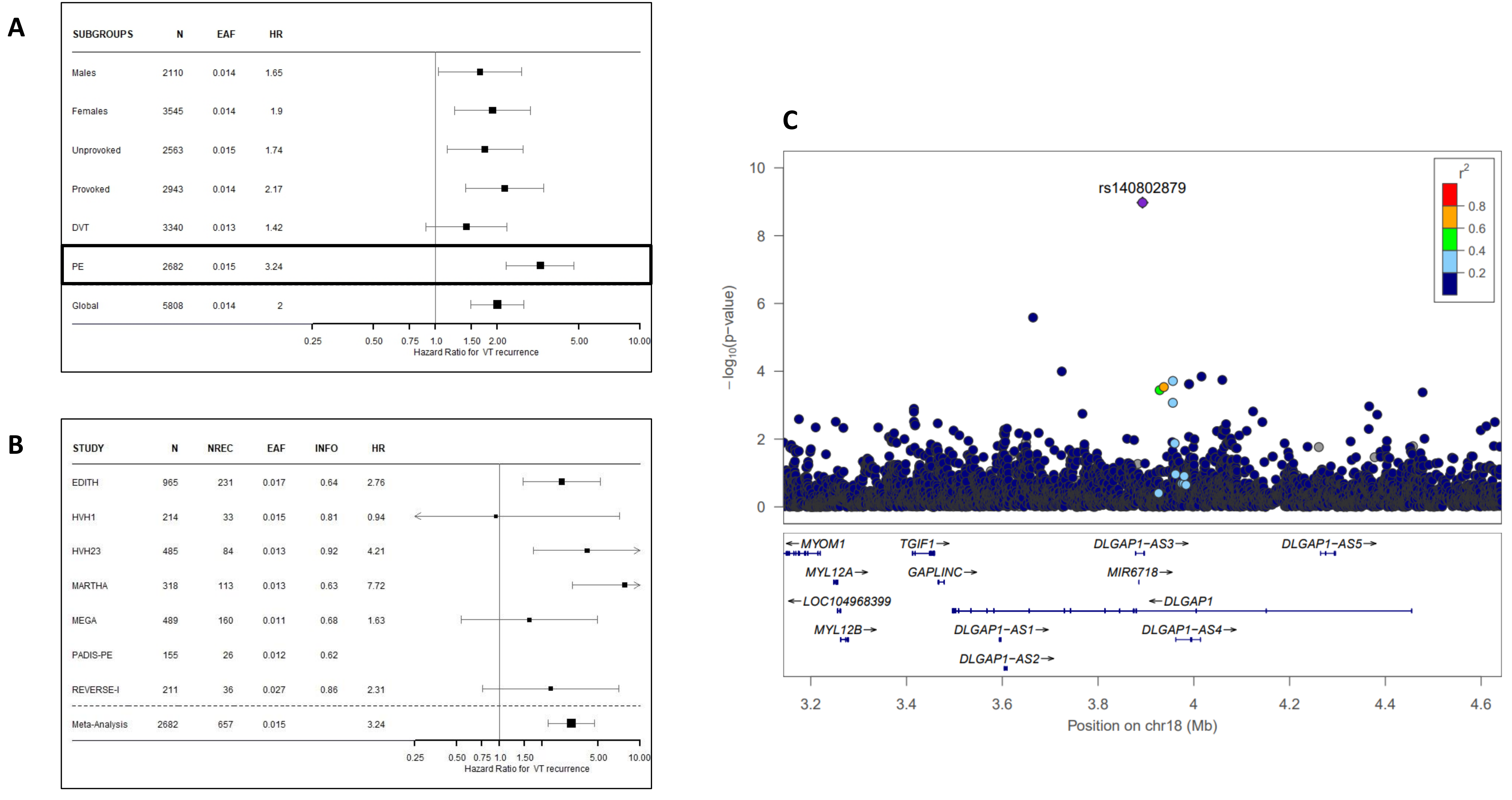
A
SUBGROUPS N EAF HR
Males 2110 0.014 1.65
Females 3545 0.014 1.9
Unprovoked 2563 0.015 1.74
Provoked 2943 0.014 2.17
DVT 3340 0.013 1.42
PE 2682 0.015 3.24
Global 5808 0.014 2
0.25 0.50 0.75 1.0 1.50 2.00 5.00 10.00
Hazard Ratio for VT recurrence
B
STUDY N NREC EAF INFO HR
EDITH 965 231 0.017 0.64 2.76
HVH1 214 33 0.015 0.81 0.94
HVH23 485 84 0.013 0.92 4.21
MARTHA 318 113 0.013 0.63 7.72
MEGA 489 160 0.011 0.68 1.63
PADIS-PE 155 26 0.012 0.62
REVERSE-I 211 36 0.027 0.86 2.31
Meta-Analysis 2682 657 0.015 3.24
0.25 0.50 0.75 1.0 1.50 5.00 10.00
Hazard Ratio for VT recurrence
C
rs140802879
r2
0.8
0.6
0.4
0.2
10
8
6
4
2
0
−log10(p−value)
←MYOM1
TGIF1→
DLGAP1−AS3→
DLGAP1−AS5→
MYL12A→
GAPLINC→
MIR6718→
←LOC104968399
←DLGAP1
MYL12B→
DLGAP1−AS1→
DLGAP1−AS4→
DLGAP1−AS2→
3.2 3.4 3.6 3.8 4 4.2 4.4 4.6
Position on chr18 (Mb)

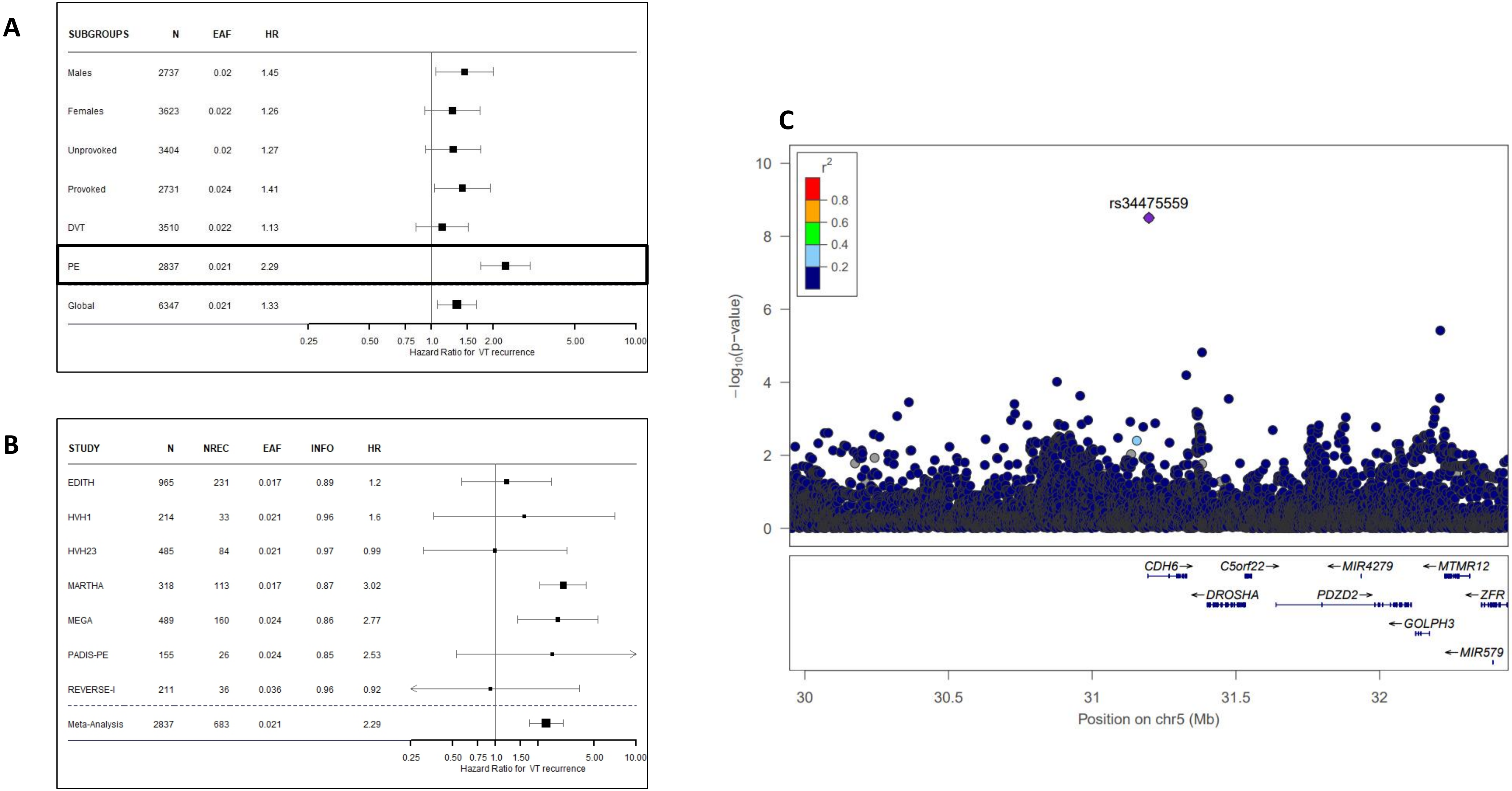
A
SUBGROUPS N EAF HR
Males 2737 0.02 1.45
Females 3623 0.022 1.26
Unprovoked 3404 0.02 1.27
Provoked 2731 0.024 1.41
DVT 3510 0.022 1.13
PE 2837 0.021 2.29
Global 6347 0.021 1.33
0.25 0.50 0.75 1.0 1.50 2.00 5.00 10.00
Hazard Ratio for VT recurrence
B
STUDY N NREC EAF INFO HR
EDITH 965 231 0.017 0.89 1.2
HVH1 214 33 0.021 0.96 1.6
HVH23 485 84 0.021 0.97 0.99
MARTHA 318 113 0.017 0.87 3.02
MEGA 489 160 0.024 0.86 2.77
PADIS-PE 155 26 0.024 0.85 2.53
REVERSE-I 211 36 0.036 0.96 0.92
Meta-Analysis 2837 683 0.021 2.29
0.25 0.50 0.75 1.0 1.50 5.00 10.00
Hazard Ratio for VT recurrence
C
r2
0.8
0.6
0.4
0.2
rs34475559
−log10(p−value)
10
8
6
4
2
0
CDH6→
C5orf22→
←MIR4279
←MTMR12
←DROSHA
PDZD2→
←ZFR
←GOLPH3
←MIR579
30
30.5
31
31.5
32
Position on chr5 (Mb)

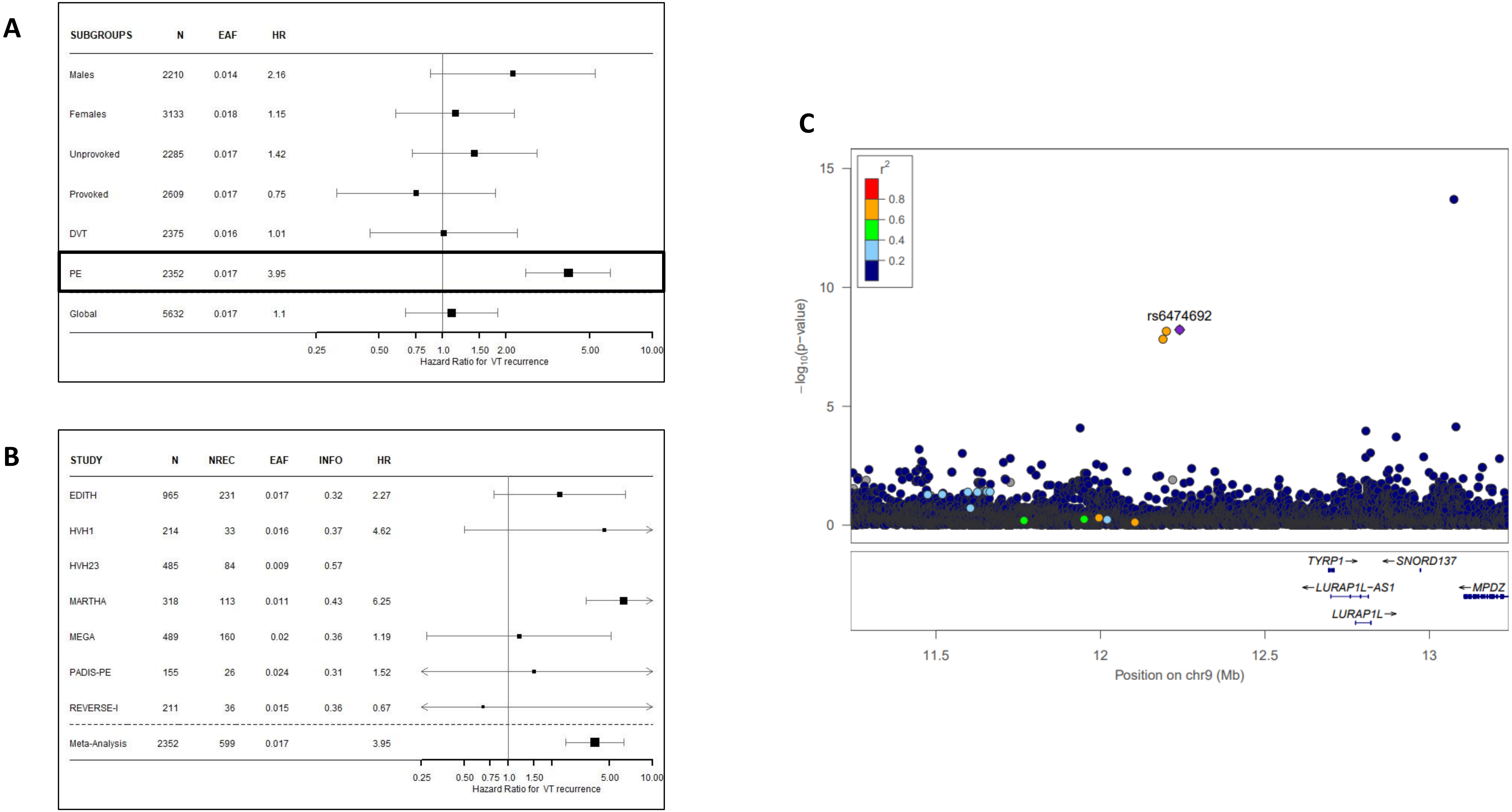
A
SUBGROUPS N EAF HR
Males 2210 0.014 2.16
Females 3133 0.018 1.15
Unprovoked 2285 0.017 1.42
Provoked 2609 0.017 0.75
DVT 2375 0.016 1.01
PE 2352 0.017 3.95
Global 5632 0.017 1.1
0.25 0.50 0.75 1.0 1.50 2.00 5.00 10.00
Hazard Ratio for VT recurrence
B
STUDY N NREC EAF INFO HR
EDITH 965 231 0.017 0.32 2.27
HVH1 214 33 0.016 0.37 4.62
HVH23 485 84 0.009 0.57
MARTHA 318 113 0.011 0.43 6.25
MEGA 489 160 0.02 0.36 1.19
PADIS-PE 155 26 0.024 0.31 1.52
REVERSE-I 211 36 0.015 0.36 0.67
Meta-Analysis 2352 599 0.017 3.95
0.25 0.50 0.75 1.0 1.50 5.00 10.00
Hazard Ratio for VT recurrence
C
r2
0.8
0.6
0.4
0.2
15
10
5
0
−log10(p−value)
rs6474692
TYRP1→
←SNORD137
←LURAP1L-AS1
←MPDZ
LURAP1L→
11.5
12
12.5
13
Position on chr9 (Mb)

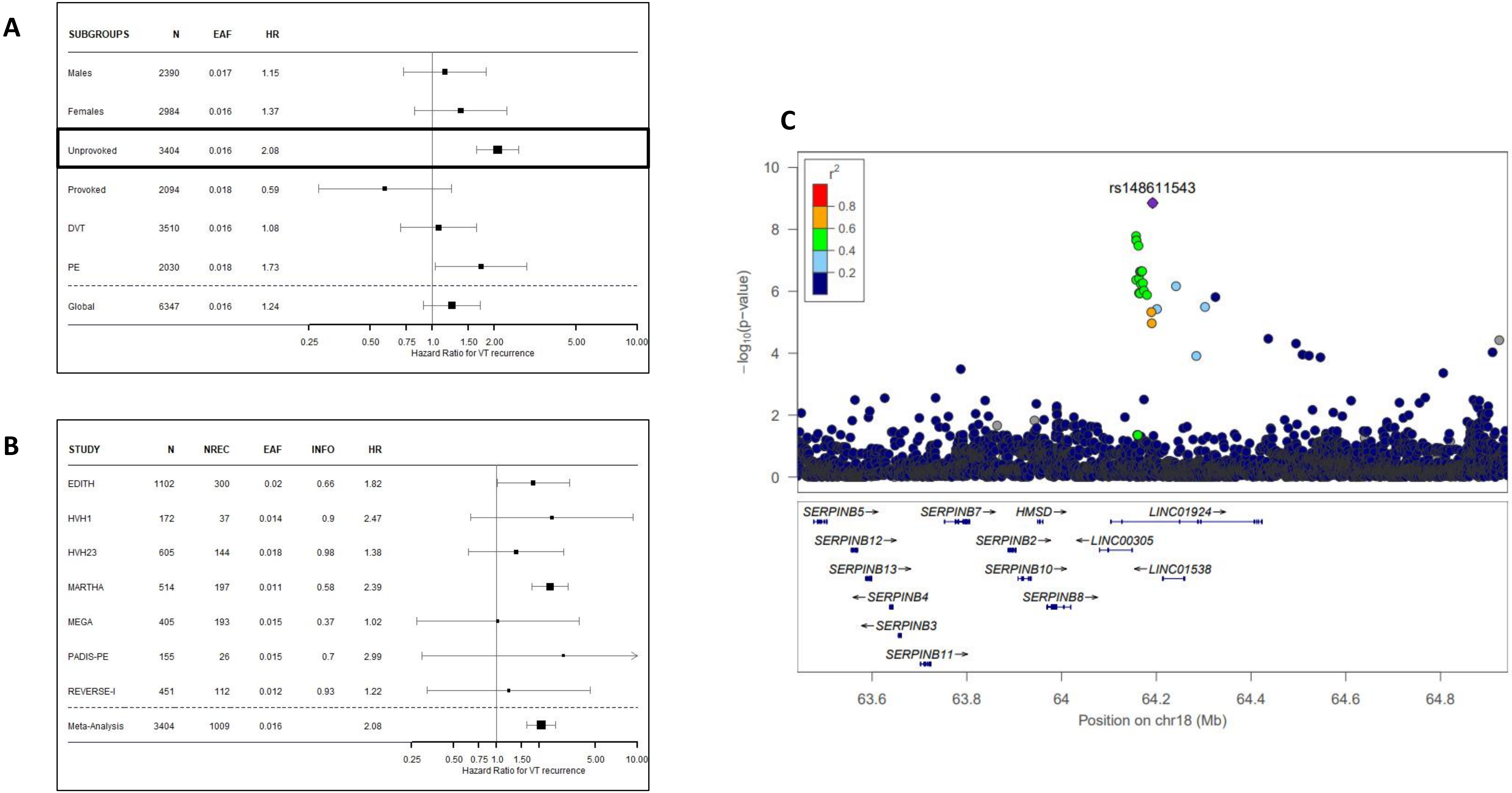
A
SUBGROUPS N EAF HR
Males 2390 0.017 1.15
Females 2984 0.016 1.37
Unprovoked 3404 0.016 2.08
Provoked 2094 0.018 0.59
DVT 3510 0.016 1.08
PE 2030 0.018 1.73
Global 6347 0.016 1.24
0.25 0.50 0.75 1.0 1.50 2.00 5.00 10.00
Hazard Ratio for VT recurrence
B
STUDY N NREC EAF INFO HR
EDITH 1102 300 0.02 0.66 1.82
HVH1 172 37 0.014 0.9 2.47
HVH23 605 144 0.018 0.98 1.38
MARTHA 514 197 0.011 0.58 2.39
MEGA 405 193 0.015 0.37 1.02
PADIS-PE 155 26 0.015 0.7 2.99
REVERSE-I 451 112 0.012 0.93 1.22
Meta-Analysis 3404 1009 0.016 2.08
0.25 0.50 0.75 1.0 1.50 5.00 10.00
Hazard Ratio for VT recurrence
C
r2
0.8
0.6
0.4
0.2
rs148611543
-log10(p-value)
0 2 4 6 8 10
SERPINB5→
SERPINB7→
HMSD→
LINC01924→
SERPINB12→
SERPINB2→
←LINC00305
SERPINB13→
SERPINB10→
←LINC01538
←SERPINB4
SERPINB8→
←SERPINB3
SERPINB11→
63.6 63.8 64 64.2 64.4 64.6 64.8
Position on chr18 (Mb)

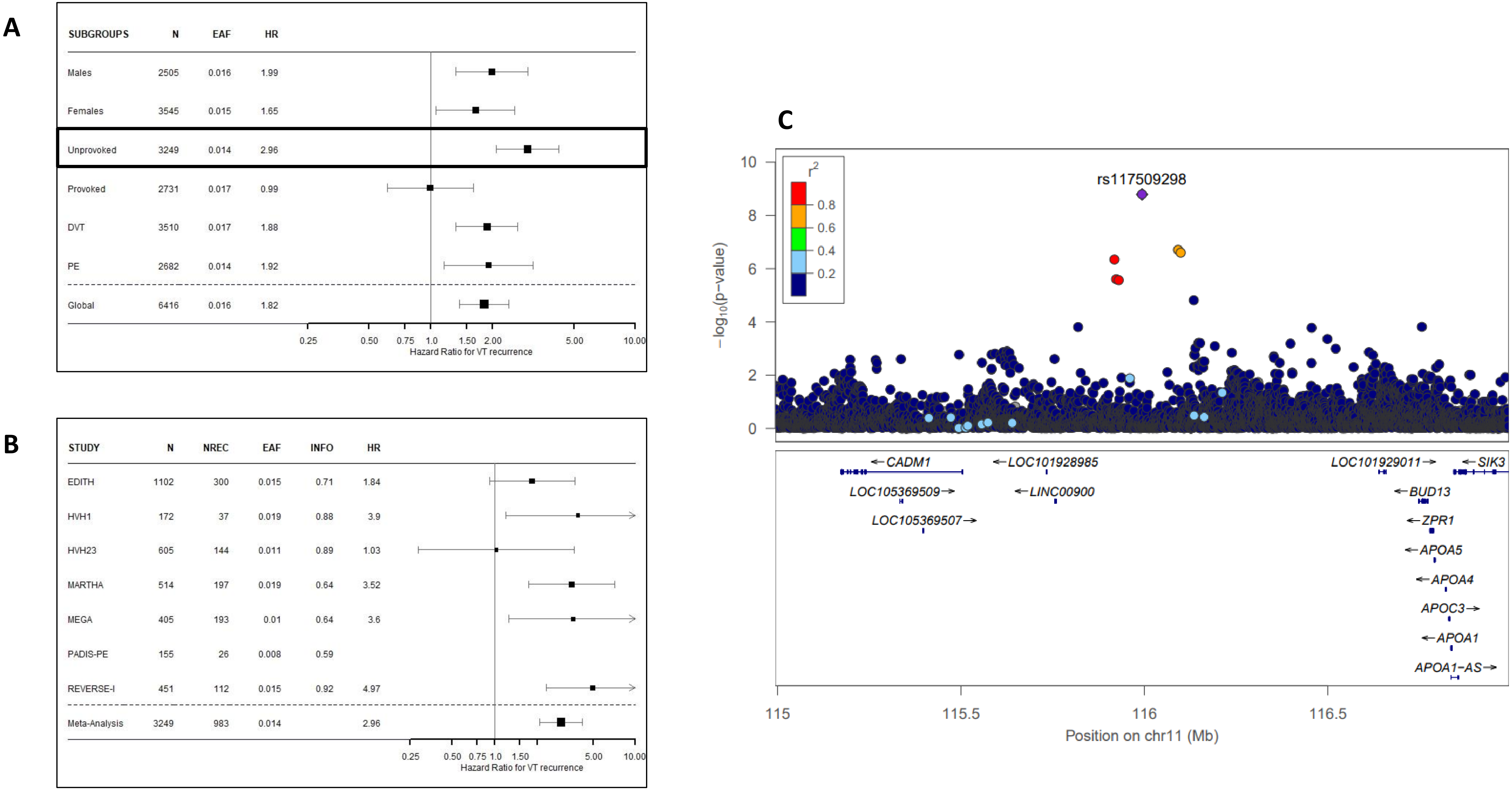
A
SUBGROUPS N EAF HR
Males 2505 0.016 1.99
Females 3545 0.015 1.65
Unprovoked 3249 0.014 2.96
Provoked 2731 0.017 0.99
DVT 3510 0.017 1.88
PE 2682 0.014 1.92
Global 6416 0.016 1.82
0.25 0.50 0.75 1.0 1.50 2.00 5.00 10.00
Hazard Ratio for VT recurrence
B
STUDY N NREC EAF INFO HR
EDITH 1102 300 0.015 0.71 1.84
HVH1 172 37 0.019 0.88 3.9
HVH23 605 144 0.011 0.89 1.03
MARTHA 514 197 0.019 0.64 3.52
MEGA 405 193 0.01 0.64 3.6
PADIS-PE 155 26 0.008 0.59
REVERSE-I 451 112 0.015 0.92 4.97
Meta-Analysis 3249 983 0.014 2.96
0.25 0.50 0.75 1.0 1.50 5.00 10.00
Hazard Ratio for VT recurrence
C
r2
0.8
0.6
0.4
0.2
rs117509298
−log10(p-value)
10
8
6
4
2
0
←CADM1
LOC105369509→
LOC105369507→
←LOC101928985
←LINC00900
LOC101929011→
←SIK3
←BUD13
←ZPR1
←APOA5
←APOA4
APOC3→
←APOA1
APOA1-AS→
115
115.5
116
116.5
Position on chr11 (Mb)

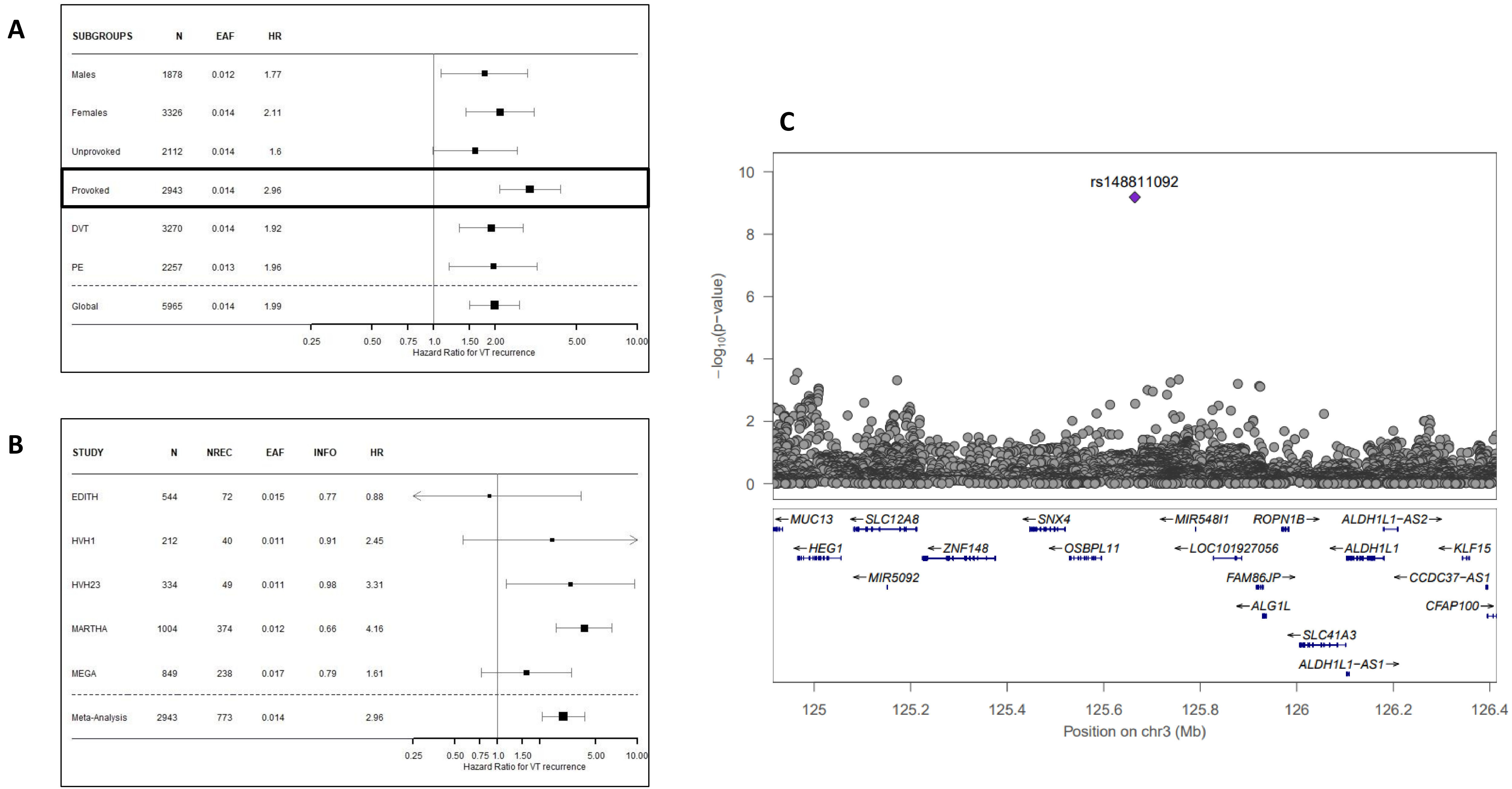
A
SUBGROUPS N EAF HR
Males 1878 0.012 1.77
Females 3326 0.014 2.11
Unprovoked 2112 0.014 1.6
Provoked 2943 0.014 2.96
DVT 3270 0.014 1.92
PE 2257 0.013 1.96
Global 5965 0.014 1.99
0.25 0.50 0.75 1.0 1.50 2.00 5.00 10.00
Hazard Ratio for VT recurrence
B
STUDY N NREC EAF INFO HR
EDITH 544 72 0.015 0.77 0.88
HVH1 212 40 0.011 0.91 2.45
HVH23 334 49 0.011 0.98 3.31
MARTHA 1004 374 0.012 0.66 4.16
MEGA 849 238 0.017 0.79 1.61
Meta-Analysis 2943 773 0.014 2.96
0.25 0.50 0.75 1.0 1.50 5.00 10.00
Hazard Ratio for VT recurrence
C
rs148811092
−log10(p−value)
0 2 4 6 8 10
MUC13
SLC12A8
SNX4
MIR548I1
ROPN1B
ALDH1L1−AS2
HEG1
ZNF148
OSBPL11
LOC101927056
ALDH1L1
KLF15
MIR5092
FAM86JP
CCDC37−AS1
ALG1L
CFAP100
SLC41A3
ALDH1L1−AS1
125 125.2 125.4 125.6 125.8 126 126.2 126.4
Position on chr3 (Mb)

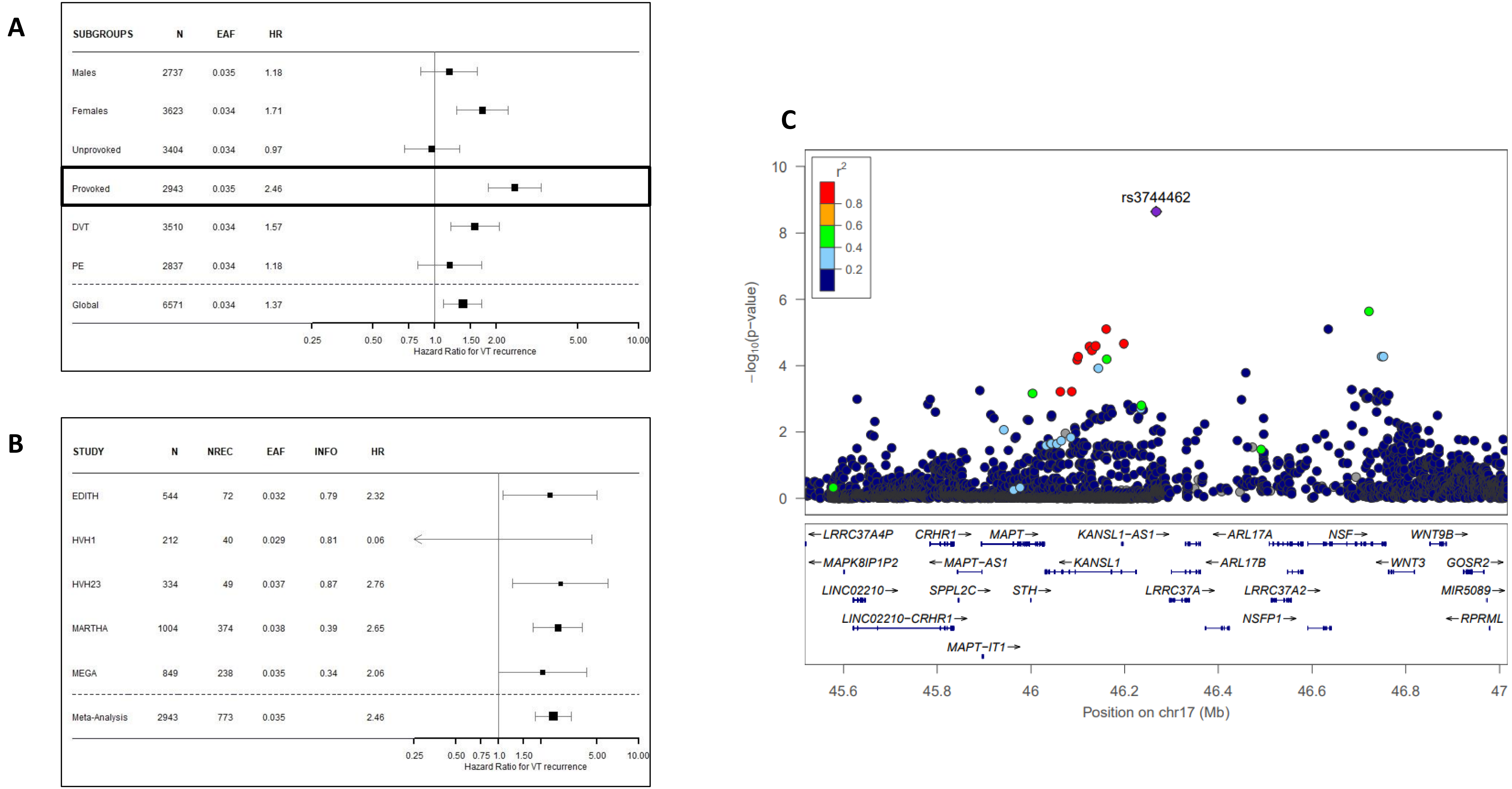
A
SUBGROUPS N EAF HR
Males 2737 0.035 1.18
Females 3623 0.034 1.71
Unprovoked 3404 0.034 0.97
Provoked 2943 0.035 2.46
DVT 3510 0.034 1.57
PE 2837 0.034 1.18
Global 6571 0.034 1.37
0.25 0.50 0.75 1.0 1.50 2.00 5.00 10.00
Hazard Ratio for VT recurrence
B
STUDY N NREC EAF INFO HR
EDITH 544 72 0.032 0.79 2.32
HVH1 212 40 0.029 0.81 0.06
HVH23 334 49 0.037 0.87 2.76
MARTHA 1004 374 0.038 0.39 2.65
MEGA 849 238 0.035 0.34 2.06
Meta-Analysis 2943 773 0.035 2.46
0.25 0.50 0.75 1.0 1.50 5.00 10.00
Hazard Ratio for VT recurrence
C
r²
0.8
0.6
0.4
0.2
rs3744462
−log10(p-value)
0 2 4 6 8 10
←LRRC37A4P CRHR1→ MAPT→ KANSL1-AS1→ ←ARL17A NSF→ WNT9B→
←MAPK8IP1P2 ←MAPT-AS1 ←KANSL1 ←ARL17B ←WNT3 GOSR2→
LINC02210→ SPPL2C→ STH→ LRRC37A→ LRRC37A2→ MIR5089→
LINC02210-CRHR1→ NSFP1→ ←RPRML
MAPT-IT1→
45.6 45.8 46 46.2 46.4 46.6 46.8 47
Position on chr17 (Mb)

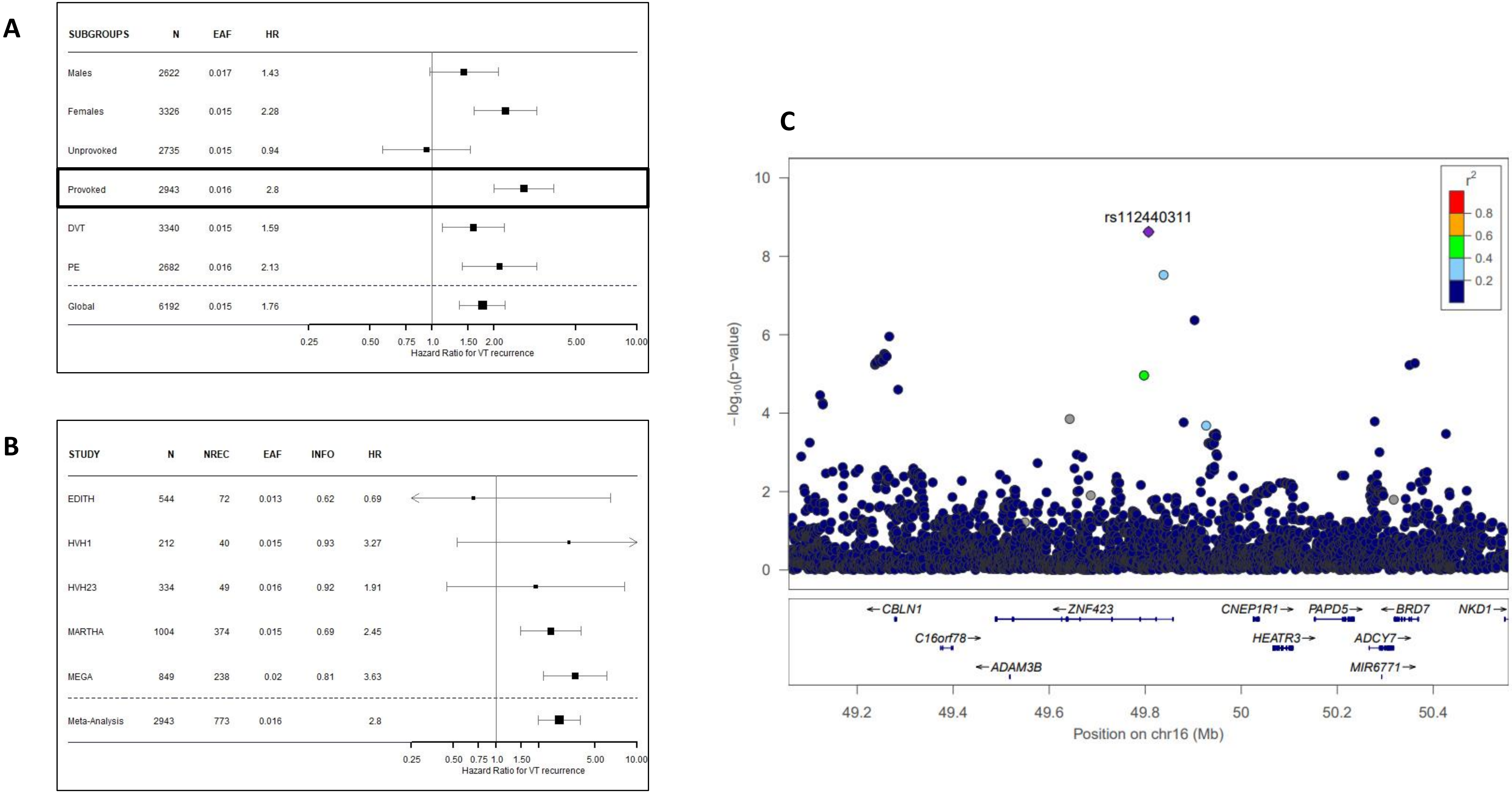
A
SUBGROUPS N EAF HR
Males 2622 0.017 1.43
Females 3326 0.015 2.28
Unprovoked 2735 0.015 0.94
Provoked 2943 0.016 2.8
DVT 3340 0.015 1.59
PE 2682 0.016 2.13
Global 6192 0.015 1.76
0.25 0.50 0.75 1.0 1.50 2.00 5.00 10.00
Hazard Ratio for VT recurrence
B
STUDY N NREC EAF INFO HR
EDITH 544 72 0.013 0.62 0.69
HVH1 212 40 0.015 0.93 3.27
HVH23 334 49 0.016 0.92 1.91
MARTHA 1004 374 0.015 0.69 2.45
MEGA 849 238 0.02 0.81 3.63
Meta-Analysis 2943 773 0.016 2.8
0.25 0.50 0.75 1.0 1.50 5.00 10.00
Hazard Ratio for VT recurrence
C
rs112440311
r2
0.8
0.6
0.4
0.2
−log10(p-value)
0 2 4 6 8 10
CBLN1
ZNF423
CNEP1R1
PAPD5
BRD7
NKD1
C16orf78
HEATR3
ADCY7
ADAM3B
MIR6771
49.2 49.4 49.6 49.8 50 50.2 50.4
Position on chr16 (Mb)

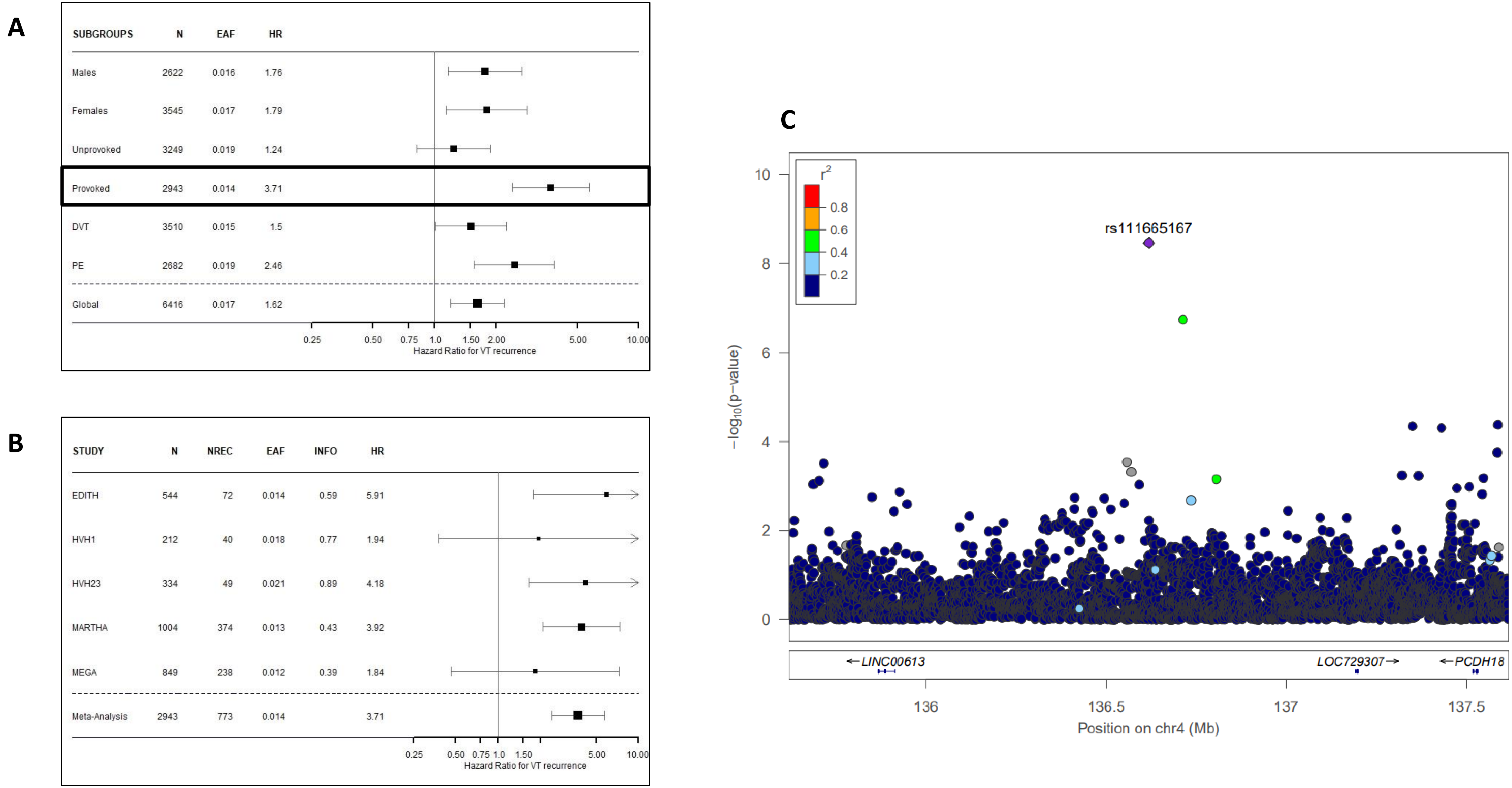
A
SUBGROUPS N EAF HR
Males 2622 0.016 1.76
Females 3545 0.017 1.79
Unprovoked 3249 0.019 1.24
Provoked 2943 0.014 3.71
DVT 3510 0.015 1.5
PE 2682 0.019 2.46
Global 6416 0.017 1.62
0.25 0.50 0.75 1.0 1.50 2.00 5.00 10.00
Hazard Ratio for VT recurrence
B
STUDY N NREC EAF INFO HR
EDITH 544 72 0.014 0.59 5.91
HVH1 212 40 0.018 0.77 1.94
HVH23 334 49 0.021 0.89 4.18
MARTHA 1004 374 0.013 0.43 3.92
MEGA 849 238 0.012 0.39 1.84
Meta-Analysis 2943 773 0.014 3.71
0.25 0.50 0.75 1.0 1.50 5.00 10.00
Hazard Ratio for VT recurrence
C
r2
0.8
0.6
0.4
0.2
rs111665167
−log10(p−value)
0 2 4 6 8 10
LINC00613
LOC729307
PCDH18
136 136.5 137 137.5
Position on chr4 (Mb)

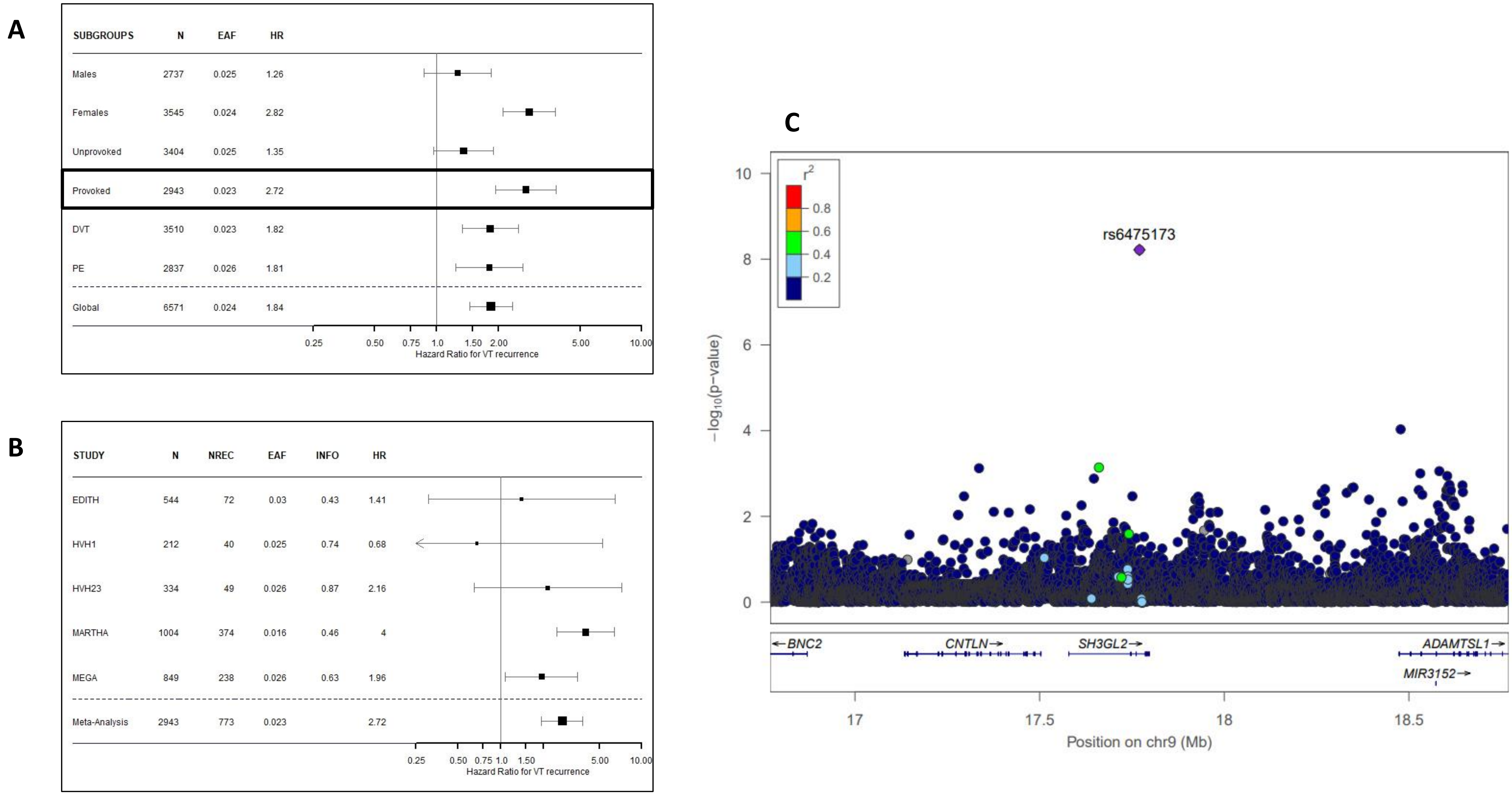
A
SUBGROUPS N EAF HR
Males 2737 0.025 1.26
Females 3545 0.024 2.82
Unprovoked 3404 0.025 1.35
Provoked 2943 0.023 2.72
DVT 3510 0.023 1.82
PE 2837 0.026 1.81
Global 6571 0.024 1.84
0.25 0.50 0.75 1.0 1.50 2.00 5.00 10.00
Hazard Ratio for VT recurrence
B
STUDY N NREC EAF INFO HR
EDITH 544 72 0.03 0.43 1.41
HVH1 212 40 0.025 0.74 0.68
HVH23 334 49 0.026 0.87 2.16
MARTHA 1004 374 0.016 0.46 4
MEGA 849 238 0.026 0.63 1.96
Meta-Analysis 2943 773 0.023 2.72
0.25 0.50 0.75 1.0 1.50 5.00 10.00
Hazard Ratio for VT recurrence
C
$r^2$
0.8
0.6
0.4
0.2
rs6475173
$-\log_{10}$(p-value)
0
2
4
6
8
10
←BNC2
CNTLN→
SH3GL2→
ADAMTSL1→
MIR3152→
17
17.5
18
18.5
Position on chr9 (Mb)

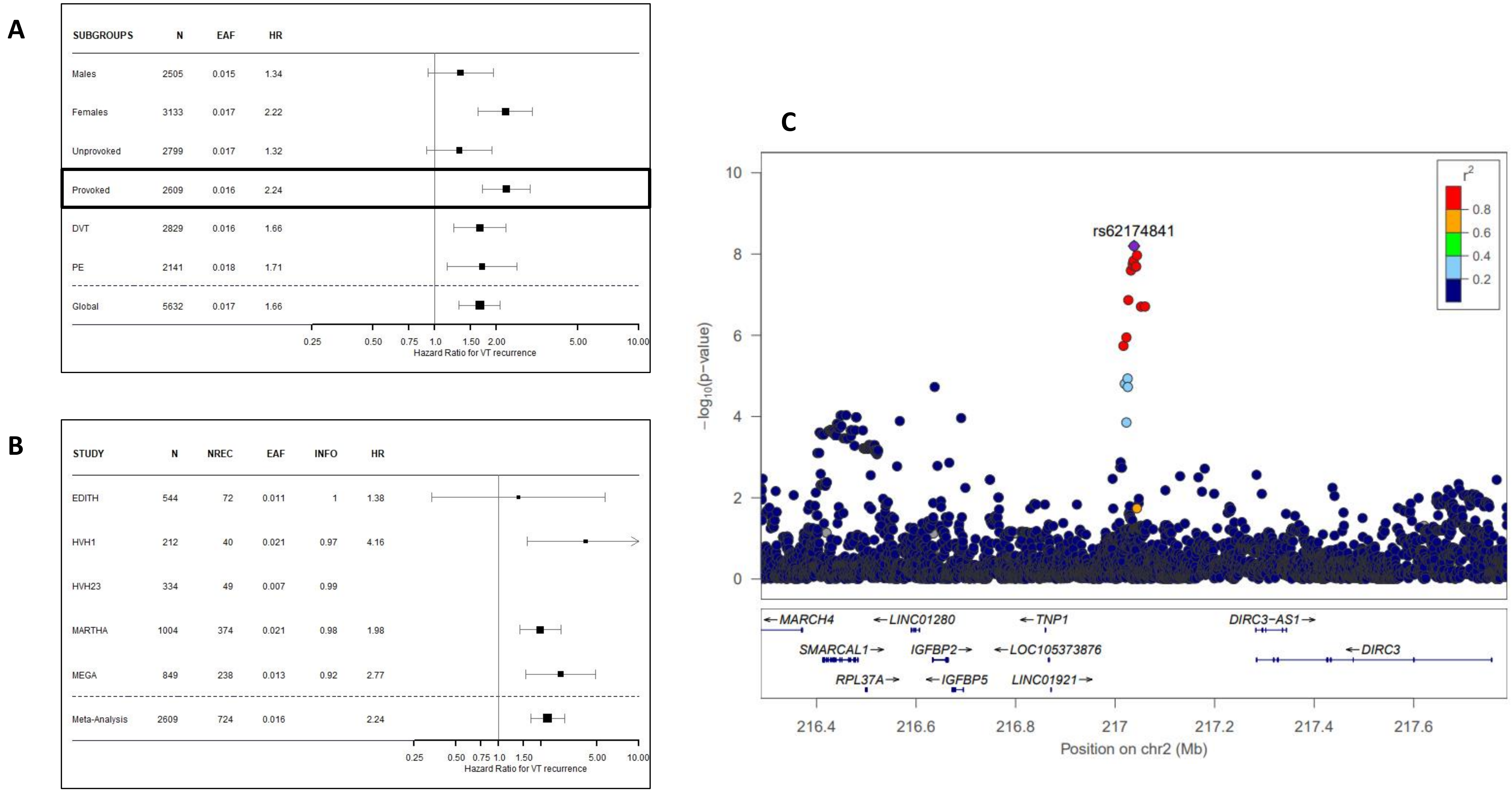
A
SUBGROUPS N EAF HR
Males 2505 0.015 1.34
Females 3133 0.017 2.22
Unprovoked 2799 0.017 1.32
Provoked 2609 0.016 2.24
DVT 2829 0.016 1.66
PE 2141 0.018 1.71
Global 5632 0.017 1.66
0.25 0.50 0.75 1.0 1.50 2.00 5.00 10.00
Hazard Ratio for VT recurrence
B
STUDY N NREC EAF INFO HR
EDITH 544 72 0.011 1 1.38
HVH1 212 40 0.021 0.97 4.16
HVH23 334 49 0.007 0.99
MARTHA 1004 374 0.021 0.98 1.98
MEGA 849 238 0.013 0.92 2.77
Meta-Analysis 2609 724 0.016 2.24
0.25 0.50 0.75 1.0 1.50 5.00 10.00
Hazard Ratio for VT recurrence
C
rs62174841
r2
0.8
0.6
0.4
0.2
−log10(p−value)
0 2 4 6 8 10
←MARCH4
←LINC01280
←TNP1
DIRC3−AS1→
SMARCAL1→
IGFBP2→
←LOC105373876
←DIRC3
RPL37A→
←IGFBP5
LINC01921→
216.4 216.6 216.8 217 217.2 217.4 217.6
Position on chr2 (Mb)

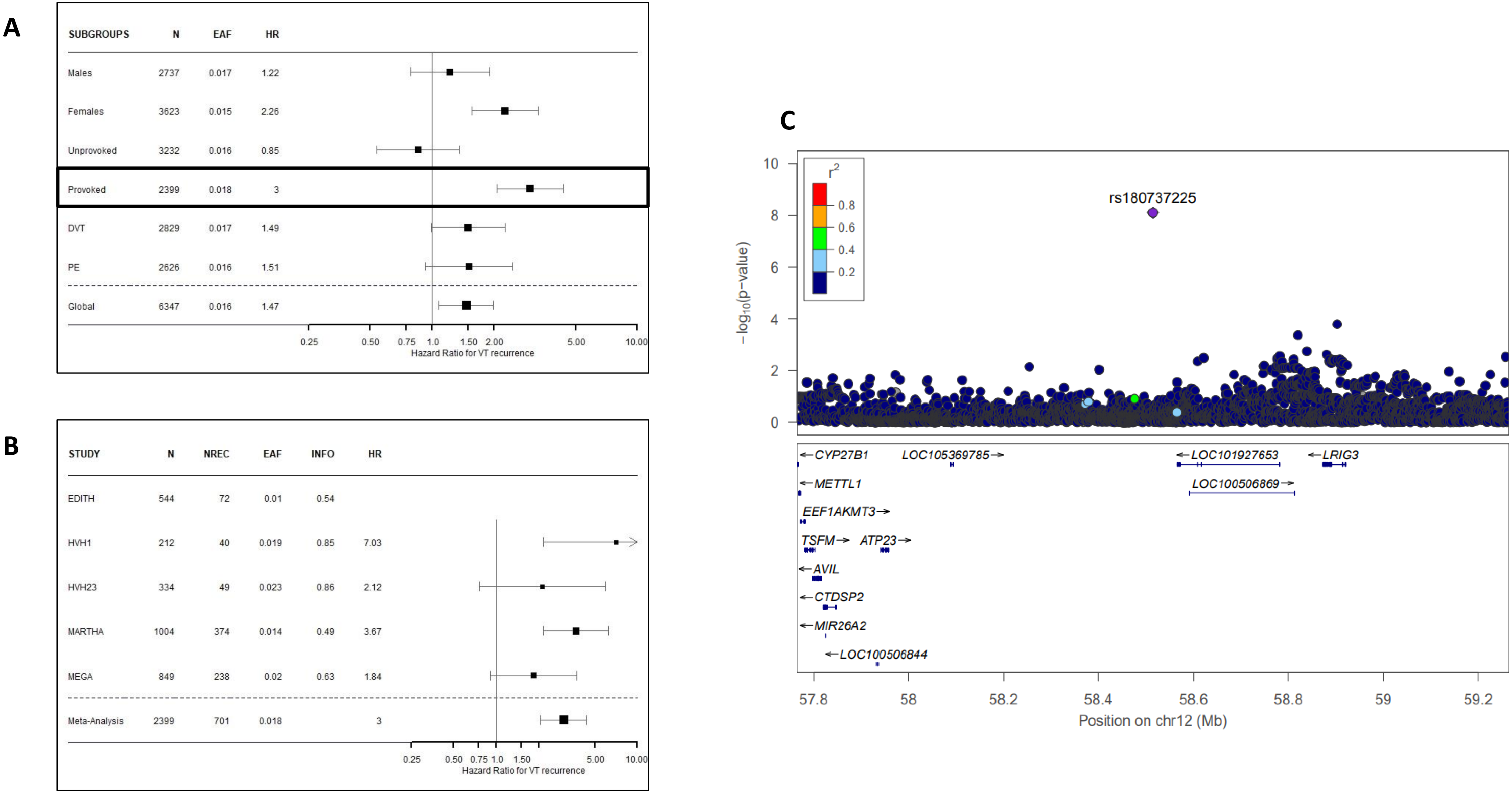
A
SUBGROUPS N EAF HR
Males 2737 0.017 1.22
Females 3623 0.015 2.26
Unprovoked 3232 0.016 0.85
Provoked 2399 0.018 3
DVT 2829 0.017 1.49
PE 2626 0.016 1.51
Global 6347 0.016 1.47
0.25 0.50 0.75 1.0 1.50 2.00 5.00 10.00
Hazard Ratio for VT recurrence
B
STUDY N NREC EAF INFO HR
EDITH 544 72 0.01 0.54
HVH1 212 40 0.019 0.85 7.03
HVH23 334 49 0.023 0.86 2.12
MARTHA 1004 374 0.014 0.49 3.67
MEGA 849 238 0.02 0.63 1.84
Meta-Analysis 2399 701 0.018 3
0.25 0.50 0.75 1.0 1.50 5.00 10.00
Hazard Ratio for VT recurrence
C
r2
0.8
0.6
0.4
0.2
rs180737225
10
8
6
4
2
0
-log10(p-value)
CYP27B1
LOC105369785
LOC101927653
LRIG3
METTL1
LOC100506869
EEF1AKMT3
TSFM
ATP23
AVIL
CTDSP2
MIR26A2
LOC100506844
57.8 58 58.2 58.4 58.6 58.8 59 59.2
Position on chr12 (Mb)

| STUDY | N | NREC | EAF | INFO | HR |
|---|---|---|---|---|---|
| EDITH | 1646 | 372 | 0.065 | 0.99 | 1.14 |
| FHS | 224 | 34 | 0.044 | 0.98 | 0.69 |
| HVH1 | 384 | 77 | 0.058 | 0.91 | 2.19 |
| HVH23 | 939 | 193 | 0.066 | 0.97 | 1.1 |
| MARTHA | 1518 | 571 | 0.105 | 0.82 | 0.75 |
| MEGA | 1254 | 431 | 0.084 | 0.89 | 1.61 |
| PADIS-PE | 155 | 26 | 0.074 | 0.99 | 0.72 |
| REVERSE-I | 451 | 112 | 0.077 | 0.99 | 1.38 |
| Meta-Analysis | 6571 | 1816 | 0.078 | | 1.14 |

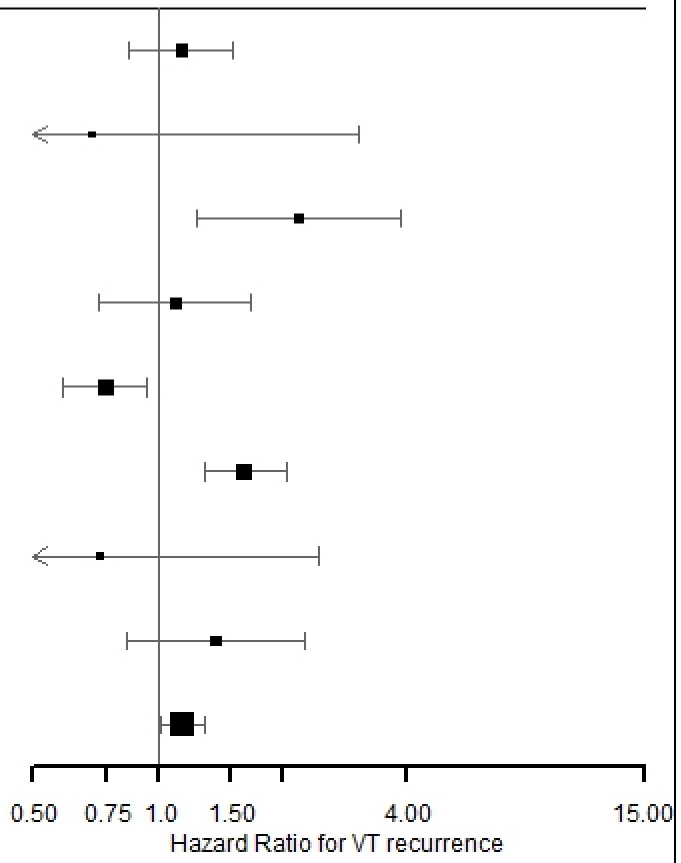

| STUDY | N | NREC | EAF | INFO | HR |
|---|---|---|---|---|---|
| EDITH | 1646 | 372 | 0.944 | 0.59 | 1.06 |
| FHS | 224 | 34 | 0.954 | 0.76 | 2.8 |
| HVH1 | 384 | 77 | 0.955 | 0.64 | 0.45 |
| HVH23 | 939 | 193 | 0.966 | 0.75 | 0.83 |
| MARTHA | 1518 | 571 | 0.909 | 0.35 | 0.86 |
| MEGA | 1254 | 431 | 0.886 | 0.29 | |
| PADIS-PE | 155 | 26 | 0.934 | 0.72 | 0.48 |
| REVERSE-I | 451 | 112 | 0.955 | 0.64 | 1.78 |
| Meta-Analysis | 5317 | 1385 | 0.94 | | 0.89 |

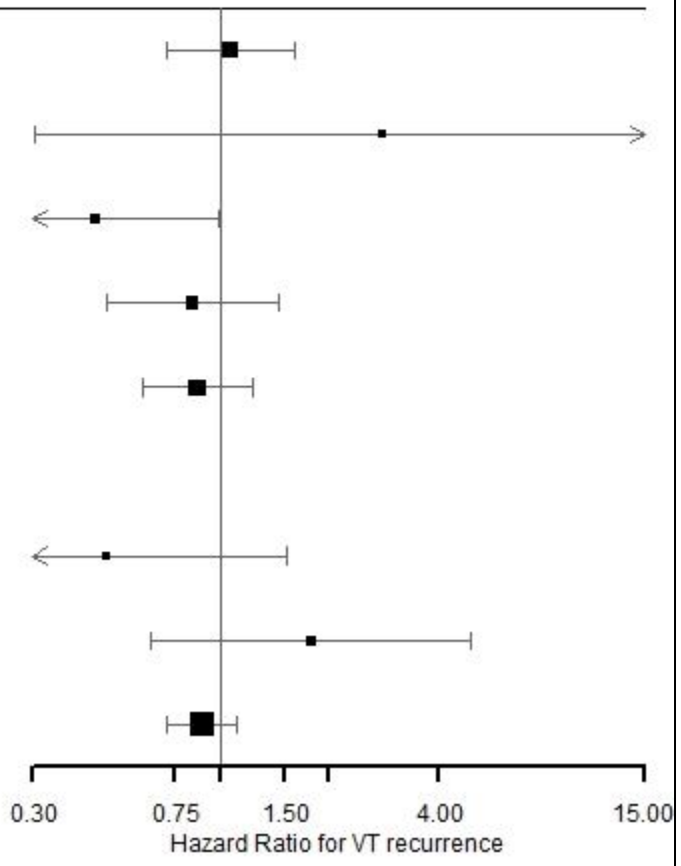
