## Supplementary Material for "Genomic Landscape of Thrombosis Recurrence Risk Across Venous Thromboembolism Subtypes"

### Participants and samples

#### 3C Study

The **Three-City Study (3C Study)** is a population-based cohort that included N=9,294 participants aged over 65 between 1999 and 2000 in three French cities (Bordeaux, Dijon and Montpellier)^1^. Participants of 3C-Dijon (N=4,931) were genotyped with an Illumina Human 610-Quad BeadChip array at the Centre National de Génotypage (Evry, France) and the imputation was performed on 1000 Genomes Phase 1 version 3. Detailed of genetic quality control procedures are described elsewhere^2^.

In a subsample of N=1,100 participants from Dijon, proteomic profiling was performed with Olink Explore 3072 panel using Proximity Extension Assay (PEA) technology on EDTA plasma tube of more than 500 μL not thawed/refrozen using the PEA technology following the manufacturer’s protocol^3^. After quality control of blood samples obtained at inclusion of the participants, one individual was removed and profiling was performed at the McGill Genome Center (Montreal, Canada) for 1,099 participants. Pre-processing of the proteomic data included plate-based normalization and QC checks based on appropriate Olink protocols. Data were transformed and normalized to Olink’s Normalized Protein eXpression (NPX) values, relative protein quantification unit in a logarithmic base 2 scale. Twelve samples were removed after principal component analysis of all proteins because they are found to deviate of more than 5 standard deviations from the mean. Besides, three proteins were removed as more than 50% of NPX values were below the protein's detection limit. Finally, proteomics data targeting 2,938 plasma proteins were available for 1,087 participants from the 3C study. For all the 1,087 participants, genetic data were also available.

#### EDITH

The “**Etude des Déterminants et Interactions de la Thrombose veineuse**” **(EDITH)** study includes 3,169 incident VT cases aged over 18, enrolled between 2000 and 2019 in Brest (France)^4^. Inclusion of patients is still in progress and those already included are regularly followed up. Patients have been typed by a high-density genotyping array, and 2,989 participants passed the standard quality controls. In the present study, patients with at least one of the following criteria were removed: uncertainty about VT documentation, history of cancer, homozygosity for FV Leiden or Factor II 20210A, personal history of VT. Finally, 1,646 VT patients were left for the VT recurrence analysis, including 372 VT recurrences. The EDITH study was approved by Brest University Hospital scientific and ethics board, in accordance with the Declaration of Helsinki. Written consent for the study and DNA analysis was obtained from all patients.

#### FHS

The **Framingham Heart Study (FHS)** was started in 1948 with 5,209 randomly ascertained participants from Framingham, Massachusetts, US, who had undergone biannual examinations to investigate cardiovascular disease and its risk factors. In 1971, the Offspring cohort (comprising 5,124 children of the original cohort and the children's spouses) and in 2002, the Third Generation (consisting of 4,095 children of the Offspring cohort) were recruited. The methods of recruitment and data collection for the Offspring and Third Generation cohorts have been described extensively elsewhere^5,6^. VT was systematically adjudicated from the baseline January 1, 1995 to the end of follow up on December 31, 2019. Death and VT cases prior to the baseline were excluded from the analysis. Following the analysis plan, the final sample size for analysis was 224 with 34 VT recurrent cases. Among the 224 samples, 30, 4, 59, and 131 were from the Original, the New Offspring Spouse, the Offspring and the Third Generation cohorts, respectively. The Institutional Review Board of Boston University Medical Center approved the study protocol, and all participants provided written informed consent.

#### HVH

This population-based, incident VT inception cohort was formed of cases from the **Heart and Vascular Health (HVH)** case-control study of VT, details of which have been published previously^7–9^. The HVH study was set within Group Health Cooperative (GHC), now Kaiser Permanente Washington (KPWA), which serves Western Washington State^9^. GHC was an integrated healthcare delivery system in which members received virtually all their care within the unified GHC system. The HVH study has been approved by the GHC/KPWA institutional review board. The VT cases were combined from 2 studies and included men aged 30-89 years and women aged 18-89 years who experienced an incident VT from January 2002 through December 2010. All subjects were members of GHC at the time of their incident VT. Using an ICD-9 code screen, trained, medical record abstractors reviewed the complete health record (outpatient, inpatient, provider notes, laboratory measures, pharmacy records) to identify eligible incident events, both provoked and unprovoked, through 2014 ^7^. Importantly, this study was based on clinical diagnoses and care delivered by GHC providers so asymptomatic events not detected by the provider could not be included in the study data. Qualifying events could not be catheter-related and required confirmatory imaging including Doppler or duplex ultrasound, computed tomography, pulmonary angiography, or ventilation-perfusion scan, or supporting clinical evidence of an incident VT with a physician diagnosis^7,8^. Ninety-eight percent of qualifying events included imaging. The final sample for the current project was composed of 1,323 VT cases, including 270 VT recurrences.

#### MARTHA

The **“MARseille THrombosis Association” (MARTHA)** study includes 2,837 unrelated VT patients who had a consultation visit at the Thrombophilia centre of La Timone Hospital in Marseille (France) between 1994 and 2008 ^10,11^. All patients with at least one documented VT and free of any chronic conditions and of any well characterised genetic risk factors including homozygosity for FV Leiden or Factor II 20210A, protein C, protein S and antithrombin deficiencies, and lupus anticoagulant, were eligible. Ethical approval was granted from the Department of Health and Science, France (2008-880 & 09.576) and all participants gave informed written consent, in accordance with the Declaration of Helsinki. As an ancillary genetic study, a subsample of 1,592 MARTHA patients has been typed by a high density genotyping array^12^. This sub-study was further extended over the 2013-2018 period and patients were re-contacted to gather information on post-inclusion VT events. Application of standard quality control procedures on the genome wide genotype data of MARTHA participants has led to the selection of 1,542 VT patients for genetic analyses. From these remaining individuals, we further excluded patients with autoimmune disease or cancer at inclusion, or with missing information on time to VT recurrence for concerned patients. Finally, 1,518 VT patients were left for the VT recurrence analysis, including 571 VT recurrences^13^.

#### MEGA

The **“Multiple Environmental and Genetic Assessment” (MEGA)** study includes almost 4,900 patients aged between 18-70 who were included for their first VT between 1999 and 2004 in the Netherlands^14^. Among them, 1,289 VT cases who were free of cancer and who provided a high-quality blood sample were eligible to have their DNA analysed and therefore had available genetic data as it has been previously described^15^. Between 2008 and 2009, questionnaires were sent to patients to gather information on a possible VT recurrence. After excluding patients with homozygosity for FV Leiden or Factor II 20210A, or missing information on VT recurrence, 1,254 patients including 431 VT recurrences were part of the analyses. The MEGA study was approved by the local ethic committee “Medical Ethics Committee of the Leiden University Medical Center”. All experimental protocols to study the genetics of VT recurrence were performed in accordance with the Declaration of Helsinki and approved by the local ethic committee “Medical Ethics Committee of the Leiden University Medical Center” for MEGA. Written informed consent to participate was obtained from all MEGA participants.

#### PADIS-PE

The **“Prolonged Anticoagulation During eighteen months vs placebo after Initial Six-month treatment for a first episode of idiopathic Pulmonary Embolism” (PADIS-PE)** is a clinical trial involving 371 incident cases of unprovoked pulmonary embolism aged over 18, enrolled between 2007-2012 in France^16^. After the end of the anticoagulant treatment period (6 or 18 months), all patients were regularly followed up for a median of 24 months. PADIS-PE study was conducted in accordance with the ethical principles stated in the Declaration of Helsinki, Good Clinical Practice, and relevant French regulations regarding ethics and data protection. The protocol and amendments were approved by a central independent ethics committee and written informed consent was obtained from all participants. All patients have been typed by a high density genotyping array. After applying standard quality controls and removing patients without missing information on VT recurrence, 155 participants including 26 VT recurrences were considered for the analyses.

#### REVERSE-I

The **“Recurrent Venous Thromboembolism** [**Risk Stratification**](https://www.sciencedirect.com/topics/medicine-and-dentistry/risk-stratification) **Evaluation” (REVERSE I)** is a multicentric study that comprises 646 patients aged over 18 who were enrolled between 2001 and 2006 after their first unprovoked VT. Participants were treated with >5 days of heparin or low-molecular-weight heparin after the index event, followed by 5–7 months of oral anticoagulants ^17^. Over a mean follow-up of 5 years, episodes of suspected recurrence were independently adjudicated. First unprovoked VT occurred in the absence of leg fracture or lower extremity plaster cast, immobilization for >3 days, surgery using general anesthetic 3 months prior to the index VT, or up to 5 years before the time of enrollment. Exclusion criteria included individuals under 18 years of age, who had already discontinued anticoagulant therapy, required ongoing anticoagulation for reasons other than VT, were geographically inaccessible for follow-up, were being treated for a recurrent unprovoked VT or a previously known high-risk thrombophilia (deficiency of protein S, C or antithrombin, known persistently positive anticardiolipin antibodies (>30U mL^−1^), a known persistently positive lupus anticoagulant, or who had two or more known thrombophilic defects (e.g. homozygous for FV Leiden or Factor II 20210A, or compound heterozygous for FV Leiden and Factor II 20210A)), or were unable to consent ^18^. From the genotyped samples, 451 participants of European origin were included in the analysis, 112 of whom experienced recurrence. Institutional research ethics board approvals were obtained by all participating centers (Ottawa Hospital Research Ethics Board). Protocols to study genetics in REVERSE I was approved by the University of Toronto Research Ethics Board.

### References

1.	3C Study Group. Vascular factors and risk of dementia: design of the Three-City Study and baseline characteristics of the study population. *Neuroepidemiology*. 2003;22(6):316–325.

2.	Duperron M-G, Knol MJ, Le Grand Q, et al. Genomics of perivascular space burden unravels early mechanisms of cerebral small vessel disease. *Nat Med*. 2023;29(4):950–962.

3.	Lind L, Ärnlöv J, Lindahl B, et al. Use of a proximity extension assay proteomics chip to discover new biomarkers for human atherosclerosis. *Atherosclerosis*. 2015;242(1):205–210.

4.	Lacut K, Oger E, Le Gal G, et al. Statins but not fibrates are associated with a reduced risk of venous thromboembolism: a hospital‐based case–control study. *Fundamemntal Clinical Pharma*. 2004;18(4):477–482.

5.	Kannel WB, Feinleib M, McNamara PM, Garrison RJ, Castelli WP. An investigation of coronary heart disease in families. The Framingham offspring study. *Am J Epidemiol*. 1979;110(3):281–290.

6.	Splansky GL, Corey D, Yang Q, et al. The Third Generation Cohort of the National Heart, Lung, and Blood Institute’s Framingham Heart Study: Design, Recruitment, and Initial Examination. *American Journal of Epidemiology*. 2007;165(11):1328–1335.

7.	Smith NL. Esterified Estrogens and Conjugated Equine Estrogens and the Risk of Venous Thrombosis. *JAMA*. 2004;292(13):1581.

8.	Smith NL, Hindorff LA, Heckbert SR, et al. Association of Genetic Variations With Nonfatal Venous Thrombosis in Postmenopausal Women. *JAMA*. 2007;297(5):489.

9.	Smith NL, Harrington LB, Blondon M, et al. The association of statin therapy with the risk of recurrent venous thrombosis. *Journal of Thrombosis and Haemostasis*. 2016;14(7):1384–1392.

10.	Oudot-Mellakh T, Cohen W, Germain M, et al. Genome wide association study for plasma levels of natural anticoagulant inhibitors and protein C anticoagulant pathway: the MARTHA project. *Br J Haematol*. 2012;157(2):230–239.

11.	Trégouët DA, Delluc A, Roche A, et al. Is there still room for additional common susceptibility alleles for venous thromboembolism? *J Thromb Haemost*. 2016;14(9):1798–1802.

12.	Lindström S, Wang L, Smith EN, et al. Genomic and transcriptomic association studies identify 16 novel susceptibility loci for venous thromboembolism. *Blood*. 2019;134(19):1645–1657.

13.	Munsch G, Goumidi L, van Hylckama Vlieg A, et al. Association of ABO blood groups with venous thrombosis recurrence in middle-aged patients: insights from a weighted Cox analysis dedicated to ambispective design. *BMC Med Res Methodol*. 2023;23(1):99.

14.	Chinthammitr Y, Vos HL, Rosendaal FR, Doggen CJM. The association of prothrombin A19911G polymorphism with plasma prothrombin activity and venous thrombosis: results of the MEGA study, a large population-based case-control study. *J Thromb Haemost*. 2006;4(12):2587–2592.

15.	de Haan HG, van Hylckama Vlieg A, Germain M, et al. Genome-Wide Association Study Identifies a Novel Genetic Risk Factor for Recurrent Venous Thrombosis. *Circulation: Genomic and Precision Medicine*. 2018;11(2):e001827.

16.	Couturaud F, Sanchez O, Pernod G, et al. Six Months vs Extended Oral Anticoagulation After a First Episode of Pulmonary Embolism: The PADIS-PE Randomized Clinical Trial. *JAMA*. 2015;314(1):31.

17.	Rodger MA, Kahn SR, Wells PS, et al. Identifying unprovoked thromboembolism patients at low risk for recurrence who can discontinue anticoagulant therapy. *Canadian Medical Association Journal*. 2008;179(5):417–426.

18.	Rodger MA, Scarvelis D, Kahn SR, et al. Long-term risk of venous thrombosis after stopping anticoagulants for a first unprovoked event: A multi-national cohort. *Thrombosis Research*. 2016;143:152–158.
