## Supplementary Note for "Genomic Landscape of Thrombosis Recurrence Risk Across Venous Thromboembolism Subtypes"

In this work, we looked for genetic loci associated with Venous Thromboembolism (VT) recurrence, that may differ from those associated with VT incidence. Studies of disease progression are, by design, prone to be affected by index event bias especially if there are shared risk factors between disease and progression^1^. Recently, strategies have been proposed to detect, quantify and overcome index event bias in genetic studies^2^.

To identify if the association of some SNPs with VT recurrence may be biased due to index event, a Miami plot can be created. Using GWAS summary statistics from *Thibord et al. (2022)* we visually assessed whether there are some shared variants between index (VT) and subsequent (VT recurrence) events (**Supplementary Note Figure 1**). As presented in **Supplementary Table S23**, genetic loci significantly (P<5×10^-8^ or P<8×10^-9^, if subgroups are considered) associated with VT recurrence are not associated with 1^st^ VT. Consequently these variants do not suffer from index event bias^1^.

By contrast, as some genetic loci for the VT incidence are marginally associated with VT recurrence (**Table 3**), we decided to assess the sensibility of our results using methods to overcome index event bias.

First, we used Inverse Probability Weighting (IPW) method, a well-known method implemented in epidemiological studies to account for sample selection bias^3^. In our context, the IPW method assigns a weight to each individual that represents their probability of not developing VT (the index event). As such, individuals with a high VT probability are down-weighted (their statistical contribution to the likelihood is reduced) and individuals with a low VT probability are up-weighted. The underlying hypothesis is that our studied samples are enriched in individuals with a higher probability of being VT patients which may result in identifying associations that are only due to VT and not specific to VT recurrence.

In a secondary analysis, we used the method proposed by *Dudbridge et al. (2019)* that can directly be applied using summary statistics^2^.

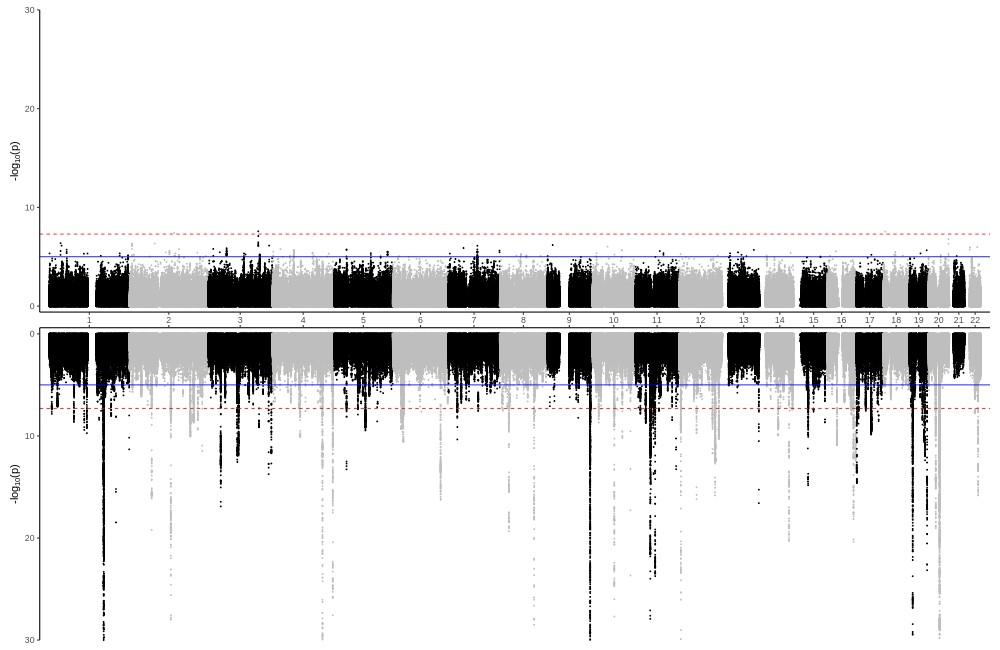

**Supplementary Note Figure 1:** Miami plot of GWAS on VT and VT recurrence

**Figure legend:** This Miami plot displays on the top of the graph GWAS results from the meta-analysis on VT recurrence, and on the bottom GWAS results on 1^st^ VT from *Thibord et al. (2022)*. The miami plot is interpreted in the same way as a Manhattan plot, except that the x-axis representing the genomic position is scaled to facilitate visual comparison of loci.

**Results**

To obtain the probabilities of having a VT that is necessary to implement the IPW method, we first fitted a logistic regression model for VT incidence in the FARIVE case-control study that did not contribute to our study for VTE recurrence. FARIVE is a case-control French multicentre study that includes ~600 VT cases and ~600 hospital controls. FARIVE study has already been described and genotypic details are provided elsewhere^4,5^. Using 95 genetic loci associated with VT in *Thibord et al. (2022)*^6^ and their respective additive allele effects that were available in FARIVE, we derived a Genetic Risk Score (GRS) for VT. The explanatory variables of the regression model for VT incidence were age, sex and the GRS for VT. Consistently with the literature, the fitted model has an Area Under the Curve (AUC) of 0.708 [0.677-0.738] ^7^ and the estimated odds ratio (OR) for VT were OR=4.57 [3.50-6.05]; P=1.23×10^-27^, OR=1.26 [0.97-1.65]; P=0.086, OR=1.16 [1.15-1.16]; P=5.27×10^-5^, respectively for GRS of VT, female sex and increase of 10 years of age.

Given the estimated association parameters obtained in FARIVE, we computed for each participant of the EDITH and MEGA studies their probabilities of being VT patients according to their individual data on GRS, sex and age. These probabilities were then inversed and used as weights in the Cox proportional hazard models. We compared the associations on VT recurrence, with and without the IPW method, for the main SNPs identified in the meta-analysis for VT recurrence as well as for the eight SNPs with the strongest effect on the risk of VT (**Supplementary Note Table 1**). For the SNPs of VT recurrence, we observed some very minor fluctuations in the estimations when using the IPW method, but the effect sizes remain unchanged. The same pattern is also observed for the eight SNPs of 1^st^ VT, although the IPW method seems to slightly enhance the trends for association observed.

To assess the presence of index event bias on the meta-analysis, we applied the Dudbridge method on the summary statistics for VT recurrence using the summary statistics for 1^st^ VT from *Thibord et al. (2022)*. After harmonization of the effect allele in both summary statistics, we identified a set of independent SNPs which have been tested in both studies^8^. For this, we used the same parameters as in *Dudbridge et al. (2019)*: 1000Genomes Phase3 Version5 reference panel; PLINK 1.9 with *R*^2^ threshold of 0.1 within 250 SNP windows. Finally, 238,732 SNPs were kept to estimate the regression slope between the association parameters for the index and the subsequent phenotype that was then used to correct for index event bias the summary statistics for VT recurrence. Results from this sensitivity analysis are presented in **Supplementary Note Table 2**. As expected, this correction had no impact on the four SNPs of VT recurrence, that were not associated with 1^st^ VT. However, surprising results were observed for the eight SNPs associated with 1^st^ VT. For example, the rs6025-T that was strongly associated with increased incident VT risk (OR=2.94, P=1.57×10^-1028^) showed a very strong negative association with VT recurrence (HR=0.43, P=3.25×10^-36^) using the Dudbridge method while it showed a nominal positive association (HR=1.15, P=0.04) in the original uncorrected analysis. The same patterns were observed for the eight SNPs strongly associated with 1^st^ VT. The results provided by the Dudbridge methods for the eight SNPs strongly associated with incident VT were inconsistent with those obtained with the IPW method. Given that the Dudbridge method has been described to be unreliable in the presence of strong effects for the index event^2^ and, according to our IPW results, we concluded that our summary statistics for VT recurrence did not suffer from index event bias, as well as the derived analyses.

**Supplementary Note Table 1:** Associations on VT recurrence, with and without the IPW method, for the main SNPs identified in the meta-analysis for VT recurrence as well as for the eight SNPs with the strongest effect on the risk of VT in the EDITH and MEGA studies.

|  |  |  | **EDITH** | | **EDITH with IPW** | | **MEGA** | | **MEGA with IPW** | |
| --- | --- | --- | --- | --- | --- | --- | --- | --- | --- | --- |
|  | **CHR:POS:NEA:EA** | **Gene** | **HR [95% CI]** | **Pvalue** | **HR [95% CI]** | **Pvalue** | **HR [95% CI]** | **Pvalue** | **HR [95% CI]** | **Pvalue** |
| **Variants associated with VT recurrence** | 3:154545386:T:C | *GPR149;MME* | 1.32 [0.80-2.18] | 0.272 | 1.49 [0.83-2.66] | 0.180 | 2.85 [1.91-4.25] | 2.53x10^-7^ | 3.10 [1.99-4.82] | 5.33x10^-7^ |
|  | 18:6214286:C:T | *L3MBTL4* | 1.87 [1.08-3.24] | 0.025 | 2.09 [1.13-3.83] | 0.018 | 1.33 [0.45-3.88] | 0.608 | 1.53 [0.61-3.84] | 0.371 |
|  | 2:137678403:G:T | *THSD7B* | 1.21 [0.64-2.31] | 0.554 | 1.02 [0.48-2.16] | 0.959 | 1.18 [0.66-2.10] | 0.572 | 1.13 [0.61-2.10] | 0.699 |
|  | 20:62463797:G:A | *GATA5* | 1.41 [0.90-2.22] | 0.138 | 1.47 [0.90-2.38] | 0.121 | 1.85 [1.21-2.83] | 4.27x10^-3^ | 2.15 [1.41-3.27] | 3.59x10^-4^ |
| **Main variants associated with 1^st^ VT** | 1:169549811:T:C | *F5* | 0.88 [0.66-1.18] | 0.389 | 0.77 [0.57-1.02] | 0.069 | 0.61 [0.48-0.77] | 2.76x10^-5^ | 0.57 [0.45-0.72] | 2.70x10^-6^ |
|  | 9:133261703:G:A | *ABO* | 1.08 [0.93-1.27] | 0.312 | 1.17 [0.99-1.37] | 0.053 | 1.16 [1.01-1.33] | 0.040 | 1.17 [0.99-1.36] | 0.053 |
|  | 4:186285095:C:A | *F11* | 1.24 [1.07-1.43] | 4.33x10^-3^ | 1.28 [1.09-1.49] | 2.34x10^-3^ | 1.17 [1.02-1.35] | 0.029 | 1.22 [1.04-1.43] | 0.017 |
|  | 4:154604543:G:A | *FGG* | 1.02 [0.87-1.20] | 0.762 | 1.10 [0.93-1.31] | 0.251 | 1.31 [1.14-1.51] | 2.12x10^-4^ | 1.31 [1.12-1.53] | 8.76x10^-4^ |
|  | 11:46739505:G:A | *F2* | 1.22 [0.79-1.89] | 0.363 | 1.30 [0.84-2.04] | 0.242 | 1.02 [0.34-3.06] | 0.975 | 1.47 [0.58-3.75] | 0.417 |
|  | 10:69485520:T:C | *TSPAN15* | 1.04 [0.77-1.41] | 0.781 | 0.99 [0.72-1.36] | 0.949 | 0.88 [0.69-1.12] | 0.296 | 0.89 [0.68-1.17] | 0.404 |
|  | 20:35189809:G:C | *PROCR* | 1.11 [0.95-1.28] | 0.189 | 1.10 [0.93-1.29] | 0.274 | 1.03 [0.89-1.18] | 0.714 | 1.05 [0.89-1.23] | 0.563 |
|  | 19:10628160:A:G | *SLC44A2* | 1.09 [0.90-1.32] | 0.370 | 1.11 [0.90-1.37] | 0.312 | 1.17 [0.96-1.43] | 0.120 | 1.13 [0.90-1.41] | 0.282 |

*VT: Venous thromboembolism / CHR: Chromosome / POS: Position in Hg38 / NEA: Non-effect allele / EA: Effect allele / HR: Hazard Ratio / CI: Confidence Interval / IPW: Inverse Probability Weighting.*

**Supplementary Note Table 2:** Associations on 1^st^ VT and VT recurrence, with and without Dudbridge correction for index event bias method, for the main SNPs identified in the meta-analysis for VT recurrence as well as for the eight SNPs with the strongest effect on the risk of VT.

|  |  |  | **1^st^ VT (*Thibord et al. 2022*)** | | **VT recurrence** | | **VT recurrence with Dudbridge correction** | |
| --- | --- | --- | --- | --- | --- | --- | --- | --- |
|  | **CHR:POS:NEA:EA** | **Gene** | **OR [95% CI]** | **Pvalue** | **HR [95% CI]** | **Pvalue** | **HR [95% CI]** | **Pvalue** |
| **SNPS associated with VT recurrence** | 3:154545386:T:C | *GPR149;MME* | 1.00 [0.97-1.04] | 0.959 | 1.84 [1.49-2.29] | 2.65x10^-8^ | 1.84 [1.49-2.29] | 3.79x10^-8^ |
|  | 18:6214286:C:T | *L3MBTL4* | 1.02 [0.96-1.09] | 0.537 | 2.16 [1.65-2.83] | 2.82x10^-8^ | 2.12 [1.60-2.80] | 1.19x10^-7^ |
|  | 2:137678403:G:T | *THSD7B* | 0.99 [0.94-1.04] | 0.741 | 1.98 [1.55-2.52] | 3.83x10^-8^ | 1.99 [1.56-2.52] | 4.52x10^-8^ |
|  | 20:62463797:G:A | *GATA5* | 1.06 [1.01-1.11] | 0.028 | 1.75 [1.42-2.16] | 1.68x10^-7^ | 1.67 [1.35-2.06] | 2.88x10^-6^ |
| **Main SNPS associated with 1^st^ VT** | 1:169549811:T:C | *F5* | 0.34 [0.33-0.35] | 1.57x10^-1028^ | 0.87 [0.77-0.99] | 0.038 | 2.31 [2.03-2.63] | 3.25x10^-36^ |
|  | 9:133261703:G:A | *ABO* | 1.34 [1.32-1.35] | 7.25x10^-507^ | 1.11 [1.04-1.19] | 2.19x10^-3^ | 0.86 [0.80-0.92] | 1.44x10^-5^ |
|  | 4:186285095:C:A | *F11* | 1.22 [1.21-1.23] | 1.15x10^-238^ | 1.09 [1.02-1.17] | 0.013 | 0.91 [0.85-0.98] | 9.87x10^-3^ |
|  | 4:154604543:G:A | *FGG* | 1.23 [1.22-1.25] | 8.63x10^-218^ | 1.14 [1.07-1.23] | 2.32x10^-4^ | 0.95 [0.88-1.02] | 0.170 |
|  | 11:46739505:G:A | *F2* | 1.99 [1.90-2.09] | 8.44x10^-170^ | 1.43 [1.12-1.81] | 3.40x10^-3^ | 0.77 [0.61-0.98] | 0.035 |
|  | 10:69485520:T:C | *TSPAN15* | 0.81 [0.79-0.82] | 1.02x10^-111^ | 0.90 [0.79-1.01] | 0.082 | 1.09 [0.96-1.23] | 0.197 |
|  | 20:35189809:G:C | *PROCR* | 1.12 [1.10-1.13] | 9.06x10^-68^ | 1.04 [0.97-1.11] | 0.306 | 0.94 [0.87-1.00] | 0.065 |
|  | 19:10628160:A:G | *SLC44A2* | 1.12 [1.10-1.14] | 9.19x10^-55^ | 1.10 [1.00-1.20] | 0.045 | 0.99 [0.91-1.09] | 0.875 |

*VT: Venous thromboembolism / CHR: Chromosome / POS: Position in Hg38 / NEA: Non-effect allele / EA: Effect allele / HR: Hazard Ratio / CI: Confidence Interval / OR: Odds Ratio.*

### References

1.	Mitchell RE, Hartley AE, Walker VM, et al. Strategies to investigate and mitigate collider bias in genetic and Mendelian randomisation studies of disease progression. *PLoS Genet*. 2023;19(2):e1010596.

2.	Dudbridge F, Allen RJ, Sheehan NA, et al. Adjustment for index event bias in genome-wide association studies of subsequent events. *Nat Commun*. 2019;10(1):1561.

3.	Seaman SR, White IR. Review of inverse probability weighting for dealing with missing data. *Stat Methods Med Res*. 2013;22(3):278–295.

4.	Trégouët D-A, Heath S, Saut N, et al. Common susceptibility alleles are unlikely to contribute as strongly as the FV and ABO loci to VTE risk: results from a GWAS approach. *Blood*. 2009;113(21):5298–5303.

5.	Munsch G, Proust C, Labrouche-Colomer S, et al. Genome-wide association study of a semicontinuous trait: illustration of the impact of the modeling strategy through the study of Neutrophil Extracellular Traps levels. *NAR Genom Bioinform*. 2023;5(2):lqad062.

6.	Thibord F, Klarin D, Brody JA, et al. Cross-Ancestry Investigation of Venous Thromboembolism Genomic Predictors. *Circulation*. 2022;146(16):1225–1242.

7.	Jee YH, Thibord F, Dominguez A, et al. Multi-ancestry polygenic risk scores for venous thromboembolism. *Hum Mol Genet*. 2024;ddae097.

8.	Chang CC, Chow CC, Tellier LC, et al. Second-generation PLINK: rising to the challenge of larger and richer datasets. *GigaSci*. 2015;4(1):7.
